# First Insights into microbial changes within an Inflammatory Bowel Disease Family Cohort study

**DOI:** 10.1101/2024.07.23.24310327

**Authors:** Philipp Rausch, Ilka Ratjen, Lukas Tittmann, Janna Enderle, Eike Matthias Wacker, Kathrin Jaeger, Malte Christoph Rühlemann, Katrin Franzpötter, Pierre Ellul, Robert Kruse, Jonas Halfvarson, Dirk Roggenbuck, David Ellinghaus, Gunnar Jacobs, Michael Krawczak, Stefan Schreiber, Corinna Bang, Wolfgang Lieb, Andre Franke

## Abstract

**Background:** The prospective Kiel Inflammatory Bowel Disease (IBD) Family Cohort Study (KINDRED cohort) was initiated in 2013 to systematically and extensively collect data and biosamples from index IBD patients and their relatives (e.g., blood, stool), a population at high risk for IBD development. Regular follow-ups were conducted to collect updated health and lifestyle information, to obtain new biosamples, and to capture the incidence of IBD during development. By combining taxonomic and imputed functional microbial data collected at successive time points with extensive anthropometric, medical, nutritional, and social information, this study aimed to characterize the factors influencing the microbiota in health and disease via detailed ecological analyses.

**Results:** Using two dysbiosis metrics (MD-index, GMHI) trained on the German KINDRED cohort, we identified strong and generalizable gradients within and across different IBD cohorts, which correspond strongly with IBD pathologies, physiological manifestations of inflammation (*e.g.,* Bristol stool score, ASCA IgA/IgG), genetic risk for IBD, and general risk of disease onset. Anthropometric and medical factors influencing transit time strongly modify bacterial communities. Various *Enterobacteriaceae* (*e.g., Klebsiella sp.*) and opportunistic *Clostridia* pathogens (*e.g., C. XIVa clostridioforme*), characterize in combination with ectopic oral taxa (*e.g. Veillonella sp.*, *Cand. Saccharibacteria sp.*, *Fusobacterium nucleatum*) the distinct and chaotic IBD-specific communities. Functionally, amino acid metabolism and flagellar assembly are beneficial, while mucolytic functions are associated with IBD.

**Conclusions:** Our findings demonstrate broad-scale ecological patterns which indicate drastic state transitions of communities into characteristically chaotic communities in IBD patients. These patterns appear to be universal across cohorts and influence physiological signs of inflammation, display high resilience, but show only little heritability/intrafamily transmission.

## INTRODUCTION

Inflammatory bowel disease (IBD), with the most common manifestations of Crohn’s disease (CD) and ulcerative colitis (UC), is characterized by chronic, relapsing inflammation of the gastrointestinal tract [1] resulting from a complex interplay of genetic, lifestyle, and environmental factors [2–4]. The incidence and prevalence of IBD are rising in many parts of the world [5,6], and the highest incidences are currently reported in North America and Europe [6,7], while short-term projections expect a prevalence of 1% in high-income countries by the year 2030 [8].

Clustering of IBD cases within families is not uncommon, suggesting a significant contribution of shared genes and lifestyle or environmental factors to IBD susceptibility [4,9–12]. First-degree relatives of IBD patients have a 3- to 20-fold greater risk of developing the disease themselves than does the general population [4,11,13,14]. In particular, for siblings of IBD patients, estimates of the relative disease risk range from 10 to 50 [15]. Meta-analyses and genome-wide association studies (GWAS) have identified 320 loci associated with either CD or UC or both [16]. Most disease-related genes and pathways are thought to influence disease risk by altering the epithelial barrier, the gut microbiota, or the inflammatory process in general [4,17–20]. Moreover, familial aggregation of IBD has been suggested to be a risk factor for a more aggressive course of the disease, earlier disease onset, and greater need for treatment [21,22], while disease risk in families with IBD cases is generally increased [23] and can be further evaluated by polygenic risk scores derived from the accumulated genetic knowledge of the disease [24].

Most prior studies on IBD risk factors used a classical case‒control design, comparing IBD patients to healthy controls. However, the molecular alterations or lifestyle factors that precede or predict the pre-clinical onset of IBD can only be identified reliably in prospective studies that collect health-related information and biomaterials from unaffected or preclinical individuals over time. Given the relatively low IBD incidence in the general population and the high familial risk for IBD, a family-based prospective study represents a more efficient study design than a population-based study. However, due to the high costs of maintenance, infrastructure, participant commitment, and long run-time, this type of study design is rare but offers great potential [23]. Thus, in 2013, the Inflammatory Bowel Disease Family Cohort (KINDRED cohort), the so-called KINDRED cohort, was initiated in Kiel (Germany) to systematically collect disease-relevant data from IBD patients and their healthy relatives. The main objectives of the study were (i) to identify lifestyle factors and biomarkers that predispose patients to, or predict, the development of IBD and (ii) to characterize the long-term clinical course of IBD patients. To this end, comprehensive and high-quality biomaterial (blood, stool, hair) as well as detailed health-related, lifestyle, sociodemographic, and kinship information was collected from the participants of this study. Regular follow-ups are performed (approximately every 2 years), which are embedded into a long-term follow-up program for healthy relatives with respect to IBD onset.

One main driver of IBD has been identified in the microbial community inhabiting the intestinal tract. Various human population studies and animal experiments have identified a multitude of ways in which the microbiome varies and interacts with its host, eventually leading to and becoming indispensable for the development of intestinal inflammation [25]. One current hypothesis is that dysbiosis, a shift in the balance of a beneficial microbial community as a whole in favor of a proinflammatory community, eventually leads to and sustains intestinal inflammation via metabolic and immunologic alterations but not single taxa [25]. These community shifts can result from disturbances introduced through environmental and lifestyle factors, diet, hygiene, medication, or even the host’s genetic makeup [25–27]. However, the development of inflammation is a nonlinear and often long-lasting process that can persist preclinical (perceived healthy), although inflammation may already fester [28]. Analyzing early changes, particularly in microbial communities, and reducing the influence of treatment-related disturbances offers immense opportunities to trace the true pathological dynamics of the microbiota during the development of IBD. Thus, recent efforts in predicting IBD in healthy or preclinical individuals have successfully been employed to identify dedicated transcriptional [29], physiological [30], and microbial markers that are broadly associated with disease onset in IBD [31,32].

## RESULTS

### Current status of the KINDRED cohort

As of April 2021, the Kiel KINDRED cohort included 1497 IBD patients and 1813 initially non-affected family members, belonging to a total of 1372 families. The data of all study participants whose baseline questionnaires were double entered and quality-checked were extracted from the database in a data freeze in March 2021, comprising 2393 individuals (1321 IBD patients and 1072 non-affected relatives). The discrepancy in the total number of study participants is due to the time lag between study inclusion and the return of biomaterial and questionnaire data (plus quality check). The baseline characteristics of the healthy participants, stratified by age, and of the IBD patients, stratified by disease subtype (CD, UC, uIBD), are summarized in Table 1 and Suppl. Tables S3 & S4, respectively.

**Table 1:**
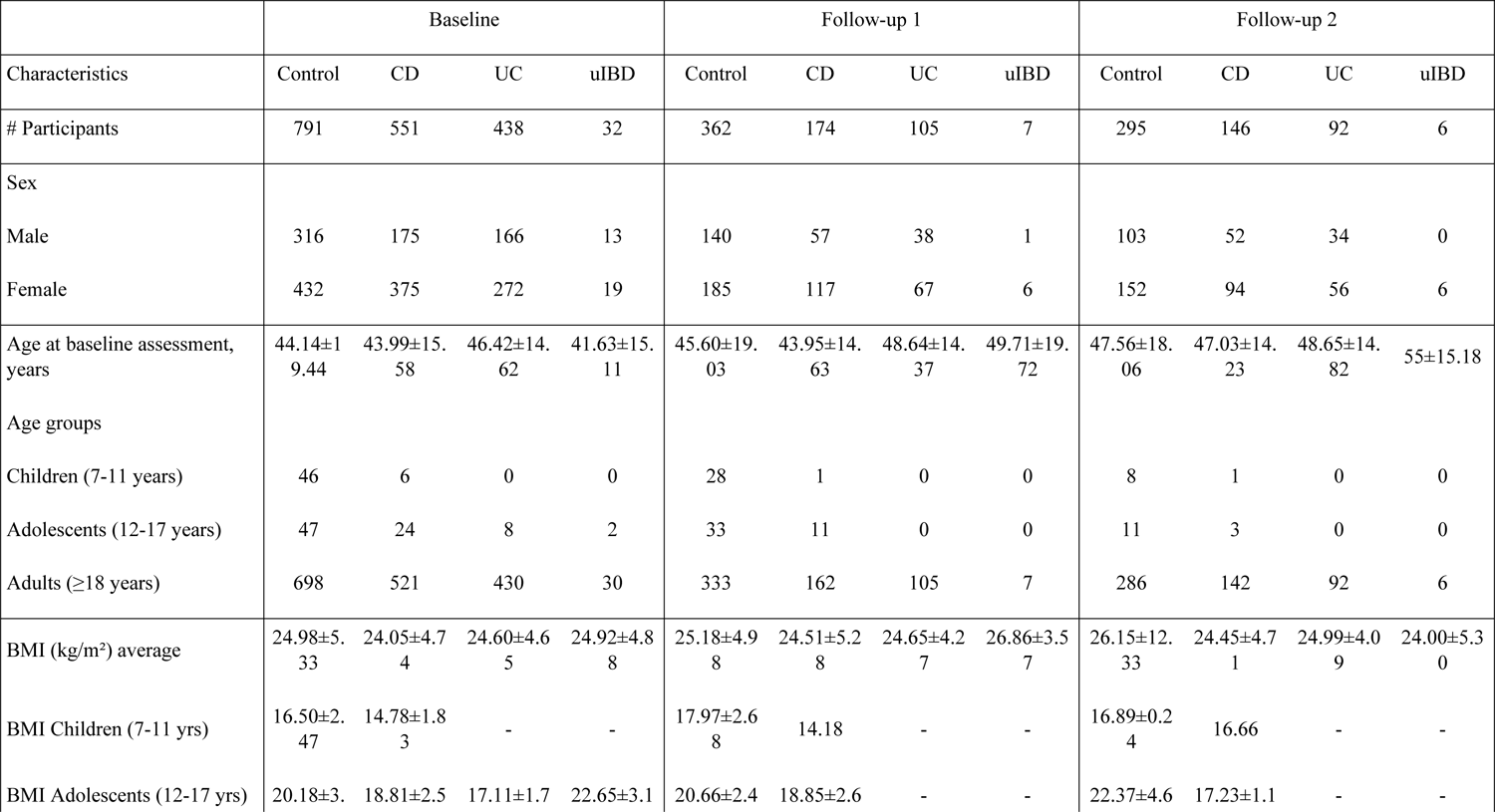
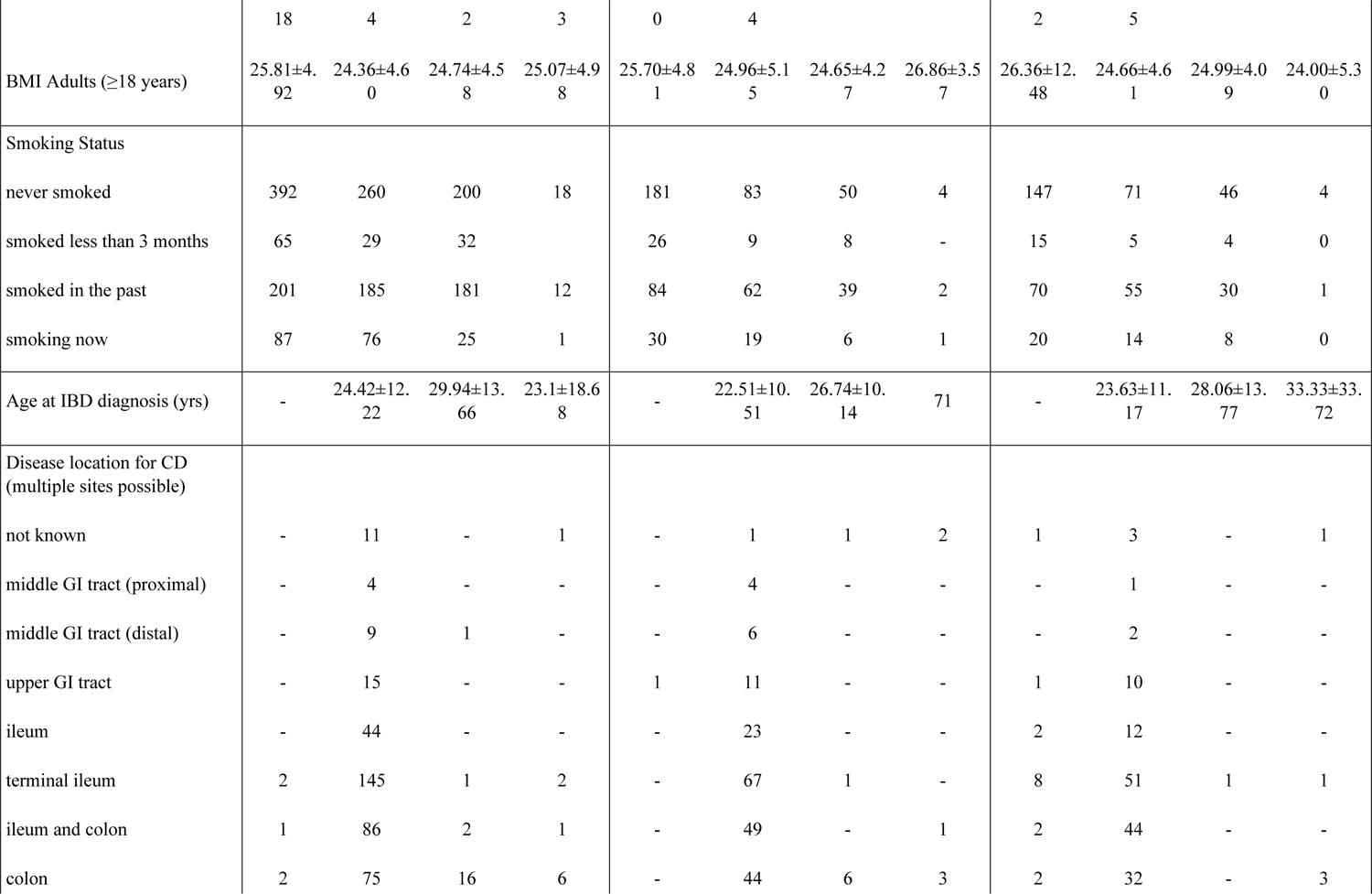
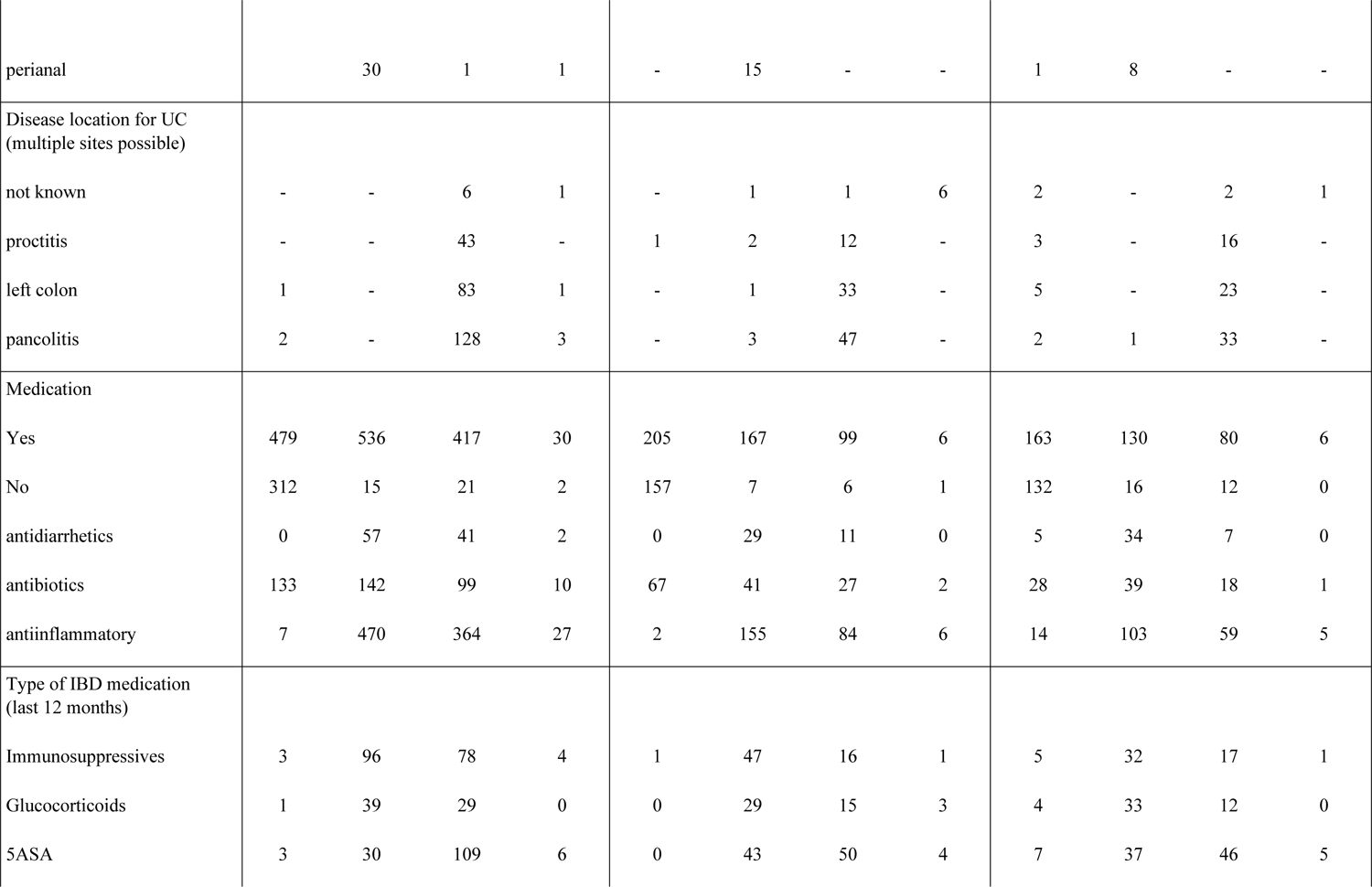
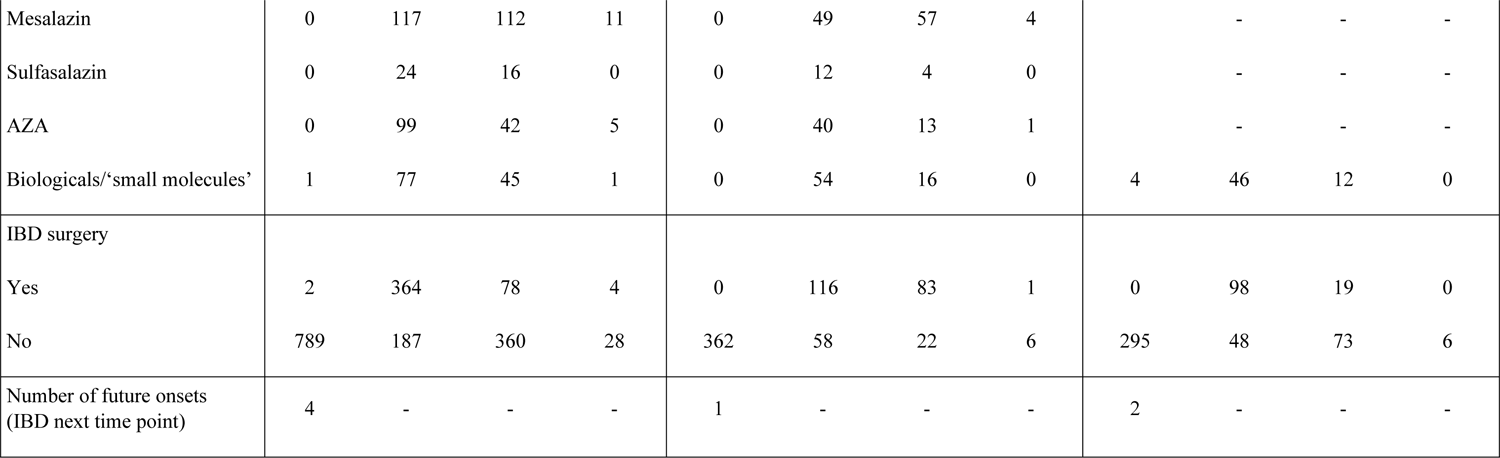
Baseline characteristics of the IBD patients (N_individuals_=1868) of the Kiel IBD Family Cohort. Values are mean ± standard deviation or absolute counts. Abbreviations: CD, Crohn’s disease; IBD, inflammatory bowel disease; UC, Ulcerative colitis, uIBD, undefined inflammatory bowel disease.

Available biosamples from the baseline collection were used to generate data for biomarker discovery, including genome-wide genotyping (Global Screening Array (GSA), version 2.0 (Illumina), N=1725). These data are complemented by fecal 16S rRNA-based microbiome profiles (N=1812), blood biomarker profiles (C-reactive protein, N=316; hemoglobin, N=49; mainly IBD patients), serum antibodies (ASCA IgA, N=781; ASCA IgG, N=780; anti-GP2 IgA, N=781; anti-GP2 IgG, N=781), and fecal indicators of inflammation (calprotectin, N=1763; occult hemoglobin/haptoglobin status, N=1760; Bristol stool score, N=882). For a subset from the first and second follow-ups (N_F1_=648, N_F2_=539), 16S rRNA sequencing data, fecal calprotectin (N_F1_=519, N_F2_=147), hemoglobin/haptoglobin measurements (N_F1_=516, N_F2_=144), Bristol stool score (N_F1_=316, N_F2_=255) and blood biomarkers for inflammation (Hb: NF1=203, NF2=162; CRP: NF1=179, NF2=127) were also available. Our study set included 7 cases of IBD onset (four in the BL → F1 group, one in the F1 → F2 group, and two in the F2 → F3 group; see Figure 1A).

**Figure 1:**
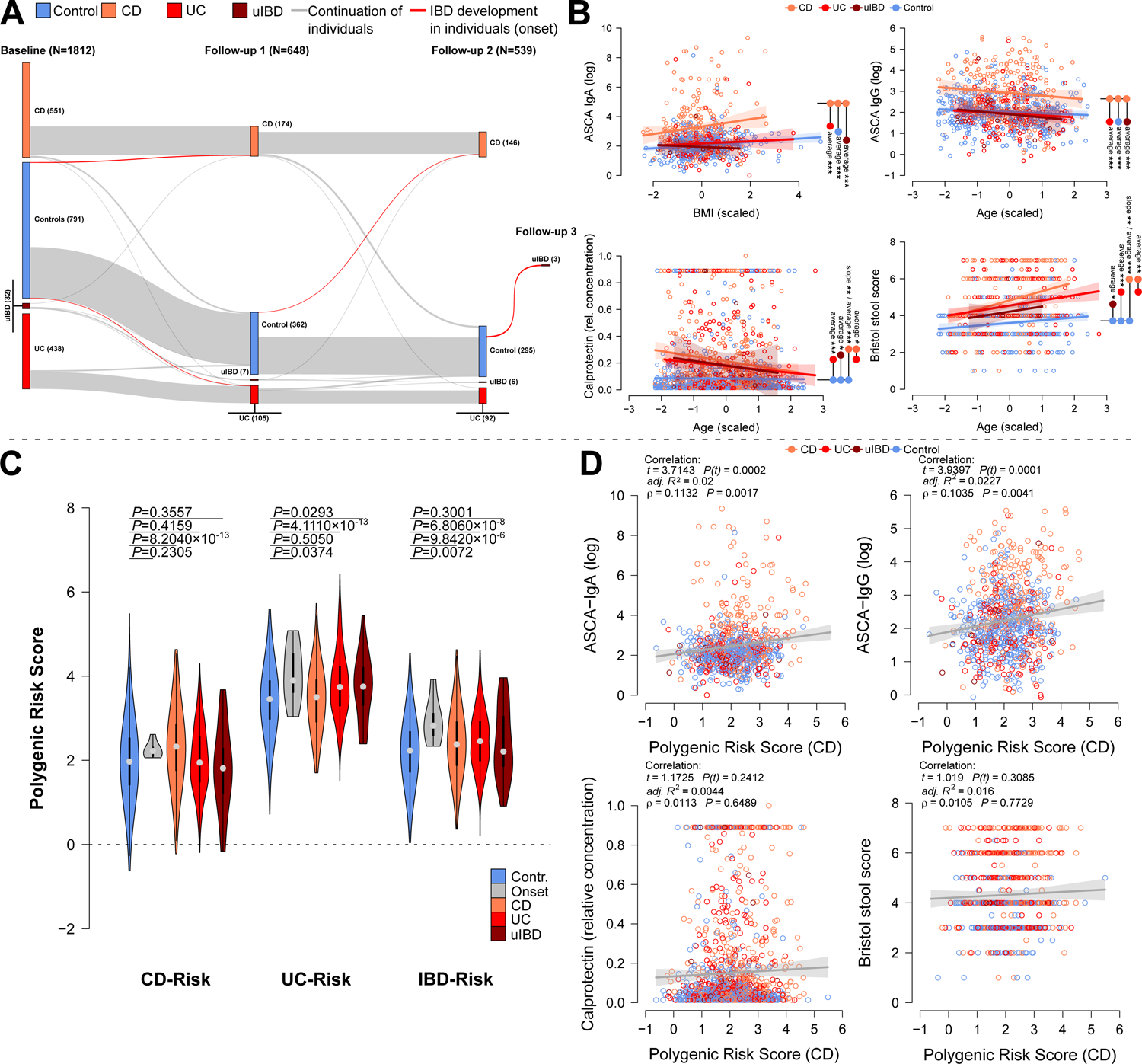
**(A)** Individual transitions in the cohort between time points. The highlighted transitions are onset cases detected during the study’s runtime (N_BL→F1_=4, N_F1→F2_=1, N_F2→F3_=2,, N_F3→F4_=3). **(B)** Analysis of selected physiological inflammation markers (ASCA IgA/IgG, Calprotectin, BSS) with respect to IBD condition and relevant covariates via linear models at baseline (see Table 1). Shown are only the optimal model results, after the model selection procedures are shown. Pairwise differences with respect to average differences, or differences in slope are highlighted in the plots (# *P*≤0.1000, * *P*≤0.0500, ** *P*≤0.0100, *** *P*≤0.0010). **(C)** The violin plot displays the average differences of genetic predisposition to CD, UC and IBD in general, as based on *LDpred2* derived PRS. The average differences in PRS were tested via Wilcoxon rank tests contrasting healthy individuals, with healthy future onset cases (grey), CD patients (red), UC patients, and patients with undefined IBD (uIBD). In particular, compared with those in healthy controls, the average risk of IBD is significantly and consistently greater in patients with CD and UC, as well as in patients with future-onset disease. **(D)** Scatterplots show the significant relationships between polygenic risk scores for CD (PRS-CD) and selected physiological inflammation markers,between PRS-CD and ASCA IgG by considering anthropometric covariates (linear model on residuals) or without (Spearman correlation). Additional correlations of IBD- and UC-PRS with physiological markers can be found in Figure S4 and Table S5.

### Distribution of physiological and clinical biomarkers of inflammation in the KINDRED cohort

We detected several highly interesting patterns among the biomarkers of inflammation, but our analyses focused on samples from the baseline examination because of the large number of samples and biomarkers. In the baseline samples, the concentrations of the antifungal antibodies anti-*Saccharomyces cerevisiae* antibody (ASCA) IgG and IgA decreased with age and increased with BMI (Table 2). ASCA antibody levels were greater in CD patients than in healthy controls and UC patients (Figure 1B). Furthermore, the concentration of anti-glycoprotein 2 IgA and IgG (GP2), which have previously been reported to be reliable biomarkers for PSC and CD [33–35], significantly differ between healthy relatives and IBD patients, and are significantly associated with patient age, BMI and sex, with the concentration of anti-GP2 Ig’s decreasing in females and increasing in males. We detected a particularly high average concentration in CD patients compared to either that in UC patients or non-IBD control subjects (Table 2). The fecal calprotectin concentration, a general measure of inflammatory processes in the gut and a metal sequestering protein, decreases with age in IBD patients but significantly increases in CD patients, followed by UC patients (Figure 1B). The concentration of C-reactive protein (CRP), similar to hemoglobin (Hb) levels, both measured only in diseased individuals, increased with BMI, while Hb levels in affected males were significantly greater than for women with IBD (Figure S4). Stool consistency, as expressed by the self-reported Bristol stool score (BSS), strongly and significantly increases with age. This age-related increase in BSS was particularly strong in CD patients (Figure 1B; Table 2). This indicates that older CD patients suffer from greater bowel irritation and thus shorter intestinal passage times. At other examination time points, the associations were mostly comparable to the associations in the baseline analyses, as reported in Table 2.

**Table 2:**
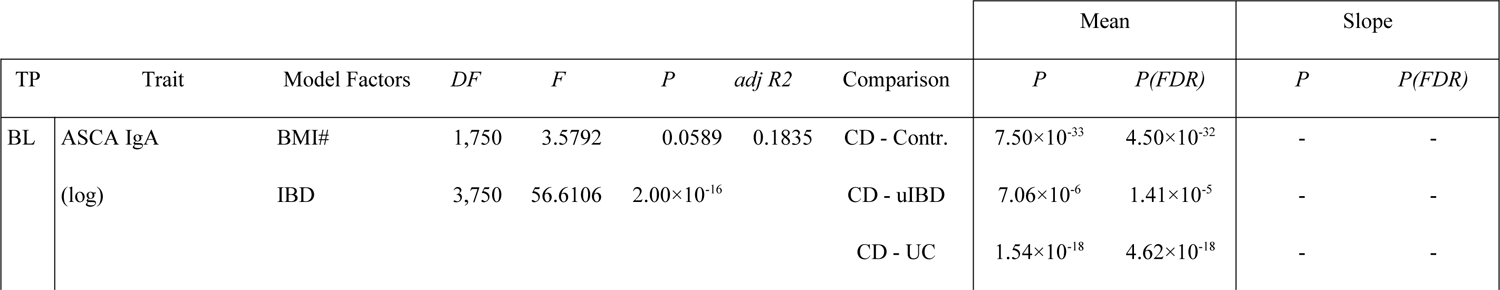
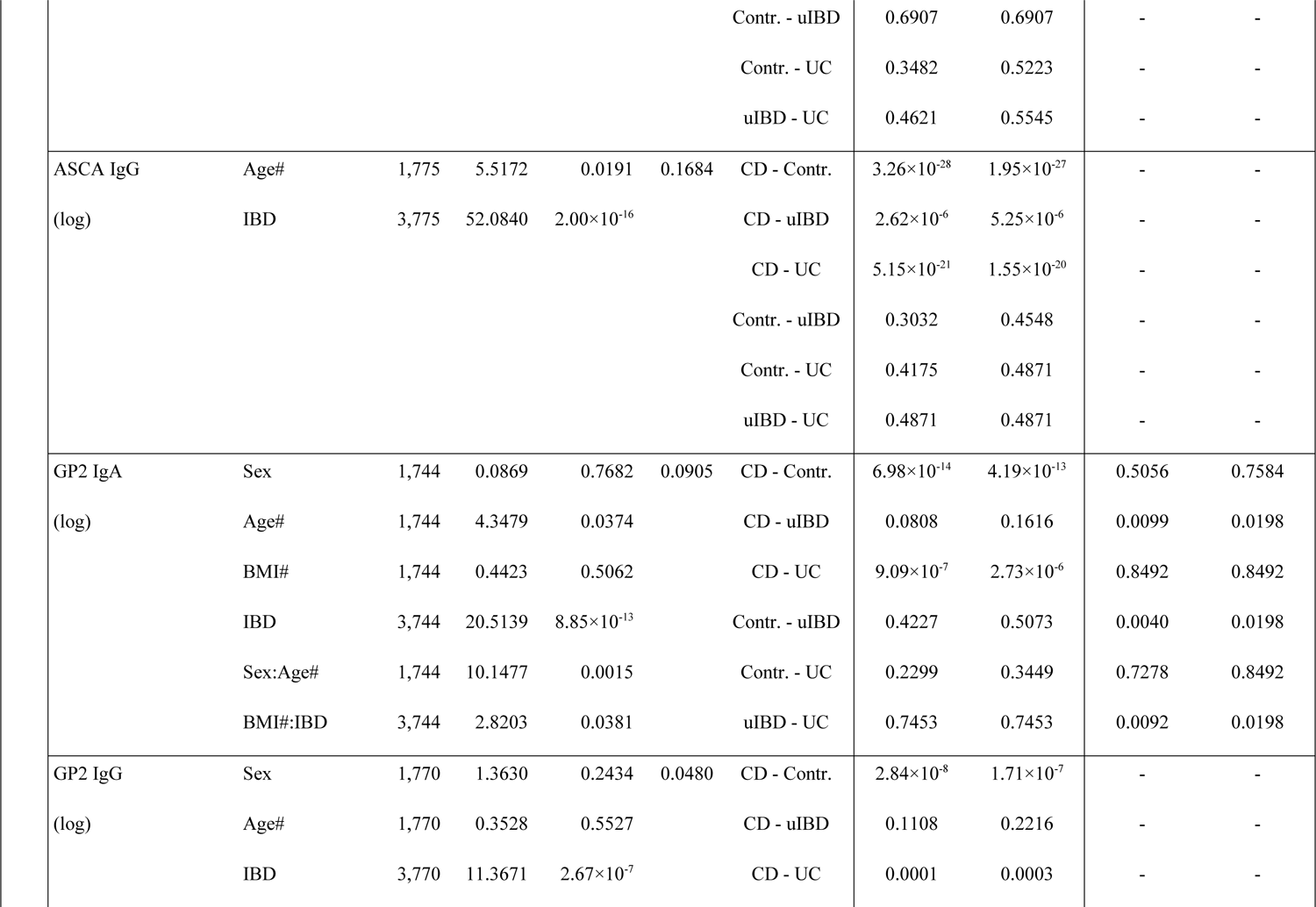
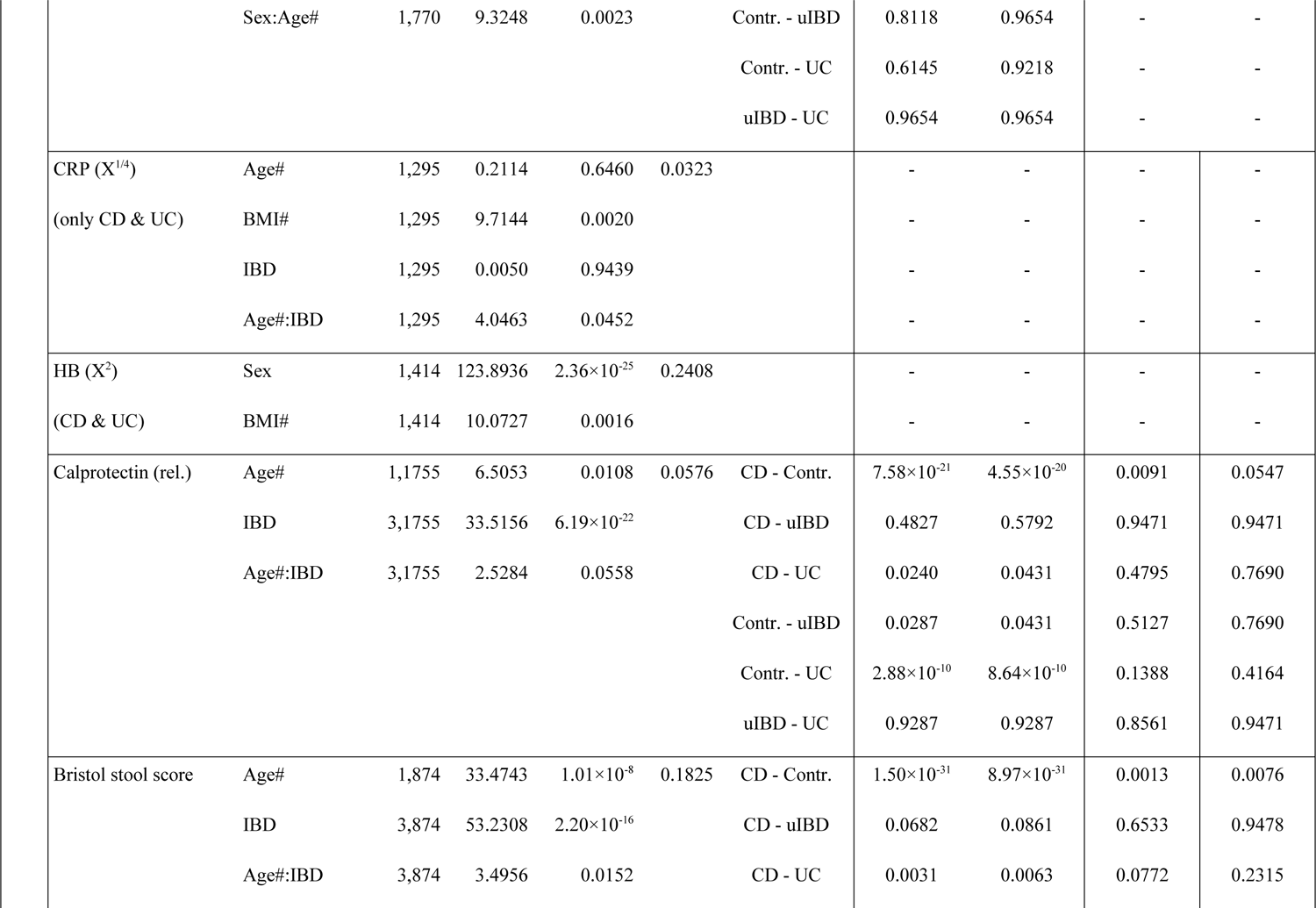
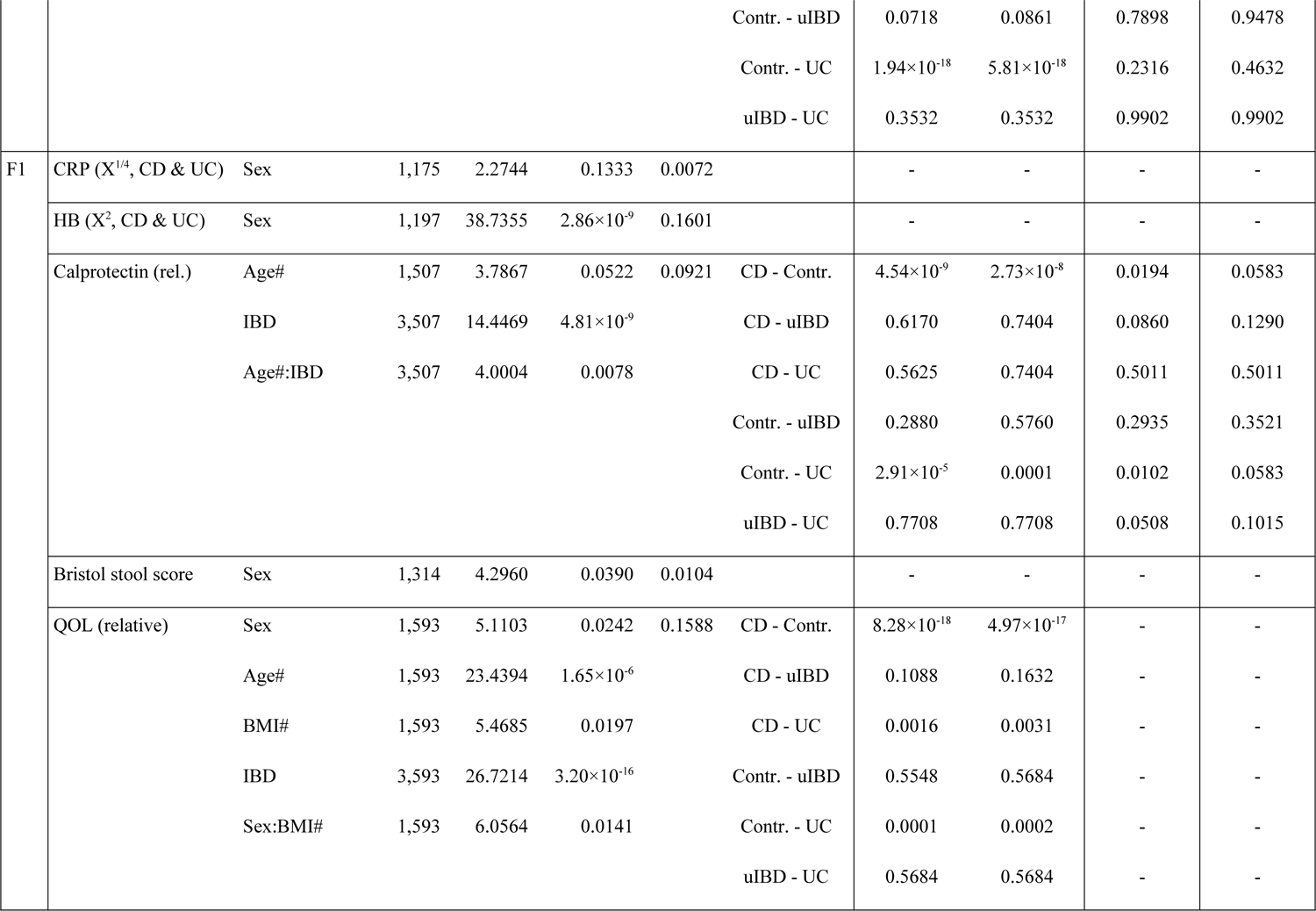
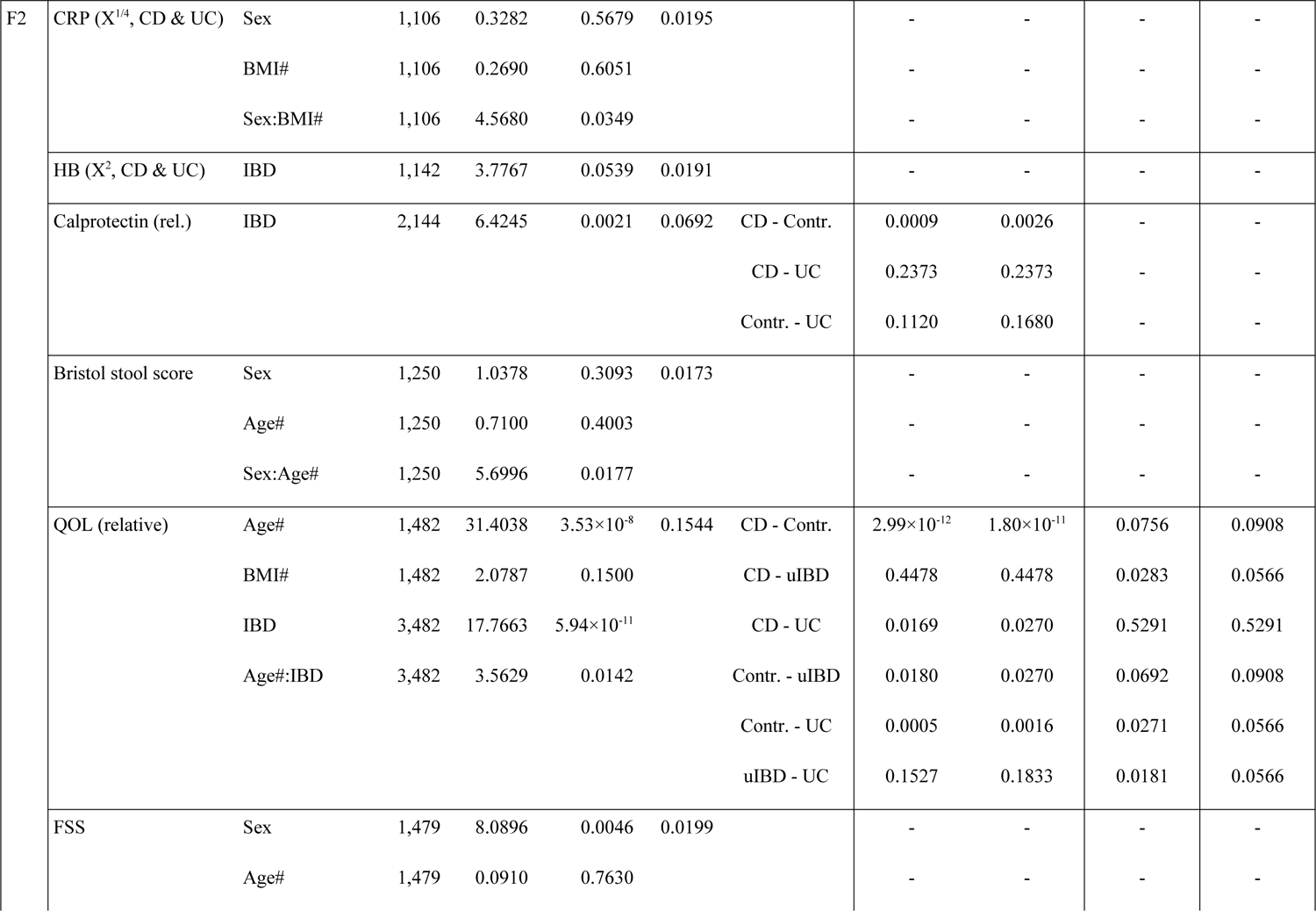
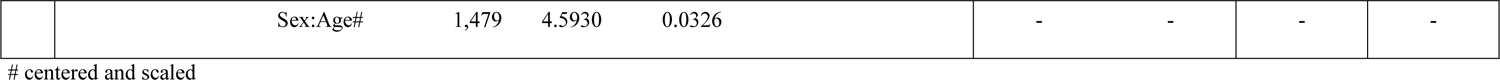
Analysis of major physiological characteristics across time points using linear models. Results depicted are models after variable selection minimizing AIC.

|  |  |  |  |  |  |  |  | Mean |  | Slope |  |
| --- | --- | --- | --- | --- | --- | --- | --- | --- | --- | --- | --- |
| TP | Trait | Model Factors | *DF* | *F* | *P* | *adj R2* | Comparison | *P* | *P(FDR)* | *P* | *P(FDR)* |
| BL | ASCA IgA | BMI# | 1,750 | 3.5792 | 0.0589 | 0.1835 | CD - Contr. | 7.50×10<sup>-33</sup> | 4.50×10<sup>-32</sup> | - | - |
|  | (log) | IBD | 3,750 | 56.6106 | 2.00×10<sup>-16</sup> |  | CD - uIBD | 7.06×10<sup>-6</sup> | 1.41×10<sup>-5</sup> | - | - |
|  |  |  |  |  |  |  | CD - UC | 1.54×10<sup>-18</sup> | 4.62×10<sup>-18</sup> | - | - |

|  |  |  |  |  |  |  |  |  |  |  |
| --- | --- | --- | --- | --- | --- | --- | --- | --- | --- | --- |
|  |  |  |  |  |  | Contr. - uIBD | 0.6907 | 0.6907 | - | - |
|  |  |  |  |  |  | Contr. - UC | 0.3482 | 0.5223 | - | - |
|  |  |  |  |  |  | uIBD - UC | 0.4621 | 0.5545 | - | - |
| ASCA IgG | Age# | 1,775 | 5.5172 | 0.0191 | 0.1684 | CD - Contr. | 3.26×10<sup>-28</sup> | 1.95×10<sup>-27</sup> | - | - |
| (log) | IBD | 3,775 | 52.0840 | 2.00×10<sup>-16</sup> |  | CD - uIBD | 2.62×10<sup>-6</sup> | 5.25×10<sup>-6</sup> | - | - |
|  |  |  |  |  |  | CD - UC | 5.15×10<sup>-21</sup> | 1.55×10<sup>-20</sup> | - | - |
|  |  |  |  |  |  | Contr. - uIBD | 0.3032 | 0.4548 | - | - |
|  |  |  |  |  |  | Contr. - UC | 0.4175 | 0.4871 | - | - |
|  |  |  |  |  |  | uIBD - UC | 0.4871 | 0.4871 | - | - |
| GP2 IgA | Sex | 1,744 | 0.0869 | 0.7682 | 0.0905 | CD - Contr. | 6.98×10<sup>-14</sup> | 4.19×10<sup>-13</sup> | 0.5056 | 0.7584 |
| (log) | Age# | 1,744 | 4.3479 | 0.0374 |  | CD - uIBD | 0.0808 | 0.1616 | 0.0099 | 0.0198 |
|  | BMI# | 1,744 | 0.4423 | 0.5062 |  | CD - UC | 9.09×10<sup>-7</sup> | 2.73×10<sup>-6</sup> | 0.8492 | 0.8492 |
|  | IBD | 3,744 | 20.5139 | 8.85×10<sup>-13</sup> |  | Contr. - uIBD | 0.4227 | 0.5073 | 0.0040 | 0.0198 |
|  | Sex:Age# | 1,744 | 10.1477 | 0.0015 |  | Contr. - UC | 0.2299 | 0.3449 | 0.7278 | 0.8492 |
|  | BMI#:IBD | 3,744 | 2.8203 | 0.0381 |  | uIBD - UC | 0.7453 | 0.7453 | 0.0092 | 0.0198 |
| GP2 IgG | Sex | 1,770 | 1.3630 | 0.2434 | 0.0480 | CD - Contr. | 2.84×10<sup>-8</sup> | 1.71×10<sup>-7</sup> | - | - |
| (log) | Age# | 1,770 | 0.3528 | 0.5527 |  | CD - uIBD | 0.1108 | 0.2216 | - | - |
|  | IBD | 3,770 | 11.3671 | 2.67×10<sup>-7</sup> |  | CD - UC | 0.0001 | 0.0003 | - | - |
|  | Sex:Age# | 1,770 | 9.3248 | 0.0023 |  | Contr. - uIBD | 0.8118 | 0.9654 | - | - |
|  |  |  |  |  |  | Contr. - UC | 0.6145 | 0.9218 | - | - |
|  |  |  |  |  |  | uIBD - UC | 0.9654 | 0.9654 | - | - |
| CRP ($X^{1/4}$) | Age# | 1,295 | 0.2114 | 0.6460 | 0.0323 | | - | - | - | - |
| (only CD & UC) | BMI# | 1,295 | 9.7144 | 0.0020 |  |  | - | - | - | - |
|  | IBD | 1,295 | 0.0050 | 0.9439 |  |  | - | - | - | - |
|  | Age#:IBD | 1,295 | 4.0463 | 0.0452 |  |  | - | - | - | - |
| HB ($X^2$) | Sex | 1,414 | 123.8936 | $2.36 \times 10^{-25}$ | 0.2408 | | - | - | - | - |
| (CD & UC) | BMI# | 1,414 | 10.0727 | 0.0016 |  |  | - | - | - | - |
| Calprotectin (rel.) | Age# | 1,1755 | 6.5053 | 0.0108 | 0.0576 | CD - Contr. | $7.58 \times 10^{-21}$ | $4.55 \times 10^{-20}$ | 0.0091 | 0.0547 |
| | IBD | 3,1755 | 33.5156 | $6.19 \times 10^{-22}$ | | CD - uIBD | 0.4827 | 0.5792 | 0.9471 | 0.9471 |
|  | Age#:IBD | 3,1755 | 2.5284 | 0.0558 |  | CD - UC | 0.0240 | 0.0431 | 0.4795 | 0.7690 |
|  |  |  |  |  |  | Contr. - uIBD | 0.0287 | 0.0431 | 0.5127 | 0.7690 |
| | | | | | | Contr. - UC | $2.88 \times 10^{-10}$ | $8.64 \times 10^{-10}$ | 0.1388 | 0.4164 |
|  |  |  |  |  |  | uIBD - UC | 0.9287 | 0.9287 | 0.8561 | 0.9471 |
| Bristol stool score | Age# | 1,874 | 33.4743 | $1.01 \times 10^{-8}$ | 0.1825 | CD - Contr. | $1.50 \times 10^{-31}$ | $8.97 \times 10^{-31}$ | 0.0013 | 0.0076 |
| | IBD | 3,874 | 53.2308 | $2.20 \times 10^{-16}$ | | CD - uIBD | 0.0682 | 0.0861 | 0.6533 | 0.9478 |
|  | Age#:IBD | 3,874 | 3.4956 | 0.0152 |  | CD - UC | 0.0031 | 0.0063 | 0.0772 | 0.2315 |
|  |  |  |  |  |  |  | Contr. - uIBD | 0.0718 | 0.0861 | 0.7898 | 0.9478 |
|  |  |  |  |  |  |  | Contr. - UC | 1.94×10<sup>-18</sup> | 5.81×10<sup>-18</sup> | 0.2316 | 0.4632 |
|  |  |  |  |  |  |  | uIBD - UC | 0.3532 | 0.3532 | 0.9902 | 0.9902 |
| F1 | CRP (X<sup>1/4</sup>, CD & UC) | Sex | 1,175 | 2.2744 | 0.1333 | 0.0072 |  | - | - | - | - |
|  | HB (X<sup>2</sup>, CD & UC) | Sex | 1,197 | 38.7355 | 2.86×10<sup>-9</sup> | 0.1601 |  | - | - | - | - |
|  | Calprotectin (rel.) | Age# | 1,507 | 3.7867 | 0.0522 | 0.0921 | CD - Contr. | 4.54×10<sup>-9</sup> | 2.73×10<sup>-8</sup> | 0.0194 | 0.0583 |
|  |  | IBD | 3,507 | 14.4469 | 4.81×10<sup>-9</sup> |  | CD - uIBD | 0.6170 | 0.7404 | 0.0860 | 0.1290 |
|  |  | Age#:IBD | 3,507 | 4.0004 | 0.0078 |  | CD - UC | 0.5625 | 0.7404 | 0.5011 | 0.5011 |
|  |  |  |  |  |  |  | Contr. - uIBD | 0.2880 | 0.5760 | 0.2935 | 0.3521 |
|  |  |  |  |  |  |  | Contr. - UC | 2.91×10<sup>-5</sup> | 0.0001 | 0.0102 | 0.0583 |
|  |  |  |  |  |  |  | uIBD - UC | 0.7708 | 0.7708 | 0.0508 | 0.1015 |
|  | Bristol stool score | Sex | 1,314 | 4.2960 | 0.0390 | 0.0104 |  | - | - | - | - |
|  | QOL (relative) | Sex | 1,593 | 5.1103 | 0.0242 | 0.1588 | CD - Contr. | 8.28×10<sup>-18</sup> | 4.97×10<sup>-17</sup> | - | - |
|  |  | Age# | 1,593 | 23.4394 | 1.65×10<sup>-6</sup> |  | CD - uIBD | 0.1088 | 0.1632 | - | - |
|  |  | BMI# | 1,593 | 5.4685 | 0.0197 |  | CD - UC | 0.0016 | 0.0031 | - | - |
|  |  | IBD | 3,593 | 26.7214 | 3.20×10<sup>-16</sup> |  | Contr. - uIBD | 0.5548 | 0.5684 | - | - |
|  |  | Sex:BMI# | 1,593 | 6.0564 | 0.0141 |  | Contr. - UC | 0.0001 | 0.0002 | - | - |
|  |  |  |  |  |  |  | uIBD - UC | 0.5684 | 0.5684 | - | - |
| F2 | CRP (X<sup>1/4</sup>, CD & UC) | Sex | 1,106 | 0.3282 | 0.5679 | 0.0195 |  | - | - | - | - |
|  |  | BMI# | 1,106 | 0.2690 | 0.6051 |  |  | - | - | - | - |
|  |  | Sex:BMI# | 1,106 | 4.5680 | 0.0349 |  |  | - | - | - | - |
|  | HB (X<sup>2</sup>, CD & UC) | IBD | 1,142 | 3.7767 | 0.0539 | 0.0191 |  | - | - | - | - |
|  | Calprotectin (rel.) | IBD | 2,144 | 6.4245 | 0.0021 | 0.0692 | CD - Contr. | 0.0009 | 0.0026 | - | - |
|  |  |  |  |  |  |  | CD - UC | 0.2373 | 0.2373 | - | - |
|  |  |  |  |  |  |  | Contr. - UC | 0.1120 | 0.1680 | - | - |
|  | Bristol stool score | Sex | 1,250 | 1.0378 | 0.3093 | 0.0173 |  | - | - | - | - |
|  |  | Age# | 1,250 | 0.7100 | 0.4003 |  |  | - | - | - | - |
|  |  | Sex:Age# | 1,250 | 5.6996 | 0.0177 |  |  | - | - | - | - |
|  | QOL (relative) | Age# | 1,482 | 31.4038 | 3.53×10<sup>-8</sup> | 0.1544 | CD - Contr. | 2.99×10<sup>-12</sup> | 1.80×10<sup>-11</sup> | 0.0756 | 0.0908 |
|  |  | BMI# | 1,482 | 2.0787 | 0.1500 |  | CD - uIBD | 0.4478 | 0.4478 | 0.0283 | 0.0566 |
|  |  | IBD | 3,482 | 17.7663 | 5.94×10<sup>-11</sup> |  | CD - UC | 0.0169 | 0.0270 | 0.5291 | 0.5291 |
|  |  | Age#:IBD | 3,482 | 3.5629 | 0.0142 |  | Contr. - uIBD | 0.0180 | 0.0270 | 0.0692 | 0.0908 |
|  |  |  |  |  |  |  | Contr. - UC | 0.0005 | 0.0016 | 0.0271 | 0.0566 |
|  |  |  |  |  |  |  | uIBD - UC | 0.1527 | 0.1833 | 0.0181 | 0.0566 |
|  | FSS | Sex | 1,479 | 8.0896 | 0.0046 | 0.0199 |  | - | - | - | - |
|  |  | Age# | 1,479 | 0.0910 | 0.7630 |  |  | - | - | - | - |

centered and scaled
|  |  |  |  |  |  |  |  |  |
| --- | --- | --- | --- | --- | --- | --- | --- | --- |
|  | Sex:Age# | 1,479 | 4.5930 | 0.0326 | - | - | - | - |

### Genetic predisposition to IBD can be expressed by polygenic risk scores (PRS) and is associated to physiological signs of inflammation

Due to the increasing number of variants associated to IBD it has become possible to quantify an individual’s genetic risk or genetic predisposition to CD, UC, or IBD using Polygenic Risk Scores (PRS) [36]. On average CD- and UC-PRS were relatively disease specific as they only showed significant differences from healthy KINDRED participants in the respective patient subset, with the exception of the more general IBD-PRS which was clearly increased in CD and UC patients alike, compared to healthy control individuals (Figure 1C). Interestingly, the average UC-PRS and general IBD-PRS were significantly elevated among future onset cases, while the CD-PRS showed no clear pattern. In addition to the average higher genetic risk present in future onset cases, the probability of IBD onset was significantly associated with UC-PRS and IBD-PRS, even though only nominally statistically significant (see Table S6, Figure S6A). The genetic risk for IBD should naturally be increased in IBD patients, compared to healthy individuals in general, a pattern displayed by all three PRS variants. However, closely related healthy first-degree relatives (FDRs) showed no significantly elevated PRS. The average genetic risk for IBD did not significantly differ among healthy FDRs, more distant relatives (>1st degree relatives), and unrelated controls (Figure S7). This pattern points to a significantly stronger accumulation of risk variants in affected individuals and onset patients, even in comparison to closely related family members and a high disease specificity of the respective PRS except for the more general IBD-PRS.

In our analyses, we tested PRS-CD, PRS-UC, and PRS-IBD for associations with different biomarkers. We detected a significant positive relationship only between CD- and IBD-PRS and the levels of ASCA-IgA/IgG and, to some extent, between IBD-PRS and the levels of anti-GP2-IgA (Figure 1D, Figure S5, Table S5). However, other inflammatory markers, such as calprotectin, BSS, CRP, and Hb did not show any, or only weak associations with genetic risk scores for IBD (Figure 1D, Figure S5, Table S5).

### Differential abundance of microbial taxa across IBD pathologies and time

Among the three time points we investigated, the fecal communities were dominated mainly by members of the phylum Bacteroidetes, followed by members of Firmicutes and Proteobacteria, and minor taxonomic groups (*e.g.* Fusobacteria, Tenericutes, Candidatus Saccharibacteria; Figure 2A, Figure S8). At this high taxonomic level strong differences between the main IBD pathologies are detectable, as reflected in the association of Bacteriodetes and Firmicutes with the control individuals and the associations of Proteobacteria, Fusobacteria and Cand. Saccharibacteria with IBD, particularly CD (Figure S6, Table S7). These associations are consistent over time and carry down to the ASV level (see below).

**Figure 2:**
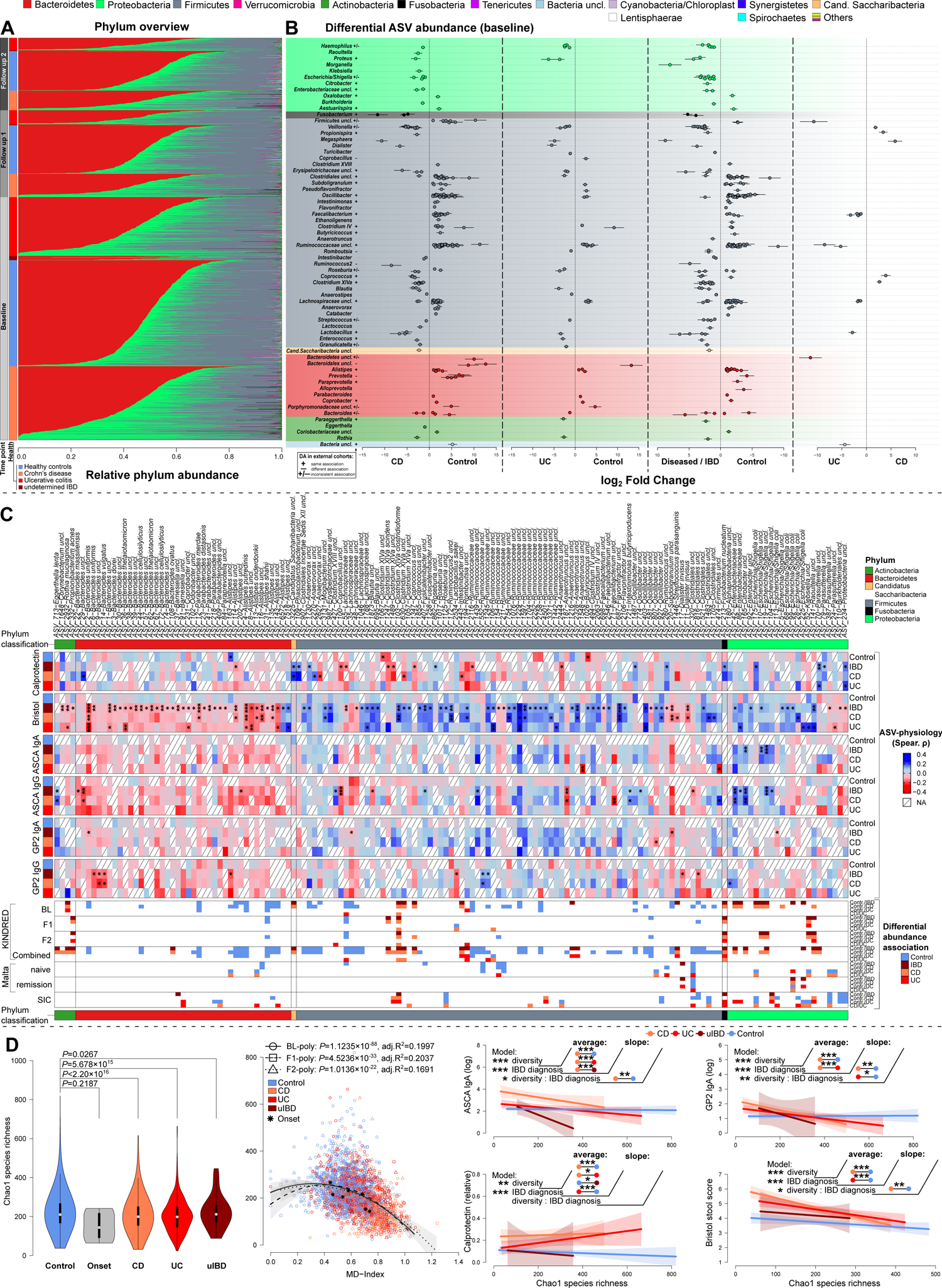
Taxon differential abundances and predictability. **(A)** Overview of individual phylum abundances across time points and health conditions in the KINDRED Cohort. **(B)** Differential abundance analyses of ASVs based on the sample time point (BL). Displayed are the log fold changes for each ASV clustered by genus classification, including standard errors of the fold changes as indicated by individual error bars. Color indicates the phylum membership of the ASVs/Genera. DA only displays significant differential abundance for the respective comparison/contrast (*P_FDR_*≤0.05). **(C)** Partial correlation of CLR transformed taxon abundances with core physiological measures via *ppcor* [143], combining the *P* values of Spearman-, Kendall-, and Pearson correlations via Brown’s method and corrected via FDR [144]. Correlations were adjusted for age, gender, and BMI and Spearman *ρ* is used to visualize correlation strength between taxa and clinical measures (# *P_FDR_*≤0.1000, * *P_FDR_*≤0.0500, ** *P_FDR_*≤0.0100, *** *P_FDR_*≤0.0010). Overlapping patterns of differential abundance for the respective taxa in the KINDRED cohort, Maltese-, and Swedish cohort is indicated in the bottom color bars. **(D)** Differences in species richness between healthy individuals, future onset cases, CD, UC, and uIBD cases (Wilcoxon test). Correlation of species richness with the microbial dysbiosis index (MD-index), which results in a negative, but non-linear relationship (optimal AIC based fit) between diversity and dysbiosis (BL (poly): *F*_2,1809_=226.97, *P*<2.2×10^-16^, *adj.R^2^*=0.1997; F1 (poly): *F*_2,645_=83.778, *P*<2.2×10^-16^, *adj.R^2^*=0.2037; F2 (poly): *F*_2,536_=55.745, *P*<2.2×10^-16^, *adj.R^2^*=0.1691; LM). Also the relationship between alpha diversity and several IBD relevant clinical measures, shows significant relationships between alpha diversity and host physiology, in a disease condition specific manner (see Table S15, Figure S11). Pairwise differences with respect to average differences, or differences in slope are highlighted in the plots (# *P*≤0.1000, * *P*≤0.0500, ** *P*≤0.0100, *** *P*≤0.0010).

By investigating abundance differences at the ASV level, we detected more than 750 significant differentially abundant taxa/ASVs among the main IBD pathologies (BL: 294, F1: 150, F2: 193, combined: 536; Figure 2B). At the baseline time point we detected taxa belonging to the *Enterobacteriaceae* (Gammaproteobacteria), such as *Haemophilus, Escherichia/Shigella, Raoultella,* and *Klebsiella,* which were strongly overabundant in IBD patients (Figure 2C). Particularly in the fecal community of CD patients we observed a high abundance of *Klebsiella* and other *Enterobacteriaceae,* which persisted over time (see Figure 2C, S9, S10). In the phylum Firmicutes we found various associations with different IBD pathologies. Members of *Veillonella, Megasphaera, Dialister, Enterococcus,* and *Granulicatella* were more abundant in CD and UC patients alike. In contrast, members of the *Ruminococcaceae* (e.g. *Clostridium IV, Oscillibacter, Faecalibacterium*) are consistently overrepresented in healthy individuals. *Erysipelotrichaceae* is more abundant in IBD patients and is frequently associated with UC. Members of the *Bacteroidetes*, with the exception of some *Bacteroides sp.* are consistently more abundant in healthy individuals, *e.g. Alistipes, Prevotella, Alloprevotella, Parabacteroides*, and *Bacteroides* (see Figure 2C, S9, S10).

Interestingly IBD-associated taxa belonging to *Veillonella, Dialister* and *Granulicatella* (Firmicutes), but also *Haemophilus, Klebsiella* (Proteobacteria), and *Rothia* (Actinobacteria) are prominent indicators of oral colonization of the intestinal microbiome. Similarly, a rarely described disease associated taxon, *Cand. Saccharibacteria* [37] are commonly found in the oral microbiome. *Fusobacteria* (*i.e. F. nucleatum*), which are generally associated with the development of colorectal cancers, were found to be overabundant in CD patients and could also be replicated in external cohorts (see Table S8-S11). The majority of associations, such as the associations of *Veillonella*, or members of *Clostridium XIVa clostridioforme*), *Clostridium XVIII ramosum*), and *Clostridium sensu stricto* groups with IBD, or the higher abundances of ASVs belonging to *Alistipes* and *uncl. Clostridiales* in healthy individuals, can be consistently detected in external cohorts, although some incongruities exist between cohorts (*e.g. Oscillibacter* ASVs in KINDRED-Contr./SIC-IBD patients).

Various taxa showed significant correlations with clinical indicators of intestinal inflammation and genetic risk scores. Indicators of oralization correlate with physiological signs of inflammation (e.g. *R. mucilaginosa-*BSS ↓, *Cand. Saccharibacter-*calprotectin ↑, *Klebsiella uncl.-* BSS ↑, *F. nucleatum*-calprotectin ↑, *Dialister invisus-*BSS↓ & anti-GP2 IgA/IgG ↓, *Veillonella uncl.-*IBD-PRS ↑). Interestingly, most Firmicutes correlated very consistently and almost entirely positively with the Bristol stool score (BSS) under IBD conditions (IBD, CD, UC), while members of the Bacteroidetes correlated very consistently negatively with BSS, encompassing pathobionts and commonly probiotic taxa (see Figure 2C, Table S12). This pattern was also present in healthy individuals, but did not reach significance. When we focused on the association of bacteria with genetic risk for IBD, we found that most Bacteroidetes (e.g. *Alistipes*) were correlated with decreasing genetic risk for CD, UC, and IBD, with the exception of *Bacteroides fragillis* (Figure S11, Table S13). Interestingly, *Barnesiella uncl.,* a common butyrate producer, correlated positively with genetic CD risk in healthy individuals, but correlated negatively with genetic CD risk in diseased individuals. In general, correlations with clinical parameters and genetic risk were most frequent in IBD patients, especially in CD patients, while almost no significant correlations were detected in UC patients. Different IBD associated Proteobacteria, specifically members of *Enterobacteriaceae* are strongly associated with increased ASCA IgG/IgA levels and calprotectin levels, as well as decreased gut transit time (increased BSS; *e.g. ASV_99 Escherichia/Shigella coli*) only in IBD/CD patients, similar to the findings for the IBD/CD associated Firmicutes *Clostridium XlVa clostridioforme* (ASV-103). Moreover, the *Enterobacteriaceae (e.g.* ASV-42-*Enterobacteriaceae uncl.,* ASV-165*-Citrobacter uncl.)*, as well as members of the *Clostridium XlVa* strongly correlated with increasing genetic risk for IBD, and more precisely CD in the patient cohorts (Figure 2C & Figure S11; Table S12). Several Firmicutes SCFA producers correlate with decreasing ASCA IgG/IgA levels and IBD risk in healthy and diseased individuals (e.g. ASV_71 *Faecalibacterium uncl.*), while the abundances of others correlate with elevated levels of genetic risk, particularly in diseased individuals (*e.g. Blautia, Ruminococcacea uncl.*). Overall, we observed consistent associations of ASVs to genetic risk for IBD and different clinical markers for IBD, with only minor associations in UC patients or with genetic risk for UC.

### Alpha diversity patterns associated with IBD pathology, clinical characteristics, and microbial dysbiosis

A critical characteristic of ecosystems is their complexity, which can be informative for assessing the community state, productivity or stability and can be based on, e.g. the number of community members, their distribution, or their phylogenetic relationships. Overall, we identified an extensive loss of species diversity in IBD patients with respect to species richness (Figure 2D), complexity or phylogenetic relatedness of community members (NTI<0, NRI<0) (Figure S12, Table 3). Individuals suffering from CD show a significantly greater loss of diversity than UC patients based on richness and complexity but not at the phylogenetic level. Onset cases only showed a slight but insignificant reduction in diversity (less richness, less complexity, less phylogenetic dispersion).

**Table 3:** Analysis of alpha diversity patterns across time points, correcting for age, sex, and BMI.

|  |  |  |  |  |  |  | Pairwise |  |
| --- | --- | --- | --- | --- | --- | --- | --- | --- |
|  | Alpha diversity | Model | *DF* | *F* | *P* | *adj. R2* | Contrast | P (adjusted) |
| BL | Shannon<br>(effective) | Age# | 1,1727 | 4.1607 | 0.0415 | 0.1214 | **CD-Contr.** | **4.95×10<sup>-12</sup>** |
|  |  | BMI# | 1,1727 | 0.0108 | 0.9174 |  | **CD-IBD** | **0.0483** |
|  |  | Sex | 1,1727 | 2.0196 | 0.1555 |  | **CD-UC** | **5.16×10<sup>-6</sup>** |
|  |  | **IBD diagnosis** | 3,1727 | **79.7519** | **2.56×10<sup>-48</sup>** |  | Contr.-IBD | 0.1270 |
|  |  |  |  |  |  |  | **Contr.-UC** | **4.99×10<sup>-12</sup>** |
|  |  |  |  |  |  |  | IBD-UC | 0.8411 |
|  | Chao | Age# | 1,1727 | 4.1801 | 0.0411 | 0.1189 | **CD-Contr.** | **4.95×10<sup>-12</sup>** |
|  |  | BMI# | 1,1727 | 0.0440 | 0.8338 |  | CD-IBD | 0.0921 |
|  |  | Sex | 1,1727 | 1.3698 | 0.2420 |  | **CD-UC** | **4.18×10<sup>-7</sup>** |
|  |  | **IBD diagnosis** | 3,1727 | **78.0602** | **2.37×10<sup>-47</sup>** |  | Contr.-IBD | 0.0723 |
|  |  |  |  |  |  |  | **Contr.-UC** | **4.99×10<sup>-12</sup>** |
|  |  |  |  |  |  |  | IBD-UC | 0.9774 |
|  | unweighted<br>NRI | Age# | 1,1727 | 21.7176 | 0.0000 | 0.0146 | CD-Contr. | 0.1364 |
|  |  | BMI# | 1,1727 | 0.1274 | 0.7212 |  | CD-IBD | 0.7220 |
|  |  | Sex | 1,1727 | 0.7269 | 0.3940 |  | CD-UC | 0.9318 |
|  |  | **IBD diagnosis** | 3,1727 | **3.0502** | **0.0276** |  | Contr.-IBD | 0.9823 |
|  |  |  |  |  |  |  | **Contr.-UC** | **0.0393** |
|  |  |  |  |  |  |  | IBD-UC | 0.5945 |
|  | unweighted<br>NTI | Age# | 1,1727 | 18.6248 | 0.0000 | 0.0555 | **CD-Contr.** | **6.52×10<sup>-12</sup>** |
|  |  | BMI# | 1,1727 | 8.1830 | 0.0043 |  | CD-IBD | 0.9981 |
|  |  | Sex | 1,1727 | 12.2254 | 0.0005 |  | CD-UC | 0.9177 |
|  |  | **IBD diagnosis** | 3,1727 | **22.9073** | **1.53×10<sup>-14</sup>** |  | Contr.-IBD | 0.0588 |
|  |  |  |  |  |  |  | **Contr.-UC** | **3.27×10<sup>-9</sup>** |
|  |  |  |  |  |  |  | IBD-UC | 0.9783 |
| F1 | Shannon<br>(effective) | Age# | 1,599 | 3.7799 | 0.0523 | 0.1384 | **CD-Contr.** | **4.54×10<sup>-10</sup>** |
|  |  | BMI# | 1,599 | 0.2665 | 0.6059 |  | CD-IBD | 0.7611 |
|  |  | Sex | 1,599 | 3.8384 | 0.0506 |  | **CD-UC** | **0.0026** |
|  |  | **IBD diagnosis** | 3,599 | **31.7658** | **4.58×10<sup>-19</sup>** |  | Contr.-IBD | 0.4993 |
|  |  |  |  |  |  |  | **Contr.-UC** | **0.0002** |
|  |  |  |  |  |  |  | IBD-UC | 0.9988 |
|  | Chao | Age# | 1,599 | 3.3731 | 0.0668 | 0.1091 | **CD-Contr.** | **4.54×10<sup>-10</sup>** |
|  |  | BMI# | 1,599 | 0.8586 | 0.3545 |  | CD-IBD | 0.8398 |
|  |  | Sex | 1,599 | 0.3035 | 0.5819 |  | **CD-UC** | **0.0081** |
|  |  | **IBD diagnosis** | 3,599 | **25.1887** | **2.33×10<sup>-15</sup>** |  | Contr.-IBD | 0.5692 |
|  |  |  |  |  |  |  | **Contr.-UC** | **0.0013** |
|  |  |  |  |  |  |  | IBD-UC | 0.9975 |
|  | unweighted<br>NRI | Age# | 1,599 | 2.3377 | 0.1268 | 0.0159 | **CD-Contr.** | **0.0231** |
|  |  | BMI# | 1,599 | 4.1609 | 0.0418 |  | CD-IBD | 0.9981 |
|  |  | Sex | 1,599 | 0.2207 | 0.6387 |  | CD-UC | 0.1840 |
|  |  | **IBD diagnosis** | 3,599 | **3.0230** | **0.0292** |  | Contr.-IBD | 0.8141 |
|  |  |  |  |  |  |  | Contr.-UC | 0.9979 |
|  |  |  |  |  |  |  | IBD-UC | 0.8492 |
|  | unweighted<br>NTI | Age# | 1,599 | 0.0932 | 0.7603 | 0.0657 | **CD-Contr.** | **1.71×10<sup>-7</sup>** |
|  |  | BMI# | 1,599 | 12.7048 | 0.0004 |  | CD-IBD | 0.7954 |
|  |  | Sex | 1,599 | 0.3129 | 0.5761 |  | **CD-UC** | **0.0060** |
|  |  | **IBD diagnosis** | 3,599 | **11.8031** | **1.61×10<sup>-7</sup>** |  | Contr.-IBD | 0.0948 |
|  |  |  |  |  |  |  | Contr.-UC | 0.6806 |
|  |  |  |  |  |  |  | IBD-UC | 0.2089 |
| F2 | Shannon | Age# | 1,483 | 2.1681 | 0.1415 | 0.1375 | **CD-Contr.** | **6.25×10<sup>-11</sup>** |
|  | (effective) | BMI# | 1,483 | 0.0739 | 0.7859 |  | CD-IBD | 0.5878 |
|  |  | Sex | 1,483 | 0.0128 | 0.9098 |  | **CD-UC** | **0.0080** |
|  |  | **IBD diagnosis** | 3,483 | **27.2317** | **2.75×10<sup>-16</sup>** |  | Contr.-IBD | 0.7581 |
|  |  |  |  |  |  |  | **Contr.-UC** | **0.0003** |
|  |  |  |  |  |  |  | IBD-UC | 0.9958 |
|  | Chao | Age# | 1,483 | 5.4159 | 0.0204 | 0.1281 | **CD-Contr.** | **6.25×10<sup>-11</sup>** |
|  |  | BMI# | 1,483 | 2.8540 | 0.0918 |  | CD-IBD | 0.7797 |
|  |  | Sex | 1,483 | 0.3818 | 0.5369 |  | **CD-UC** | **0.0010** |
|  |  | **IBD diagnosis** | 3,483 | **23.0590** | **5.77×10<sup>-14</sup>** |  | Contr.-IBD | 0.6626 |
|  |  |  |  |  |  |  | **Contr.-UC** | **0.0197** |
|  |  |  |  |  |  |  | IBD-UC | 0.9929 |
|  | unweighted<br>NRI | Age# | 1,483 | 3.5979 | 0.0584 | 0.0179 | CD-Contr. | 0.1956 |
|  |  | BMI# | 1,483 | 2.1588 | 0.1424 |  | CD-IBD | 0.5201 |
|  |  | Sex | 1,483 | 1.4772 | 0.2248 |  | CD-UC | 0.9997 |
|  |  | **IBD diagnosis** | 3,483 | **2.5548** | **0.0547** |  | Contr.-IBD | 0.2385 |
|  |  |  |  |  |  |  | Contr.-UC | 0.3879 |
|  |  |  |  |  |  |  | IBD-UC | 0.5123 |
|  | unweighted<br>NTI | Age# | 1,483 | 3.0392 | 0.0819 | 0.0231 | CD-Contr. | 0.0585 |
|  |  | BMI# | 1,483 | 3.0373 | 0.0820 |  | CD-IBD | 0.5655 |
|  |  | Sex | 1,483 | 0.0126 | 0.9105 |  | CD-UC | 0.9990 |
|  |  | **IBD diagnosis** | 3,483 | **3.8270** | **0.0099** |  | Contr.-IBD | 0.2120 |
|  |  |  |  |  |  |  | Contr.-UC | 0.1015 |
|  |  |  |  |  |  |  | IBD-UC | 0.6040 |

Based on the differential abundance patterns we derived two indices of host health based on the bacterial abundance and distribution, namely, the Microbial Dysbiosis index (MD-index) as introduced by Gevers *et al.* (2014) and the more generalized GMHI (general microbiome health index) as developed by Gupta *et al*. (2020) [38,39]. Focusing on the MD-index we can observe a clearly negative correlation of alpha diversity with community dysbiosis at any level, further implying an accelerated loss of taxon diversity with increasing dysbiosis (breakpoint at MD=0.615, Figure 2D, Table S13). The phylogenetic composition changes to become more random with increasing dysbiosis but shows a less severe pattern than species richness or abundance distribution (Figure S12, Table S13). Across all time points we can identified a trend of strongly decreasing diversity in UC and CD patients with increasing dysbiosis, while healthy controls showed a far slower decline of diversity under increasing dysbiosis, particularly in the taxon-based indices (Table S14). The correlations of different alpha diversity metrics with the GMHI showed comparable but weaker relationships as observed with the MD-index. Alpha diversity metrics are further negatively (Chao1, Shannon H) and positively (NRI, NTI) correlated with physiological signs of inflammation, such as ASCA-IgA/IgG, calprotectin, and BSS, particularly in IBD patients, which translates to more severe signs of inflammation at a lower community diversity in diseased individuals (CD, UC, uIBD; Figure 2G, Table S15).

### Community differences between IBD pathologies are associated with clinical-, anthropometric-characteristics, patterns of community dysbiosis, and genetic risk

By investigating community similarities at different levels, we revealed various patterns of health-associated, anthropometric, and lifestyle related community differences. In particular, IBD pathology showed a strong association with community differences, with the strongest separation between microbial communities originating from healthy individuals and those originating from CD patients (Figure 3A; Table S16). In this context, we observed a strong association of the taxonomic community composition with the MD-index and to a lesser extent with GMHI (Table S16). This gradient of dysbiosis parallels the community differentiation between healthy and diseased individuals, but also recapitulates significant gradients of physiological markers for inflammation such as BSS, ASCA IgA/IgG levels, and calprotectin levels in the communities (see Figure 3A, 3B, 3J; Figure S13, S14; Table S17 & Table S18). We also detected significant, yet weak correlations between taxonomic community composition and polygenic risk scores, which were very weak for UC associated genetic risk and strongest for CD and general IBD risk at the taxonomic-(Figure S7D) and functional levels (Figure S7E) and followed the direction of the dysbiosis gradient. Similarly, the cumulative genetic risk for CD and general IBD positively correlated with the MD-index, while UC-PRS displayed no significant correlation, similar to GMHI (Figure S5B & S5C).

**Figure 3:**
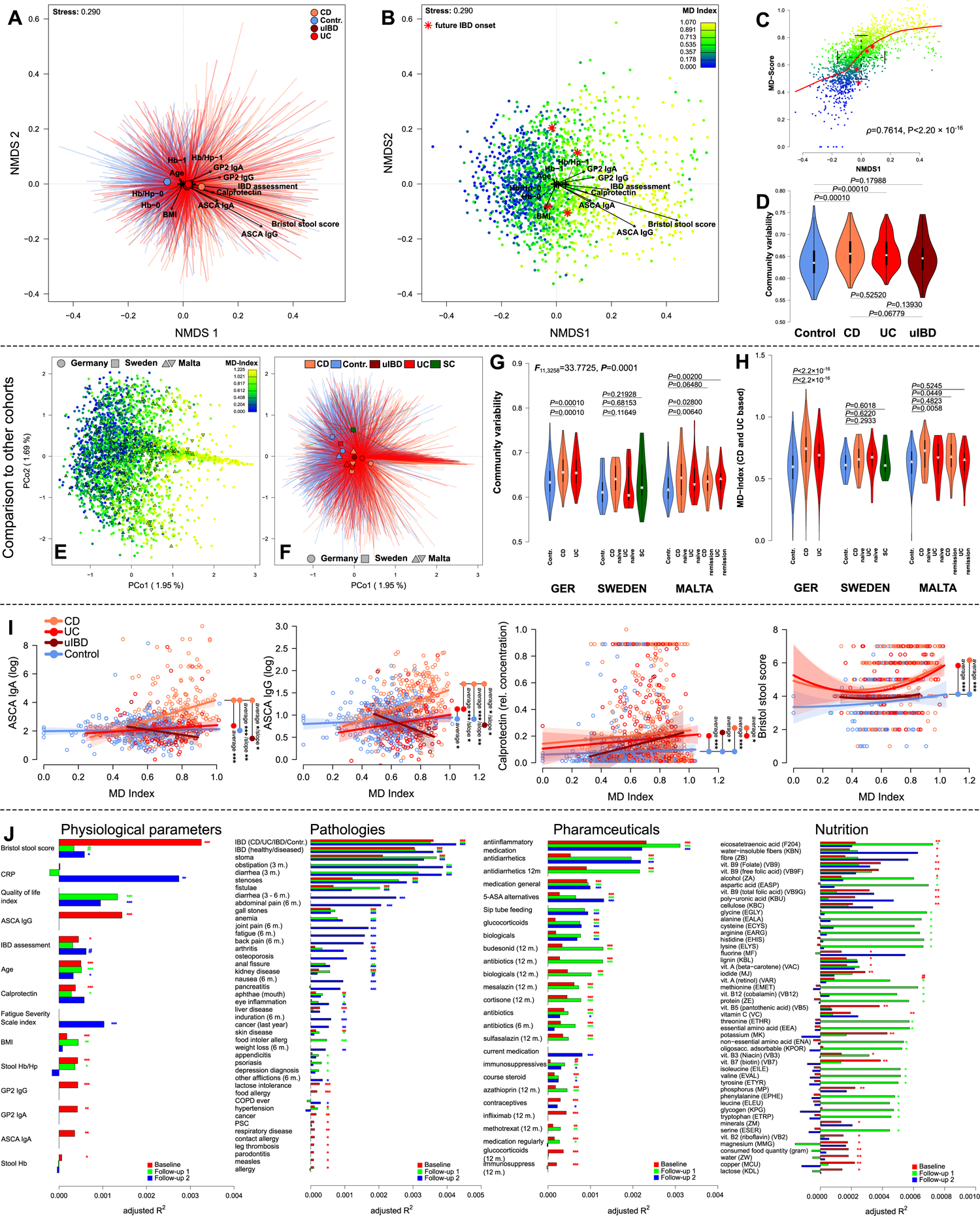
**(A)** Non-metric Multidimensional Scaling (NMDS) of Bray-Curtis distances among baseline samples, displaying the significant clustering by health conditions and significant correlations of clinical inflammation measures with community distance (BL, see Table S16 & Table S17). **(B)** NMDS displaying the gradient of community dysbiosis as expressed by MD-index [38], in parallel with clinical measures of inflammation and healthy onset cases highlighted in red (*, develop IBD in F1). **(C)** Correlation of MD-index and the first NMDS axis showing a clear gradient of dysbiosis in the community. Onset cases are distributed in the range of standard deviation around the mean of the community dsitribution (NMDS1) and the severity of dysbiosis (MD-index). **(D)** Community variability between health conditions as measured by the distance to the group centroid, is overall significantly different between health conditions (*F*_3,1808_=46.0315, *P*=0.00001, PERMANOVA), and significantly increased in CD and UC patients as compared to healthy controls (Table S19). **(E)** Principle coordinate analysis of German-, Swedish-, and Maltese samples, highlighting the transferrability of the dysbiosis gradient across cohorts (MD-index derived from german samples), **(F)** as well as a common disease wise clustering of communities irrespective of sample origin. **(G)** Community variability between health/IBD conditions within and between the German-, Swedish-, and Maltese cohorts showing a common theme of increased variability in IBD cases. **(H)** Mean differences of dysbiosis (MD-index derived from german samples) within and across cohorts, with the strongest differences between healthy and CD individuals. **(I)** Scatterplots visualize the correlation of selected physiological inflammation markers with the microbial dysbiosis score [38], and show disease specific differences as compared to healthy control individuals. Pairwise differences with respect to average differences, or differences in slope are highlighted in the plots (see Figure S16, Table S18; # *P*≤0.1000, * *P*≤0.0500, ** *P*≤0.0100, *** *P*≤0.0010). **(J)** Visualization of the explained variation of significant anthropometric variables as based on serial PERMANOVA of Bray-Curtis distances in all three time points available and focused on physiological measures (Table S17), different reported pathologies of individuals (Table S21), use of pharmaceuticals (Table S22), and nutrient intake as derived from FFQ data (Table S23). Variables are displayed if they show significant clustering in at least one time point (# *P_FDR_*≤0.1000, * *P_FDR_*≤0.0500, ** *P_FDR_*≤0.0100, *** *P_FDR_*≤0.0010).

Furthermore, onset cases were mostly distributed around the mean of the community compositional distribution and dysbiosis gradient, and thus did not represent community outliers (Figure 3C, Figure S13 & S14). The MD-index (and GMHI) strongly distinguished IBD and non-IBD individuals and corresponded with the clustering of microbial communities with respect to IBD pathology, even if applied to external patient cohorts (Sweden, Malta) (Figure 3E-3H, Figure S14G). The MD-index is most strongly elevated in CD patients and further distinguished CD patients (particularly treatment-naïve) from healthy controls in the Maltese cohort (Figure 3H). GMHI displayed on average a more consistent decrease in diseased individuals across all cohorts, and further distinguished UC patients from healthy controls (Figure S14H). These “universal” disease patterns are also reflected in the sample clustering by IBD condition, irrespective of cohort origin (conditional dbRDA (conditioned by geographic origin): *F*_4,3308_=4.0406, *P*<0.0001, *adj. R^2^*=0.0037) and through the significantly elevated taxonomic and functional community variability in IBD patients compared to healthy controls within (Figure 3D, Figure S14H & S15, Table S20) and across cohorts (Figure 3G & S14H).

Both dysbiosis scores corresponded closely with several physiological signs of inflammation, such as the reactivity to fungal antigens as expressed via ASCA IgG or IgA titers, or related immunological markers, like anti-GP2 IgG and IgA levels (Figure 3I & 3J, Figure S16, Table S18). ASCA antibody levels increased most strongly in CD patients with increasing MD score (decreasing GMHI). In contrast, anti-GP2 IgA/IgG levels positively correlate with MD but appear to show a slight overall increase and higher average MD levels in CD and UC patients, as expected. Overall, the relationship of GMHI and MD-index with ASCA or anti-GP2 antibody levels were comparable (Table S18). The GMHI and MD-index displayed strong associations with the BSS, with lower stool consistency in IBD patients and increasing dysbiosis (MD ↑, GMHI ↓) (Figure 3I). The MD-index was strongly associated with increasing BMI and calprotectin levels, which were on average higher in CD and UC patients than in healthy controls (Figure 3I, Figure 1B). The concentrations of serum CRP- and Hb levels, which were only assessed in diseased individuals, were not significantly associated with microbial dysbiosis metrics (Figure S16). The practitioner’s overall assessment of disease severity in IBD patients only (1-no/4-severe inflammation), revealed a strong positive association with MD, while GMHI was not significantly correlated.

When we investigated dysbiosis patterns among FDRs, we observed that the MD and GMHI in affected individuals were greater or lower, respectively, than those in healthy family members. However, healthy FDRs of IBD patients, generally related, or unrelated healthy individuals, did not show a greater degree of dysbiosis. However, the MD-index was weakly associated with an increased probability of IBD onset (DF=1,1735, *Z*=-2.0685, *P(Z)*=0.0386, N_onset_=7/1815; DF=1,726, *Z*=-1.2579, *P(Z)*=0.2084, N_onset_=7/794; binomial GLM), while the GMHI showed no significant association with IBD onset.

IBD pathologies were consistently the most influential health conditions for the taxonomic and functional differences of the microbial communities (Figure 3J & Figure 4D, Table S21). The most pronounced differences were between the healthy controls and CD patients, followed by the differences between healthy controls and UC patients, while functional and taxonomic differences between UC- and CD patients persist (see Table S16, Table S21). Together with the presence of a stoma/artificial bowel outlet (colostomy, ileostomy, permanent/temporary stoma), other pathologies that influence the anatomic structure or gut transition time, such as obstipation, diarrhea, and intestinal stenosis strongly influence the taxonomic and functional composition of the microbial communities (Figure 3J). Thus, chronic and acute diseases, which influence passage time, have a significant impact on the taxonomic and functional composition of the intestinal microbiome. This pattern remained consistent even after excluding potential confounding effects such as age, BMI, sex, and IBD pathology (see Table S21).

**Figure 4:**
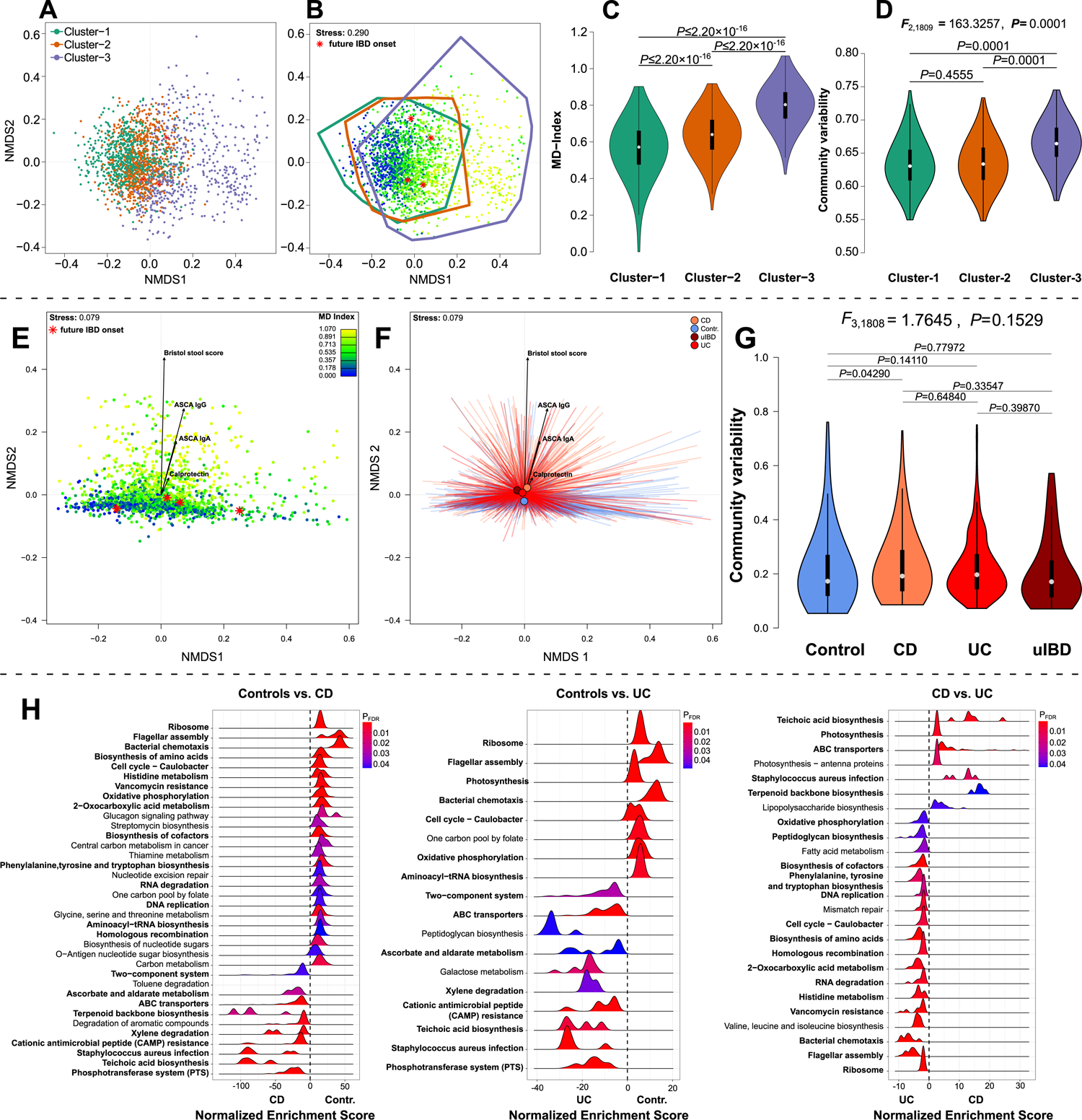
**(A)** Community clusters of the microbial community at the baseline time point, determined by Dirichilet Multinomial Mixture modelling (DMM) and optimal clustering was determined via Laplace goodness of fit optimization. **(B)** Overlay of the Microbial Dysbiosis score gradient (Gevers *et al.*, 2014) and community clusters (outlines), including healthy onset patients at baseline (indicated by **\***). **(C)** Community clusters display a significantly elevated level of dysbiosis in clusters 2 and 3 (pairwise Wilcoxon tests), **(D)** as well as elevated levels of community variability in cluster-3 (PERMANOVA). **(E)** Non-metric Multidimensional Scaling (NMDS) of Bray-Curtis distances of PICRUSt2 based KO abundances among baseline samples (Douglas *et al.*, 2020), displaying the gradient of community dysbiosis as expressed by the taxonomy based MD-index (Gevers *et al.*, 2014), **(F)** and displaying the significant clustering by health conditions and significant correlations of clinical inflammation measures with community distance (see Table S16 & Table S17). Healthy IBD onset patients are highlighted in red (*, patients who developed IBD by the next follow-up). **(G)** Community variability between health conditions as measured by the distance to the group centroid, was significantly increased in CD and UC patients as compared to healthy controls (PERMANOVA, Table S19). **(H)** Density plots show the significant Gene Set Enrichments [142] of differentially abundant PICRUSt2 based KOs, between controls, CD patients, and UC patients (*P_FDR_*≤0.05). Differential abundance was tested via *DESeq2* and the enrichment score was derived from the -log10(P-values)*direction of fold change (Table S23). Repeatedly detected metabolic pathways are highlighted in bold.

In addition to the impact of different pathologies, most importantly IBD, we observed a significant impact of various pharmaceutical treatments on microbial communities. Interestingly, antibiotic treatments between 6 weeks and one year prior had significant but not prominent effects as reported by [40]. However, the most influential were drugs that modify the fecal passage time (*e.g.* Loperamide, surfactants), further pronouncing the strong and consistent association of BSS with community composition. In particular, these pharmaceuticals remained influential after correcting for IBD condition, age, BMI, and sex (Figure 3J, Table S22) and associated with higher levels of BSS (*W*=24756, *P*=3.3910×10^-9^, Wilcoxon-test).

Investigating the impact of macro and micronutrient intake on the microbiota revealed a very interesting parallel distribution of F20:4 (Eicosatetraenoic acid/Arachidonic acid) and MD/GMHI indices with the microbial community composition (Figure 3J, Figure S13G-I), which was the only nutrient whose intake associated with an increasing dysbiosis signal in the microbial community, in direct correlation (Figures S14J; Table S23), and consistent in follow-up time points (see Figure 3J & S13G-I; Table S23). This eicosanoid precursor can be a potent, immunologically active molecule [41] and its uptake at baseline was weakly correlated to MD (*ρ*=0.0486, *P*=0.0772, Spearman correlation), BMI (*ρ*=0.1278, *P*=4.7168×10^-6^), ASCA IgG (*ρ*=0.0647, *P*=0.0881), and calprotectin (*ρ*=0.0514, *P*=0.0648).

Increasingly dysbiotic communities were further correlated to increasing consumption of glycogen, various amino acids and general protein uptake (ZE), which can be interpreted as elevated uptake of animal products (Figures S14J), which paralleled the gradients of inflammatory markers (*e.g.* calprotectin; see Figures S13D-F & Figures S13G-I). In contrast, the increased uptake of various vitamins and trace minerals, as well as long-chain carbohydrates (e.g. ZB-Fiber) significantly coincided with a less inflammatory/disturbed community structure (GMHI and MD) and were associated with reduced physiological signs of inflammation, potentially via SCFA production [42]. The functional community composition was only associated with vitamin B derivatives and trace mineral consumption (see Figure 4D & Figure S17G-I; Table S23).

### Uncharacteristic community clusters (DMM) are associated with dysbiosis and clinical signs of inflammation

Unsupervised community clustering revealed three community clusters, which do not fully correspond to previous reports [43], but share common indicators reported in the literature (Figure 4A). Cluster-1 is characterized by a high abundance of *Prevotella* and *Ruminococcaceae* (*Prevotella*: *P_FDR_*=1.1162×10^-26^; *Ruminococcus*: *P_FDR_*=2.1846×10^-59^; Kruskal test), while cluster-2 shows a high abundance of *e.g. Bacteroides* and *Faecalibacterium* (*Bacteroides*: *P_FDR_*=1.4604×10^-17^; *Faecalibacterium*: *P_FDR_*=1.5310×10^-84^; Kruskal test). In contrast, individuals in Cluster-3 showed a greater propensity for inflammation as indicated by the elevated MD-index (*P*<2.2×10^-16^; Kruskal test, Figure 4B & 4C) and community variability (Figure 4D), as well as increased levels of physiological inflammation markers such as ASCA-IgG/IgA, and increased stool moistness/softness (Figure S18A). This cluster further showed a higher abundance of *Enterobacteriaceae* (*e.g. Escherichia/Shigella*: *P_FDR_*=4.0936×10^-61^; Kruskal test), which is unconventional for so-called “enterotypes”. However, this community type has been reported before and appears to correspond to a dysbiotic community state, positioned outside the standard community states reported sofar [44–46]. In addition, we detected a significantly greater level of genetic risk for CD (*F*_2,1665_=12.5869, *P*=3.7541×10^-6^; LM) and general IBD (*F*_2,1665_=4.3156, *P*=0.0135; LM) among individuals in this unconventional community cluster, but not for UC risk (*F*_2,1665_=0.1712, *P*=0.8427; Figure S18B), indicating the strong effect of IBD and a broad genetic disease risk on community composition.

### Imputed community functions reveal the importance of amino acid related pathways and bacterial surface structures in health and disease

On average, communities differ more strongly between IBD pathologies based on the imputed functions compared to taxonomic differences, particularly regarding the difference between healthy controls and CD patients (Table S16). The ranking of single associations of pathologies, physiological parameters, and pharmaceutical treatments with functional community differences overlaps with the taxon-based analyses, although fewer associations can be detected (Figure 4E & 4F, Figure S17, Table S17, S21, S22, S23). Additionally, physiological signs of inflammation, such as ASCA IgA/IgG or decreasing stool consistency parallel the dysbiosis gradient in the functional composition (Figure 4E, Figure S17, Figure S17D; Table S16, S17). The MD-index and GMHI explained a greater percentage of community variation at the functional level (see Figure 4A, Figure S17M; Table S16), while functional community differences were only weakly correlated with the genetic risk scores for CD (*P*<0.05; Figure S6E).

At the level of single functions, we detected various differentially abundant KEGG Orthologous Groups (BL: N_total_=5071, F1: N_total_=4347; F2: N_total_=4402). By investigating systematic changes in functional abundances via Gene Set Enrichment Analysis [47], we identified various highly persistent functional differences between disease states, which were further supported in the external cohorts (Table S24). Various metabolic pathways involved in the metabolism and biosynthesis of amino acids, ranging from general *Biosynthesis of amino acids*, to *Phenylalanine, tyrosine and tryptophan biosynthesis,* among others, are consistently enriched in healthy individuals or rarely in UC patients compared to CD patients. Other amino acid-related pathways were similarly enriched in healthy individuals (*Arginine biosynthesis; Alanine, aspartate and glutamate metabolism; Valine, leucine and isoleucine biosynthesis; Lysine biosynthesis*; see Figure 4E; Table S24). Basic metabolic functions and cell growth were more strongly represented in healthy controls and were repeated among the other cohorts in a similar direction (*e.g. Citrate cycle (TCA cycle), Oxidative phosphorylation*; Figure 4H, Figure S19-S22; Table S24), as were pathways involved in bacterial motility and biofilm formation, which were enriched in healthy individuals (*Bacterial chemotaxis, Flagellar assembly, Biofilm formation - Pseudomonas aeruginosa*). Oxidative phosphorylation also entails anaerobic electron transfer reactions (anaerobic respiration) that reduce other electron acceptors than oxygen for ATP generation (*e.g.* nitrate, sulfate, fumarate), as performed by various intestinal bacteria (*e.g. Desulfovibrio*). In contrast, functions related to carbohydrate degradation (*e.g. ABC transporters, Phosphotransferase system (PTS), Fructose and mannose metabolism, Galactose metabolism*) were very consistently enriched in IBD-associated communities and may, in combination with changing SCFA levels (F1 & F2: *Propanoate metabolism and Butanoate metabolism*), contribute to mucosal degradation. Bacterial traits that are central to the generation and integration of immune modulatory factors of gram-negative (*Lipopolysaccharide biosynthesis*) and gram-positive bacteria (*Teichoic acid biosynthesis, Peptidoglycan biosynthesis*) were also consistently enriched among IBD patients (Figure S19-S22; Table S24).

### Network analyses show systemic and transferrable patterns of community disturbance, and high disease dependent taxon centrality in community networks

To analyze the microbial community as a whole, we constructed co-abundance networks to identify important taxa and patterns in the fecal microbial communities that may be central for community and host homeostasis (Figure 5A). Across networks (split by time point, or by time point and pathology), we observed significant differences in the composition and topography at the network level between IBD pathologies and healthy community networks in the KINDRED cohort (control *vs*. IBD: *F*_1,7_=2.4873, *P*=0.0947, *R^2^*=0.2622, *adj. R^2^*=0.1568; see Figure 5B),. This also held true when external IBD cohorts were included (Control *vs*. IBD: *F*_1,12_=2.1301, *P*=0.0604, *R^2^*=0.1507, *adj. R^2^*=0.0800, Control *vs*. IBD (incl. general IBD networks): *F*_1,18_=2.4144, *P*=0.0412, *R^2^*=0.1183, *adj. R^2^*=0.0693, Graphlet distance; Figure 5C & Figure S27A). Furthermore, we observed signs of network/community contraction in IBD communities in the KINDRED cohort, as implied by the increasing network density and radius in diseased community networks, which remained consistent when we included networks derived from healthy and diseased external cohorts (Figure S27B). In addition, “natural connectivity” is increased in IBD communities, which strongly suggests a greater resistance to disturbances or stochastic changes by those communities. A higher “natural connectivity” after disease manifestation implies more densely connected networks with fewer bottlenecks, and pronounced community structure, which tend to possess a greater robustness against random fluctuations and community decay [48,49].

**Figure 5:**
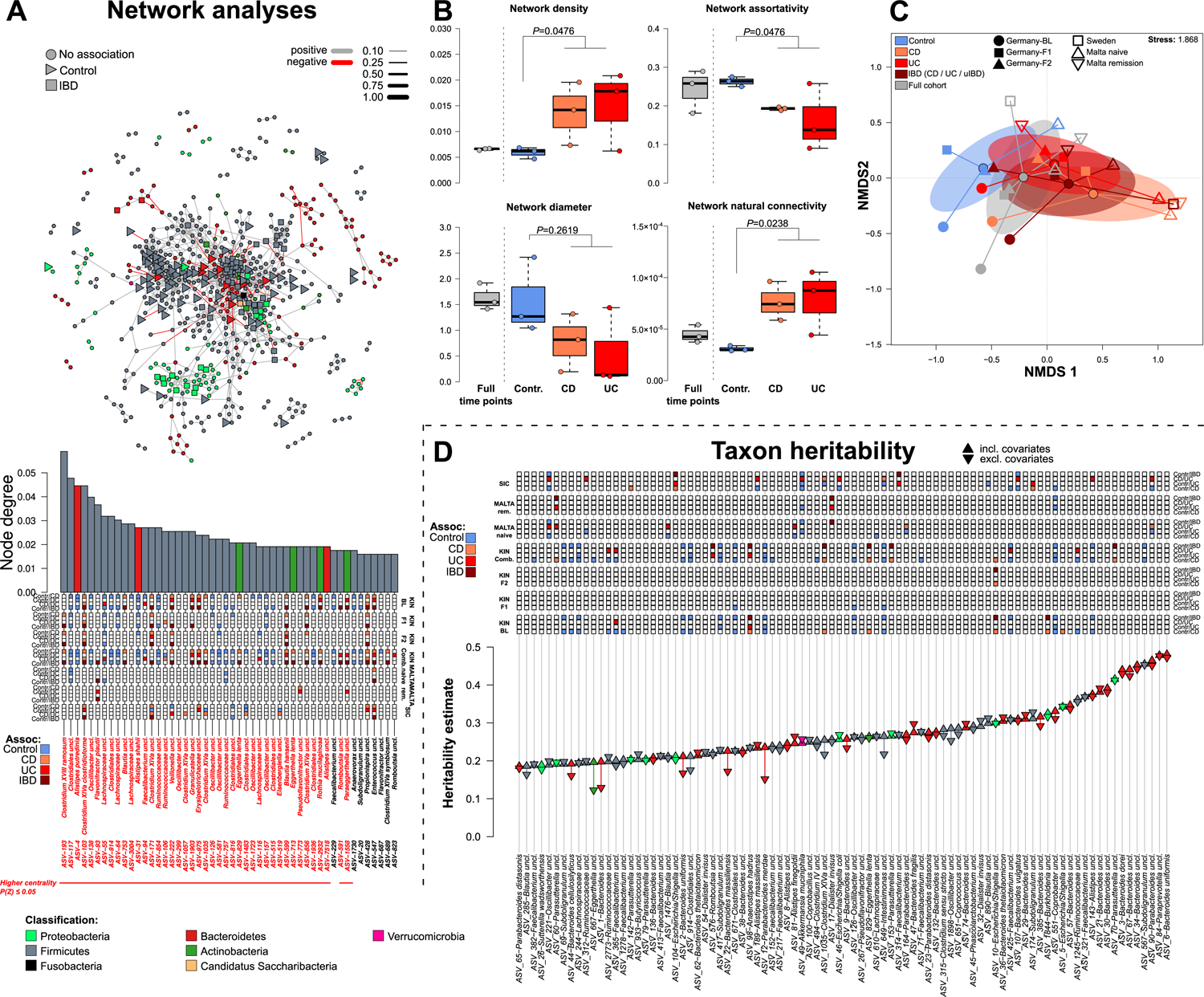
**(A)** Spiec-Easi networks of baseline samples (N=1812). Bacterial nodes highlight significant differentially abundant ASVs in the network [157]. Bacteria not showing any differential abundance patterns between IBD patients and healthy controls are signified via (●), bacteria overabundant in IBD (combined CD, UC, uIBD) via (▪) and bacteria more abudnant in controls are signified via (►). The barplot visualizes node centrality based on the number of connections (node degree) at the baseline time point. Colored boxes highlight corresponding ASV differential abundance patterns in KINDRED, external cohorts, and their association to IBD onset and remission. Significance of centralities is derived from Z-tests against a randomized networks (10’000). **(B)** Global network characteristics were derived from the baseline and disease specific sub-networks, which can be informative for stability and structure of the respective networks (Wilcoxon-test healthy vs. CD/UC). Network metrics include centrality based assortativity [150], network diameter, radius, and size as well as density/clustering [151,152], and natural connectivity [48]. **(C)** Network similarity of disease and time point specific sub networks, as well as subnetworks of external cohorts (Malta, Sweden) based on graphlet distance [153] and displayed via NMDS. Networks show a clear compositional difference between healthy and diseased networks (Control *vs.* IBD (incl. IBD networks): *F*_1,18_=2.4144, *P*=0.0412, *R^2^*=0.1183, *adj. R^2^*=0.0693; PERMANOVA). **(D)** Heritability estimates derived from the likelihood based method *lme4qtl* (Ziyatdinov *et al.*, 2018) using either only kinship information with or without additional environmental and anthropometric covariates (▴-incl. environmental covariates, ▾-no covariates). Only the upper quartile of taxa are highlighted (based on *h^2^* estimate including environmental covariates). Additional information like differential abundance in IBD accross cohorts are depicted for each taxon (Figure S28A, Table S26).

At the single taxon level/node level (Figure 5A, Figure S23-S26), we observed various pathogenic and IBD associated groups, consisting mainly of *Clostridium XVIII ramosum* (*Erysipelotrichaceae*: ASV_193) and *Clostridium XlVa clostridioforme/spec.* (e.g. ASV_103, ASV_171), as well as various members of the oral community (e.g. *Veillonella, Rothia, Cand. Saccharibacteria, Fusobacterium)* and *Enterobacteriaceae* (*e.g. Klebsiella*), to be consistently central across time points (particularly in diseased networks) and characteristically abundant in IBD patients. Similarly *Flavonifractor plautii* (ASV_93, *Ruminococcaceae*) or the Actinobacterium *Eggerthella lenta* (ASV 713), are elevated as well in IBD, highly central, and known for pro-inflammatory characteristics [50–54] (Figure S23-S26, Table S25). In contrast, ASVs belonging to *Alistipes* (*i.e. A. putredinis*, *Alistipes spec.*, *A. shahii*) are highly central and consistently abundant in control individuals across time points and external cohorts, like other SCFA producing bacteria (e.g. *Porphyromonadaceae: Barnesiella, Odoribacter, Parabacteroides spec*.; *Lachnospiracea: Clostridium XlVb uncl*.; *Ruminococcacea* (*Clostridial cluster IV*, *Ruminococcus;* Figure 5A, Figure S23-S26, Table S25). However, through the nature of network construction through statistical means, centralities, and key-stone status may vary [55,56].

### Heritability of microbial taxa

Using a mixed model framework, we investigated the heritability of single ASVs using pedigree relationships in the cohort. Most of these associations were driven not only by host kinship or family conditions, but also by environmental/ anthropometric/ lifestyle characteristics (age, sex, BMI, and IBD status). At the ASV level, the taxa showing the highest heritability estimates were mainly members of the *Bacteroides*, while particularly *Bacteroides uniformis* (ASV-6, *h^2^* =0.4723) and *Paraprevotella uncl.* (ASV 84, *h^2^* =0.4783) had the highest heritability estimates (Table S26). Most taxa were impacted by environmental influences in addition to the apparent kinship effect/familiar transmission (*h^2^ < h^2^;* Figure 5D, Figure S28A; Table S26). Interestingly, potential probiotic bacteria abundant in healthy individuals, or mutualistic taxa (not associated to IBD) displayed high heritability (*e.g. Parabacteroides uncl., Feacalibacteria uncl., A. muciniphila, B. thetaiotaomicron, Alistipes uncl.*). *Akkermansia muciniphila* displayed a lower impact of environmental/anthropogenic variables (*h^2^* > *h^2^*) on the inheritance patterns, and displayed a better fit solely by relatedness (*h^2^* =0.2573, Figure 5D, Figure S28A), similar to other highly heritable taxa, such as the ubiquitous human gut commensal *Bacteroides uniformis* [57] (Table S26). However, among the top 25% percent of the most heritable ASVs, we observed disease-associated taxa, mainly belonging to the Proteobacteria such as e.g. *Escherichia/Shigella* (ASV-5, ASV-10) or the typical oral *Dialister invisus* (ASV-17, Firmicutes), which were more abundant in IBD patients. The community wide dysbiosis indices (MD-index and GMHI), showed relatively strong heritability as well, which implies a genetic influence or intra-family transmission of dysbiosis. GMHI was within the boundaries of ASV based estimates in our cohort (*h^2^* =0.1401), while the heritability of the MD-index was clearly lower (*h^2^* =0.0813) and distinctly lower than estimates reported by Turpin *et al.* 2016 (*h^2^*=0.2728) for the MD-index or the heritability estimates for IBD itself (CD: 0.25-0.42, UC: 0.04-0.15) [58,59]. Overall, heritability of the communities appears to be disturbed in cases of IBD, as we observed a reduced correlation of community composition and genetic distance (kinship, IBS distance) in IBD cases compared to healthy individuals (Figure S28B), although overall correlations were still significant (kinship-*r*=0.4406, m^2^12=0.8059, *P*=0.0009, IBS-*r*=0.4272, m^2^12=0.8175, *P*=0.0009; Procrustes test).

## DISCUSSION

In this cohort study we investigated the state and evolution of IBD by combining taxonomic and functional microbial data from several successive time points with extensive anthropometric, medical, nutritional, and social information. Genome-wide association studies focusing on IBD revealed various genetic risk loci for IBD, which may contribute to immune dysregulation, antimicrobial immune reactions, and changes in host-microbiome homeostasis [60,17]. Various studies have investigated the interplay between microbes, metabolites, and host characteristics. These analyses revealed distinct physiological and microbial manifestations of the different IBD pathologies, which are reflected in external disease cohorts. The unsupervised community clustering (DMM community cluster 3) and significant differences of IBD patients across cohorts, highlight the severity of community turnover during IBD [46] and a strong signal of dysbiosis which may even warrant one or multiple distinct community states. Similarly, the strong differentiation of IBD-specific networks from healthy ones points to large changes in community dynamics after disease manifestation. These changes potentially result in a resilient or resistant community state as implied by the more connected, denser, and robust networks after IBD manifestation (radius, density, natural connectivity) even across cohorts [49]. The consistently observed increased community variability in diseased individuals is a probable result of community transitions. This phenomenon, the so called “Anna Karenina pattern”, describes a pattern in which microbial communities of dysbiotic/diseased individuals are more different from each other than communities of healthy individuals are among each other. This phenomenon may be attributed to a loss of regulation or strongly individualized stochastic effects on community composition after IBD development [61] and has been detected in other IBD studies [25,62–64]. In summary, different community characteristics, such as increasing interindividual variability, greater community density/connectivity, and large compositional changes after IBD manifestation, point toward large and to some extend universal community transformations in IBD development.

### Physiological and genetic markers of inflammation are strongly disease specific, associated to the microbiome, and associate to disease onset

The two main IBD pathologies, CD and UC, are physiologically, anatomically, and microbiologically quite distinct. We identified disease specific patterns in several physiological markers of IBD. In particular, anti-fungal ASCA IgG and IgA have strong associations to CD and IBD in general. ASCA Ig levels were not reported to be associated to high risk genetic markers of CD (CARD15/NOD2), but that its effect sizes might be genetically determined [65]. Thus, ASCA levels themselves are not genetically determined, but rather their responsiveness, which reflects the strngth of inflammatory reactions to environmental antigens, potentially also of bacterial origin. In contrast we showed a significant association with polygenic risk scores for IBD, particularly CD, which was not yet shown before (Lee et al., 2021), and might originate from the wider array of cumulative genetic risk factors known since the investigation by Halfvarson et al., 2005. This strong relationship of the PRS was not present for other physiological IBD biomarkers in this study.

Similar to ASCA Ig levels, anti-glycoprotein 2 antibodies (GP2) have been described as strong biomarkers for IBD [33,35], while fecal calprotectin represents a biomarker of general intestinal inflammation. All displayed strong disease specific differences and are strongly correlated with bacterial community characteristics such as composition, diversity, dysbiosis indeces, but far weaker associated to the genetic risk scores for IBD. Bacterial taxa that associated to ASCA levels overlap only in parts with a recent study in UC patients at the family level (*e.g. Ruminococcaceae*) [66].

Decreased stool consistency, an indicator of reduced intestinal transit time, elevated fecal water content and eventually diarrhea, are hallmarks of IBD pathology. Although not associated with genetic risk for IBD, stool consistency decreases dramatically with increasing dysbiosis, diversity, and community composition. Recently Vandeputte et al. showed the effects of fecal passage time on community patterns in a Belgian cohort, surpassing dietary and disease related signals [67,68]. We found similar patterns, with increasing passage time (lower BSS) associated with increasing populations of Bacteroidetes and decreasing abundances of Firmicutes members. These patterns have mostly a similar orientation, but are significantly exaggerated in IBD patients, which implies a greater impact of bacteria on passage time/stool consistency or a greater impact of passage time on the bacterial community under intestinal inflammation. In line with the centrality of stool consistency and transit time for community composition, we observed a strong gradient of community composition and IBD associated dysbiosis with decreasing transit time. Similarly, among the most influential pharmaceutical treatments in the cohort are anti-diarrhea drugs, and the most influential medical conditions in addition to IBD are conditions affecting fecal flow (stoma, obstipation, diarrhea, stenoses, etc.). Changes in transit time can change the taxonomic and functional composition-, bidirectionally interact with the microbiota, and alter the metabolite composition by transitioning from saccharolytic to proteolytic metabolism [69,70].

We determined a clear connection between the genetic risk of IBD development, particularly CD and general IBD, and the microbial community composition or level of dysbiosis. This finding emphasizes the potential connection between genetic risk factors such as *e.g. FUT2* or *NOD2* [60,71] and their combined influence on the microbial community, potentially leading to dysbiotic community dynamics in interaction with environmental factors. Polygenic risk scores integrate the abundance and penetrance of known risk variants for diseases such as IBD into a single genome-wide risk to develop the disease. Although PRS effectively capture established IBD variants, these scores may not account for yet unidentified genetic factors and were mainly derived from cohorts of predominantly European ancestry, thus missing out population specific variants [24]. However, in this cohort genetic risk for general IBD and CD showed stable and repeated connections to microbial- and physiological indicators of IBD and were derived and applied in a central European cohort. The relatively weak association of UC-PRS with microbial community characteristics, in contrast points to a reduced importance of the host-microbe interactions and/or lower density and penetrance of associated risk variants in UC [72]. In alignment with this, UC patients displayed comparatively weak and fewer associations with the microbiome, as seen before [64]. However, the associations that were detected are mostly consistent with other studies [66,73]. Naturally, PRS for IBDs are higher in IBD cases than in controls, however the clear and significantly elevated risk levels and the weak, albeit significant, increase in the probability of developing IBD with a higher PRS emphazises the potential of these scores to stratify individuals at risk and underscores the close interaction between genetic risk and potential environmental triggers such as the microbiome in IBD development [17]. *Barnesiella sp.,* a common mucosa-associated butyrate producer, showed a positive association with CD risk in healthy individuals but was negatively associated with CD risk in diseased individuals. A recent study revealed that this taxon is associated with a TLR4 modulating variant (*biliverdin reductase A, BLVRA*), which increases the risk for CD, but may dampen inflammation in a diseased state through SCFA production [74–76]. In contrast, we detected a reversal of this pattern in the commonly “probiotic” *Bacteroides fragillis* [77], which displayed a positive association with CD-PRS in IBD and CD patients. PRS also encompass important IBD-associated genes/risk variants for autophagy (e.g. *ATG16L1*, *NOD2*), which are unable to sense the anti-inflammatory signals of this taxon and may thus result in a proinflammatory cytokine bias through this taxon [78].

Broad genetic patterns, as determined by kinship, influenced the taxonomic and functional composition of the bacterial communities [79]. Social distance as well as broad relatedness have been described in detail to allow for bacterial transfer [80,81] and the transfer of potential phenotypes. Thus, horizontal and vertical intrafamily transmission [80], may add to the continuation and transfer of inflammation and dysbiosis as experimentally trackable in mouse experiments [82,83]. Interestingly the ubiquitous human gut commensal *Bacteroides uniformis* [57] among other *Bacteroides* species shows a strong association with host genetic variation as has been described elsewhere for humans and mice [84,76]. Although mainly commensals, which are associated to long chain carbohydrate degradation, this capability of *Bacteroides* may also drive pathogenic patterns in susceptible environments/hosts, due to the weakening of the mucosal barrier [85,86]. Thus, the varying genetic background of the host and strain/species specifc functional variation may explain the varying association of Bacteroides species to IBD, particularly in light of their relatively stable host-genetic associations [76]. However, in contrast to our study, the heritability of Bacteroidetes in general and *Bacteroides* in particular were shown to be low in twin studies [79], but were on a similar level with recent studies which used more complex kinships [87]. Another mucolytic species, *Akkermansia muciniphila,* was highly heritable in other studies, as well [79,87], and has often been associated with weight loss and metabolic health. Here we showed a clear association with non-IBD individuals, even though its role in IBD can be ambiguous [88,89]. The majority of heritable taxa belong to Firmicutes (149 ASVs *vs.* 96 ASVs other); however, highly heritable *Christensenellaceae* were not detected, potentially due to their low abundance/prevalence in the cohort [81,79,90]. Overall, the potential inheritance patterns we detected strongly resemble patterns of shared communities in mother-offspring comparisons, thus potentially highlighting the dominant matrilineal influence of these analyses [80]], although cohabitation could not be completely disentangled from family membership due to data restrictions. In addition we detected a general correspondence between differences of community composition and genetic distance, highlighting a community wide influence of genetics and heritability. This pattern appears to be disturbed among IBD patients, potentially due to the overwhelming effect of disease and related factors on the microbial communities which masks the broad genetic effects we detected.

### Oralization of the intestinal community and blooming of Enterobactericeae are common IBD characteristics across cohorts

A strong microbial pattern in IBD, particularly for CD, arises from the highly consistent association of oral bacteria with the disease. Various taxa across different phyla with a typical oral origin show clear associations with IBD (*Cand. Saccharibacteria, Rothia, Veillonella, Dialister, Fusobacteria, Klebsiella*) and highlighted a potential loss of colonization resistance of the community or detrimental physiological alterations. *Cand. Saccharibacteria* (formerly TM7) is a rarely described obligate bacterial epibiont involved in IBD-related microbial dysbiosis and oralization [37], but appeared very consistently in the investigated cohorts [64]. Additionally, *R. muciliginosa* is an oral taxon with a clear association with IBD, which interestingly closely associates with *Cand. Saccharibacteria* (Figure S29). It may thus represent a potential host for this taxon in the intestinal environment, as it until recently belonged to the same family as its oral hosts (*Actinomycetaceae*) [91]. Alternatively, this group may represent an independent TM7 group adapted to the intestinal environment, as observed in the ruminant TM7-G3 cluster [92] with its own host species. *F. nucleatum* and its cell components have been proposed as causal triggers of inflammation in e.g. rheumatoid arthritis [93] or even IBD [94], and have been implicated in the development of colorectal cancer [95]. Recent studies also displayed several overlapping, mostly oral bacteria associated to UC, such as *Veillonella dispar*, *Megasphaera micronuciformis*, *Haemophilus parainfluenzae*, and other *Enterobacteriaceae* [96] and a reduction of these ectopically colonizing oral bacteria can promote remission in IBD patients [66]. Furthermore, oralization of the microbiome and oral health itself are strongly associated to IBD development, risk, and severity [97,98].

However, the most consistent pattern of IBD association was observed for *Enterobacteriaceae,* such as potentially oral *Klebsiella sp.*, which are highly proinflammatory and often associated with UC and PSC, although we mainly detected stable associations with CD and IBD [97]. In addition, *Escherichia/Shigella* show clear and valid pathogenic or proinflammatory characteristics [38,99] and appear to characterize a distinct, potentially dysbiotic community type (DMM cluster-3), as previously proposed [44,45]. Various *Enterobacteriaceae*, are known to elicit strong immunological reactions and are associated with the development of IBD at various levels ranging from ecological (facilitation [100]) to metabolic [101] interactions that allow these taxa to benefit from inflammatory processes [102]. However, some members of the *Escherichia/Shigella* have been shown to be beneficial for the immunological development early in life [103] or carry beneficial characteristics [104]. Differentially abundant taxa, particularly oral taxa (*Fusobacterium, Dialister, Veillonella*) and the abundance patterns of *Enterobacteriaceae* correspond well with a recent large meta-analysis of IBD studies, as do abundances of potentially beneficial taxa (*e.g. Faecalibacterium, Ruminococcaceae, Alistipes*) [105].

### Amino acid metabolism and flagellar functions associate strongly with healthy individuals

Interestingly, one of the most enriched functions differing between healthy and diseased individuals is *Aminobenzoate Metabolism*. Anthranilate or 2-Aminobenzoic acid is a key metabolite of bacterial aromatic compound degradation, a precursor of tryptophan biosynthesis, and a key metabolite to reduce or even prevent IBD pathogenesis [106,107]. Additionally, other functions involved in amino acid- and tryptophan metabolism, *e.g. Phenylalanine, tyrosine and tryptophan biosynthesis*, were also enriched. Thus, aromatic amino acid biosynthesis appears to have a strong, and health relevant effect, as the enrichment of amino acid metabolism is very prominent in control subjects. Interestingly, bacterial functions involved in *Flagellar assembly* show a strong and consistent association with healthy subjects as well (antagonistic to IBD) and may arise from the abundance of so-called “silent” flagellins present in *Lachnospiraceae,* which have potential anti-inflammatory activity via TLR5 binding [108]. However, *Lachnospiraceae* derived flagellins display a strong immune reactivity in CD patients [109,110], which may represent an aberrant response driven by an already disturbed mucosal barrier, rather than a *de novo* reaction to members of the Lachnospiraceae [111,109]. *Clostridium clostridioforme* (*Clostridium XlVa*) is highly CD-specific and immunologically reactive according to our results and other studies [112,113]. A recent study identified a strong and cross cohort IBD association of *C. clostridioforme* with specific biosynthetic gene clusters, whose products lead to increased inflammation and mucosal barrier dysfunction [114]. *C. clostridioforme* strongly correlates with the concentration of the potentially pro-inflammatory long chained fatty acid eicosatetraenoic acid/arachidonic acid (F20:4) [113], and its uptake is positively associated to dysbiosis (*e.g.* long saturated: long unsaturated: lipoids) correlated with community composition and dysbiosis. Furthermore eicosatetraenoic acid positively associated with CD- and UC-associated taxa, *R. gnavus* but also *C. clostridioforme*, whereas typical probiotic taxa showed a significant negative relationship with this metabolite [113]. In addition to its immune- and inflammatory characteristics, this fatty acid possesses bactericidal activity and the potential to disrupt bacterial cell membranes [115]. This might indicate that IBD-associated taxa might be able to metabolize and synthesize F20:4 and are less susceptible to it, while it interferes with the growth health-associated species [113,114]. In contrast, the consumption of vitamins, long-chain carbohydrates, and long unsaturated fatty acids is correlated with lower levels of dysbiosis [42,116]. Bacterial traits that are central to the generation and integration of immune modulatory factors of gram-negative (*Lipopolysaccharide biosynthesis*) and gram-positive bacteria (*Teichoic acid biosynthesis, Peptidoglycan biosynthesis*) on the bacterial surface are also consistently enriched among IBD patients. Comparable patterns of these surface/capsular and membrane factors have also been observed in a recent meta-analysis of various IBD cohorts [117].

## CONCLUSION

The KINDRED cohort is a prospective cohort study that collects biomaterial and various types of data from IBD patients and their first- or second-degree relatives. The main aim of the KINDRED cohort is to identify lifestyle factors as well as serum and stool markers associated with the onset of the disease (in initially healthy relatives with a positive family history for IBD) and to investigate the individual disease course in patients on a multiomic level, integrating lifestyle, genetics, and microbial information. Using this multiomic dataset we identified strong and generalizable dysbiosis gradients, which correspond strongly with IBD pathologies, physiological manifestations of inflammation (*e.g.* BSS, calprotectin, anti-GP2 and ASCA IgA/IgG), genetic risk for IBD and general risk of disease onset. Patterns of overabundance and importance of various opportunistic pathogens (*e.g., Enterobacteriaceae, C. XIVa clostridioforme*), in addition to consistent patterns of oralization, characterize IBD patients. Functionally, pathways involved in amino acid metabolism and flagellar assembly are beneficial, while mucolytic functions are associated with IBD. Broad scale ecological patterns point totipping-point dynamics being involved in the drastic state transitions of communities into the chaotic communities characteristic for IBD (community variability, community differentiation). However, some limitations are present in this study. Healthy relatives were not examined in an examination center and all information about these individuals were obtained through self-administered questionnaires. Another potential weakness of the KINDRED cohort is its open design, where recruitment has run over a long period of time (from 2013, and ongoing), with a slow increase of onset cases, and steady decrease of participants with time due to lack of compliance or withdrawal from the study during the regular, yet only biennial follow-ups.

## MATERIALS AND METHODS

### Basic study design and cohort

The KINDRED cohort is a German-wide, prospective cohort study that collected both, questionnaire data and biomaterials from IBD patients and their (affected and unaffected) family members. Despite great efforts to include as many family members as possible, a focus has always been on unaffected individuals with a family history of IBD. The recruitment of study participants started in October 2013 and is still ongoing. Follow-up information and new biomaterial samples were collected prospectively at intervals of approximately two years (Figure S1-S3). As of April 2021, the IBD Family Cohort has thus enrolled 1497 IBD patients together with 1813 (initially) non-affected first- or second-degree relatives from Germany (minimum age at inclusion: 7 years). Participants, including the IBD patients, were asked to provide questionnaire data and biomaterials (blood, stool, hair) at baseline and after every 2 years of follow-up (Table S1 & S2). The resulting patient counts are shown in Table 1 and Figure 1A. In addition, physician-administered questionnaires were collected from IBD patients to obtain physician-validated information, such as diagnosis, disease pattern/location, activity, and medication (Table S1). Using these data, the Kiel IBD Family Cohort aims to facilitate the comprehensive molecular, clinical, lifestyle, nutritional, and sociodemographic characterization of patients and high-risk individuals, and to identify preclinical signs for the onset of IBD. Additional information on eligibility criteria, enrollment, data and biomaterial collection, as well as data management, and privacy protection is given in the Supplement of this article. A standard battery of questionnaires was used to assess self-reported dietary behavior (12 month recall questionnaire) and metrics of well-being and quality of life (quality of life index, Fatigue Severity Scale index), were also employed (see Supplement) [118–120]. Nutritional data were adjusted by total energy consumption to decouple caloric consumption from diet composition. Individuals with undefined or unclear IBD diagnosis but with intestinal inflammation (e.g. suspected CD/UC, colitis) are summarized under the category uIBD (undefined IBD, Table S3).

### Ethics and human samples

The KINDRED cohort study protocol was approved by the ethics committee of the Medical Faculty of Kiel University (AZ A117/13). Every study participant provided written informed consent on forms that were age-adapted. For participants under the age of 18 years, informed consent must also be signed by their parents. When the participants reached the ages of 12 and 18 years a new informed consent (reconsent) form was signed by the participants themselves and, in the case of 12-year-old adolescents, by their parents.

A cohort of treatment naïve and newly diagnosed IBD patients in an active disease state from Malta (naïve: N_CD_=31, N_UC_=25), including a healthy control cohort (N_Contr._=96), was recruited as described elsewhere [64]. In addition we included a Maltese patient cohort currently in disease remission and treatment (remission: N_CD_=32, N_UC_=66), as described recently [63].

Individuals from the Swedish Inception Cohort in IBD (SIC IBD) were included as treatment-naïve patients, between 20–77 years of age. Symptoms, such as diarrhea, abdominal pain, bloody or mucous stools for >2 weeks, were inclusion criteria. The final diagnosis of IBD was established according to internationally accepted criteria including clinical, microbiological, endoscopic, histological, and radiological evaluation (N_CD_=17, N_UC_=16). Patients with gastrointestinal symptoms but without endoscopic and histological signs of IBD-associated inflammation were considered symptomatic controls (SC, N_SC_=16). In addition, 17 healthy individuals were included as healthy controls (N_contr._=16, one failed sequencing).

### Physiological measurements

Calprotectin (indicator of intestinal inflammation) was measured in fecal samples using a Bühlmann fCAL™ ELISA kit (BÜHLMANN LABORATORIES AG) and analyzed using SoftMax Pro Software (Molecular Devices). ASCA IgA/IgG and anti-GP2 IgA/IgG were measured externally using ELISA (Medipan GmbH) including batchwise calibration samples. Occult fecal blood was determined by PreventID® Haemo/HaptOccult (Preventis GmbH, Bensheim, Germany). Other immunological measures (CRP Hb) were obtained during the initial examination using standard clinical tests.

### Polygenic Risk Scores

GSA data were quality controlled via gwas-qc (https://github.com/ikmb/gwas-qc) not correcting for closely related individuals in the cohort using hg19 (genome build 37) of the human genome and 1000 Genome reference set. Potential sample mix ups and unclear relatedness patterns were manually checked and corrected if needed. Imputation was performed after chromosome wise transformation into bgzip VCF files via plink2 [121]. Single VCF files were uploaded to hybridcomputing.ikmb.uni-kiel.de/webservice/sites/, imputed and phased via EagleImp (Genome build: GRCh37/hg19, Reference: 1000 Genomes Phase 3, r² filter: 0.1, allowed reference swaps, and strand flips) [122].

To examine the genetic susceptibility of individuals in the KINDRED cohort for IBD and its subtypes CD and UC we calculated Polygenic Risk Scores (PRS). GWAS summary statistics were taken from the meta-analysis of IBD from Liu et al [20]. The summary statistics are based on a total of 5956 CD and 6968 UC patients with an additional 21770 population of controls with European ancestry. PRS were calculated with *LDpred2* within the *bigsnpr* (v. 1.12.2) R package [123,36]. After quality control of the summary statistics [124] the method calculates a posterior mean effect size based on linkage disequilibrium information and base effect size for all remaining available markers in both data sets. We used the *auto*-method of *LDpred2,* which automatically estimated the parameter sparsity *p* and the SNP heritability *h^2^* and did not require to tune hyper-parameters in the validation set.

### Stool sample processing

DNA extraction, sequencing and bioinformatics processing of 16S rRNA gene libraries from stool samples were performed as described previously in detail [125].

### Data processing

Data processing of 16S sequences was performed using DADA2 1.10 [126] via the workflow for big datasets (https://benjjneb.github.io/dada2/bigdata.html, https://github.com/mruehlemann/ikmb_amplicon_processing/blob/master/dada2_16S_workow_with_AR.R) resulting in abundance tables of Amplicon Sequence Variants (ASVs). All sequencing runs were handled separately for error correction, read merging, and combined chimera detection. ASVs underwent taxonomic annotation using the naïve Bayesian classifier implemented in DADA2 using the Ribosomal Database Project 16 release [127,128]. ASV sequences were aligned via NAST-alignment to the SILVA core database and filtered for informative sites (constant gaps, constant bases) in mothur [129]. Phylogenetic tree construction on ASV alignment generated was carried out using FastTree 2.1 with the CAT substitution model with Γ-correction and improved accuracy, employing more minimum evolution rounds for initial tree search [-spr 4], more exhaustive tree search [-mlacc 2], and a slower initial tree search [-slownni] [130]. Unstratified KO categories of metagenome predictions were generated using the native ASV abundances and sequences in PICRUSt 2.5.0 using default workflows [131].

### Statistical methods

#### Alpha diversity

Species richness (Chao1), Simpson diversity (1-D), and phylogenetic alpha diversity (Nearest Taxon Index [NTI], Net Relatedness Index [NRI]) were calculated and analyzed in R 3.5.3.17-19. Phylogenetic measures of alpha diversity [Nearest Taxon Index (NTI), and Net Relatedness Index (NRI)] were derived using the *PhyloMeasures* package, based on 999 permutations against a null model preserving relative species richness within the communities [132–134] [NRI=-1 × (MPDobserved-mean (MPDrandom))/SD(MPDrandom); NTI=-1 × (MNTDobserved-mean(MNTDrandom))/SD(MNTDrandom)]. Both metrics are phylogenetic effect sizes, for which positive values indicate phylogenetic clustering, values close to zero indicate neutral or random community assembly, and negative values indicate phylogenetic overdispersion, either over the whole phylogenetic tree (NRI) or across the closest related species/tips of the phylogenetic tree (NTI). The relationships of physiological- and microbial inflammation markers/indices with alpha diversity were analyzed using linear models after correcting for relevant covariates (LM: residuals(variable∼gender+scaled BMI + scaled Age) ∼ IBD (Contr., CD, UC, IBD) * physiological variable). Model fits were visualized via *base* R and *jTools* [135]. General correlation of MD with alpha diversity was tested via simple linear models, segmented linear models (*segmented*), or polynomial linear models and assessed via the AIC.

#### Beta diversity

Analyses were conducted via distance based (conditional) Redundancy analyses and permutative ANOVA, as well as with a multivariate test for homogeneity of variances (10000 permutations) [136,137] using Bray-Curtis dissimilarity (differential abundance). Global and pairwise differences in community varaibility were assessed via permutation test of multivariate homogeneity of group dispersions (10’000 permutations, via the *betadisper* function). Community clustering according to anthropometric, community distances of naïve and rarefied communities were highly correlated and thus higher coverage naïve samples were used for analysis (Bray-Curtis: Mantel: r=0.9588, P<0.001; Procrustes: m^2^12=0.06012, r=0.9695, P<0.001; Jaccard: Mantel: r=0.9941, P<0.001; Procrustes: m^2^12=0.006143, r=0.9969, P<0.001).

#### DMM community clustering

Community clustering was performed via Dirichilet Multinomial Mixture modeling as implemented in the R packages microbiome (*microbiome_1.20.0*) [138] and DirichletMultinomial (*DirichletMultinomial_1.40.0*) [139] after centered log ratio transformation (CLR) implemented in *compositions* [140]. The optimal number of clusters was determined via Laplace goodness of fit optimization in a range of 1-15 clusters.

#### Differential abundance analysis

Taxon abundances were filtered by having normalized counts of at least 5 among 1% of samples (*DESeq2* median of ratios transformation). Negative binomial GLMs with Wald tests as implemented in *DESeq2* (including automated effect filtering) were employed to detect differentially abundant taxa for each time point separately as well as for all time points combined [141]. To reduce the effect of potential confounding effects we included subject age, BMI and sex as covariates and in addition the time point itself, for the combined dataset. P-values were adjusted via FDR and only comparisons excluding the small subset of uIBD patients were investigated further (Contr./CD, Contr./UC, CD/UC, Contr./all-IBD).

Abundance∼sex+ BMI (centered-scaled) + age (centered-scaled) [+ time point (BL/F1/F2)] + IBD pathology (Control, CD, UC, uIBD) Abundance∼sex+ BMI (centered-scaled) + age (centered-scaled) [+ time point (BL/F1/F2)] + Health status (Control/[CD, UC, uIBD]) Similar methods were applied to investigate the functional characteristics of the bacterial communities, as based on imputed bacterial functions and pathways via PICRUSt2 [131]. To generalize the single KO associations, we performed Gene Set Enrichment Analyses using a function wise score (-log10(P_Wald-test_) × sign(log fold change)) by multiplying the natural logarithm of P-values multiplied by the sign of fold change via clusterProfiler (4.4.4) using the *gseKEGG* function [47,142].

Taxon abundances were correlated with a reduced set of clinical variables (ASCA IgA, ASCA IgG, anti-GP2 IgA, anti-GP2 IgG, fecal calprotrectin, BSS) as well as polygenic risk scores for IBD in general and for CD or UC in particular. CLR transformed species abundances were correlated with core physiological measures via *ppcor* [143], combining the P-values of Spearman-, Kendall-, and Pearson correlations via Brown’s method to detect and incorporate the most consistent associations across different association measures [144]. The resulting P-values were FDR corrected and Spearman *ρ* was used for visualization via *ComplexHeatmap* (v2.14.0) [145].

#### Dysbiosis score

Dysbiosis scores based on the distribution of bacterial community members were calculated following Gevers *et al.* (2014) (Microbial Dysbiosis index/MD-index) and the more generalized General Microbiome Health Index (GMHI) as developed by Gupta *et al*. (2020) [38,39]. For the MD-index we combined significant taxa abundant in either CD or UC patients (uIBD not included) as compared to healthy/non-IBD cohort members at the baseline time point of the KINDRED cohort [log1p(sums(abundance CD, abundance UC)) / log1p(sums(non-IBD))]. GMHI was trained on the relative abundances at the baseline time point (training: 90%, testing: 10%) of the KINDRED cohort using code adapted from the Dutch Microbiome Project (”https://github.com/GRONINGEN-MICROBIOME-CENTRE/DMP/tree/main/gmhi“). Physiological and microbial inflammation markers/indices were analysed using linear models including relevant covariates (variable∼gender+scaled BMI + scaled Age + IBD (Contr., CD, UC, IBD)), followed by stepwise model selection to minimize AIC without significant loss of fit. The correlation of the MD/GMHI with alpha diversity was assessed with linear and quadratic fits and the best fit was selected via minimal AIC for each time point. Model fits were visualized via *base* R and *jTools* [135].

#### Heritability analyses

To estimate the heritability of single communities in a rather heterogeneous and patchy pedigree with many confounding variables we applied a recently published approach employing linear mixed models, as implemented in *lme4qtl* (*relatLmer*) [146]. Thus, we were able to incorporate fixed effects in our heritability estimates using a kinship matrix based on reported pedigree information (*kinship2*) (Sinnwell et al., 2014) and CLR transformed taxon abundances. Evaluating the models with and without covariates (AIC, based on ML fit) allows us to further evaluate whether taxon heritability/family association is influenced significantly by environmental factors or family relations alone. To assess patterns among close relatives we categorized relatedness patterns (related (>1st degree), first degree (sibling, parent)) for comparison, as well as calculated distances between healthy family members and diseased and eventually diseased individuals (onsets). Similar techniques were applied for the analyses of KO heritability, including subject age and sex as covariates. Gene Set Enrichment Analyses were performed on K-numbers using a function wise score based on full *h^2^* estimates via clusterProfiler (4.4.4) using the *gseKEGG* function [142]. Community-wide correspondence between taxonomic and functional community dissimilarity and relatedness (kinship matrix, plink2 based “Identity by State” distance derived from imputed SNP sets) was assessed via PROCRUSTES tests (5000 permutations).

#### Network analyses

Networks were generated individually for healthy and IBD (UC, CD) cohort subsets, as well as for the complete time point subsets. The node-based values (degree, betweenness, PageRank-index, eigenvalue-centrality, k-nearest neighbor degree) were calculated in *igraph* 1.2.4.1 [147–149]. Network-wide measures include centrality based assortativity [150], network diameter, radius, and size, density/clustering [151,152], and natural connectivity [48]. To assess whether bacteria were more important than expected by chance, observed centralities (mean of control subsamples) were compared against a permuted set of networks (10,000 times, combined for control subsamples) via one-sided Z-tests. Graphlet (4-node) frequency correlation (*orca* v1.1-1, Spearman) based Euclidean distance [153,154] and an edge sharing distance (frequency of shared pairwise correlations) to assess network similarity between the different subgroup networks (CD, UC, Contr., IBD at baseline, follow-up 1, follow-up 2).

#### Onset association

To predict the binary outcomes of disease onset based on diversity measures and physiological/clinical variables, we employed generalized linear models with a binomial error structure and a “clog-log” link function (“complementary-log-log”) as implemented in MASS [155]. The nonsymmetric complementary log-log link function (“clog-log”) was chosen, for its ability to better cope with unbalanced distributions between positive and negative outcomes in the target variable [156]. Baseline samples with the addition of one follow-up 1 sample, which developed disease in the second follow-up, were used for prediction (N_onset_=7), in the context of all remaining baseline samples or only healthy baseline controls. Models included either no covariates, or the established set of covariates (scaled age, sex, scaled BMI). Model fits were visualized via *base* R and *jTools* [135].

#### Models

IBD-onset next time point (1/0)∼ metric IBD-onset next time point (1/0)∼sex+ BMI (centered-scaled) + age (centered-scaled) + metric/clinical variable Data structure: BL (N_onset_=4, N_CD_=551, N_UC_=438, N_uIBD_=32, N_contr._=787); BL-controls (N_onset_=4, N_contr._=787); BL+ (N_onset_=7, N_CD_=551, N_UC_=438, N_uIBD_=32, N_contr._=787); BL-controls+ (N_onset_=7, Ncontr.=787)

## Data Availability

All KINDRED cohort data are stored and managed by the PopGen biobank at the Institute of Epidemiology at Kiel University, Germany. Researchers can apply for data access to the KINDRED data by submitting a research proposal, including the scientific background, research question, success prospects, study design, potential conclusions, and scientific collaborators of their study, at the local biobank P2N via the following form: http://www.uksh.de/p2n/Information+for+Researchers.html. Due to the informed consent obtained from the participants, phenotypes, as well as genotyping and all 16S rRNA gene-sequencing data, can not be deposited publicly.

Raw sequence data and relevant meta-data can be accessed online under the accession number PRJEB44440 (Malta IBD cases in remission), PRJEB47161 (Malta treatment naive IBD cases), PRJEB47162 (Malta controls) [63,64]; and data of the Swedish SIC-IBD inception cohort will be made available under the accession number PRJEB77933 at the European Nucleotide Archive (https://www.ebi.ac.uk/ena/) after acceptance of this manuscript at a peer-reviewed journal.

## ACKNOWLEDGEMENTS

We would like to thank Ilona Urbach, Ines Wulf, and Tonio Hauptmann of the IKMB microbiome laboratory and the staff of the IKMB sequencing facilities for their excellent technical support. This study was supported by the Deutsche Forschungsgemeinschaft (DFG) Research Unit 5042 (“miTarget-The Microbiome as a Therapeutic Target in Inflammatory Bowel Diseases”), the University of Malta, and the University of Örebro. Special thanks go to all participants of the KINDRED cohort study for enabling such a unique prospective study.

## DECLARATIONS

### Ethics approval

The study was performed in accordance with the principles of the Declaration of Helsinki. Approval was granted by the ethics committee of the Medical Faculty of Kiel University (AZ A117/13). Ethical approval for the Maltese cohorts was obtained from the University of Malta Research Ethics Committee (Ref 32/2017) and patients were recruited from the gastroenterology outpatient clinic at Mater Dei Hospital, Malta [63,64]. Ethical approval for the Swedish SIC-IBD cohort was obtained from the Uppsala Regional Ethics Committee (2010/313) and suspected IBD patients and controls were recruited from gastroenterological units at six Swedish hospitals.

### Consent to participate

Informed consent was obtained from all individual participants included in the study (or, in the case of children, from their parents). Consent for publication: Not applicable.

### Authors’ contributions

GJ, SS, MK, CB, AF and WL designed the study; JE, GJ, and WL, LT and KJ conducted the study; LT, KJ and PRa curated the data; PE, RK, JJ and DR contributed external samples and additional clinical analyses; PRa and IR analyzed the data; PRa, IR, CB, WL and AF drafted the paper; all authors take responsibility for the final content. All authors have read the manuscript, made significant intellectual contributions and approved the final version of the manuscript.

## Abbreviations used

CD: Crohn’s disease

IBD: inflammatory bowel disease

IC: informed consent

UC: ulcerative colitis

uIBD: undetermined inflammatory bowel disease

MD: index-Microbiome dysbiosis index

GMHI: General Microbiome Health Index

PRS: Polygenic Risk Scores

