## Supplemental Information for "First Insights into microbial changes within an Inflammatory Bowel Disease Family Cohort study"

**Abbreviations used:** CD-Crohn’s disease; IBD-inflammatory bowel disease; IC-informed consent; UC-ulcerative colitis; uIBD-undetermined inflammatory bowel disease, MD-index-Microbiome dysbiosis index; GMHI-General Microbiome Health Index; PRS-Polygenic Risk Scores

### Supplementary Methods

### Study cohort

#### *Eligibility criteria and enrolment*

The KINDRED includes IBD patients and their first- or second-degree relatives, with a minimum age of 7 years, from all over Germany. Families are deemed eligible if at least one IBD patient and one relative agree to participate. In a few exceptional cases, singleton IBD patients are also included provided that future participation of family members is likely. Most IBD patients are informed and invited into the study by their treating physicians or by IBD patient organizations in Germany, such as the German Crohn's disease / ulcerative colitis association (DCCV e.V.). The study is also advertised and promoted through a website (https://www.epi.uni-kiel.de/forschung/ced-studien/familienstudie).

Recruitment into the KINDRED follows a standardized protocol (**Suppl. Fig. 1**). If patients declare their interest in participating, they are sent a study set and a family documentation sheet. Due to data protection concerns, the study center is not allowed to contact healthy relatives directly without their prior expression of interest in participating. Therefore, every IBD patient is asked to relay details about the study to their family members and to inform the study center through the family sheet about potentially interested candidates. The latter are then formally invited into the study and receive a study set. The study set contains detailed information about the study, an informed consent form, a comprehensive participant questionnaire, and biomaterial tubes for blood, stool, and hair samples. IBD-affected study participants receive an additional questionnaire to be completed by their treating physician. If study sets are not returned within reasonable time, up to three reminders are sent in five-week intervals.

#### *Follow-up*

Study participants are re-contacted approximately every two years (**Suppl. Fig. 2**) and asked to fill in a questionnaire with health-related information and to provide new biomaterial samples (blood, stool, hair) at each study follow-up. IBD patients additionally receive a new physician questionnaire. At each follow-up, healthy study participants are asked whether they have been diagnosed with IBD in the meantime. Notably, in general, all initially non-affected participating family members are encouraged to contact the study center immediately after being newly diagnosed with IBD during the course of the study. In this case, the newly diagnosed IBD patient immediately receives a study set, including biomaterial collection tubes and participant and physician questionnaires so as to facilitate the collection of data from very early stages of the disease course (**Suppl. Fig. 3**).

The first follow-up (2-year follow-up) of the study participants that were recruited in 2013 started in 2015, the second follow-up (4-year follow-up) for this group in 2017, the third follow-up in 2019 (6-year follow-up), and the fourth follow-up of the participants recruited in 2013 started in 2021 (8-year follow-up; 5$^{th}$ data assessment). For participants that were recruited after 2013, the respective follow-up assessments are conducted after the appropriate time intervals.

### Data and biomaterial collection

The KINDRED study prospectively collects biomaterial (blood, stool, hair) as well as comprehensive sociodemographic, socioeconomic, clinical, and lifestyle data from participant questionnaires (adapted to the individual status as IBD patient, healthy relative, and participating child/adolescent; please see below), plus data from physician questionnaires in the case of IBD patients. The study aims to collect complete questionnaire data and biomaterial from each participant but, if a participant refuses to provide some of the data or biomaterial, their incomplete contribution is also accepted.

#### *Questionnaires*

Separate questionnaires have been developed for healthy family members and IBD patients (for details, see **Suppl. Table 1**). Both questionnaires have a common backbone of questions related to sociodemographic and socioeconomic characteristics, general health status, lifestyle factors, and quality of life [1,2] (**Suppl. Table 1**). The wording is slightly modified for children and adolescents. In addition, a validated and standardized web-based food frequency questionnaire (FFQ) [3] and a set of physical activity questions [4] are administered to assess the study participants' diet and their usual physical activities during the preceding 12 months.

The questionnaire for IBD patients also includes questions about their IBD (**Suppl. Table 1**), thereby complementing the physician questionnaire that patients are asked to have filled out by their treating physician during the next visit. The physician questionnaire (**Suppl. Table 1**) enquires detailed disease-related information and includes established and validated questions and scores, such as the Harvey-Bradshaw-Index (HBI), the Mayo Score, and the Crohn's Disease Activity Index (CDAI). All self-reported IBD diagnoses were validated against the physician questionnaires or other medical records.

#### *Collection, work-up, and storage of biomaterial*

At the initial (baseline) assessment, blood, stool, and hair samples are collected from all participants (**Suppl. Table 2**), including approximately 35 ml blood from adult participants and adolescents (12 to 18 years) and 15 ml from children between 7 and 11 years of age. Study participants also receive sets for the self-collection of stool and hair, accompanied by written instructions on how to collect the sample. Upon receipt at the study center, the biomaterial samples are pseudonymized and stored in the local popgen biobank at Kiel University (**Suppl. Table 2**).

**Data management, and privacy protection**

All clinical data are pseudonymized and stored separately from the identifying data in a central study database at the Institute of Epidemiology and the popgen biobank at Kiel University. The data management and the privacy protection concept of popgen has been reviewed and approved by the independent data protection authorities of Schleswig-Holstein (ULD) and the ethics committee of the Medical Faculty of Kiel University.

In this work, we focused on keeping our study data in accordance with the FAIR principles [5]. That means that data must be findable, accessible, interoperable and reusable. Also, as we used data that contains a fingerprint of the donor, data protection is of utmost importance. For this purpose, we established andata management tool called iRods (Rule-Oriented Data management systems) for internal use {iRODS Consortium. https://irods.org}. Each user has to log in to the system with an account via API and has access only to content intended for them. Data is shared between the users via subproject groups to only individuals who are in the authorized group. Users outside this group have no access to the data stored there. This guarantees secure use of sensitive data. In addition, the rights system allows a distinction to be made as to whether a user only has viewing rights or can modify the data. The system allows metadata to be stored for the project as json-files. This not only allows the metadata to be closely located to the research data, but also to be searchable directly within the system. Likewise, it also allows the data to be reused at a later date. The metadata input mask was created for this purpose in compliance with the STORMS checklist intended for microbiome studies [6].

**Supplemental Results:**

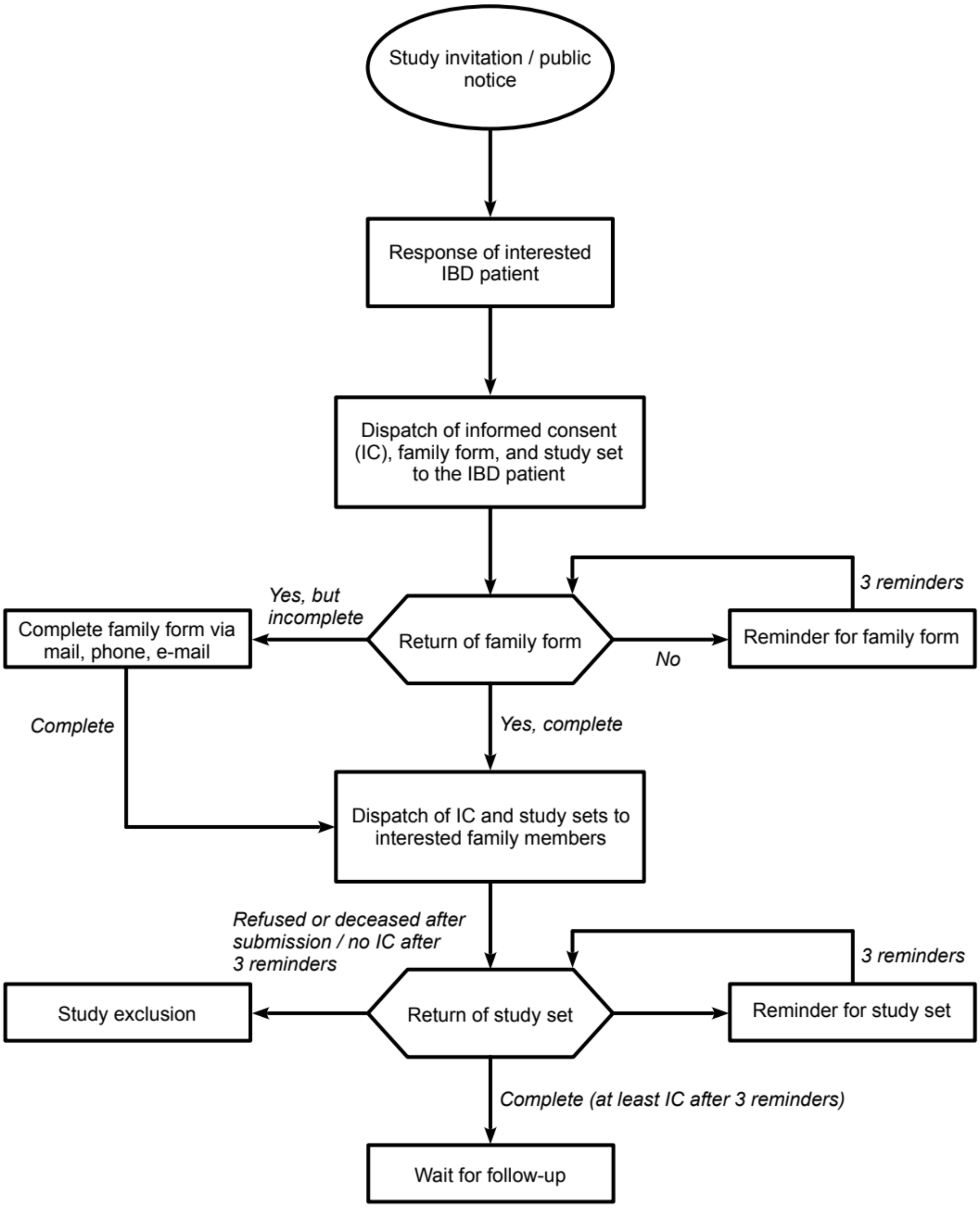


**Figure S1:** Process of study participant enrolment. Abbreviations: IBD, inflammatory bowel disease; IC, informed consent.

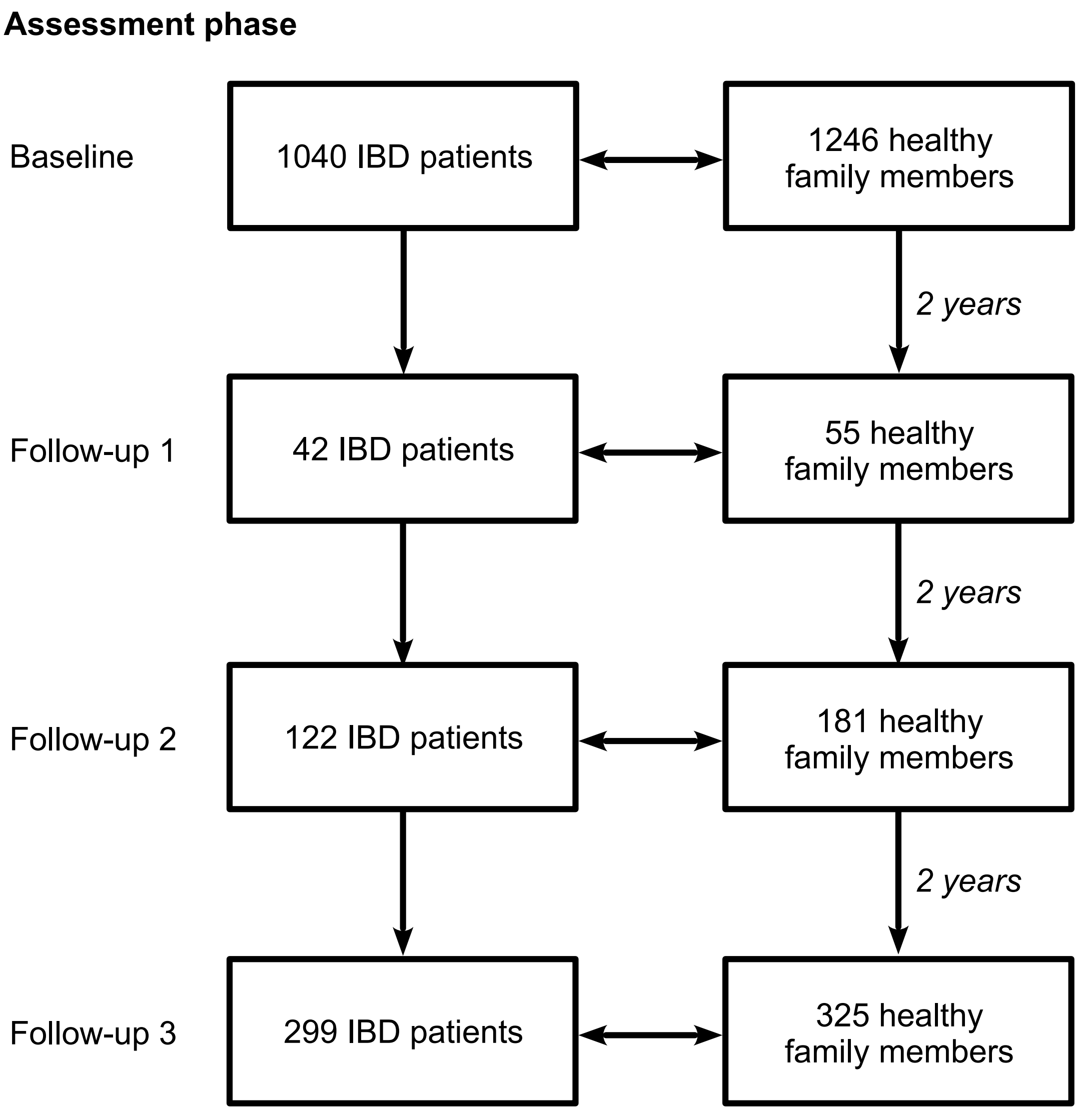


**Figure S2:** Current number of study participants (IBD patients and healthy family members) by assessment phase in the prospective Kiel IBD Family Cohort. Abbreviations: IBD, inflammatory bowel disease.

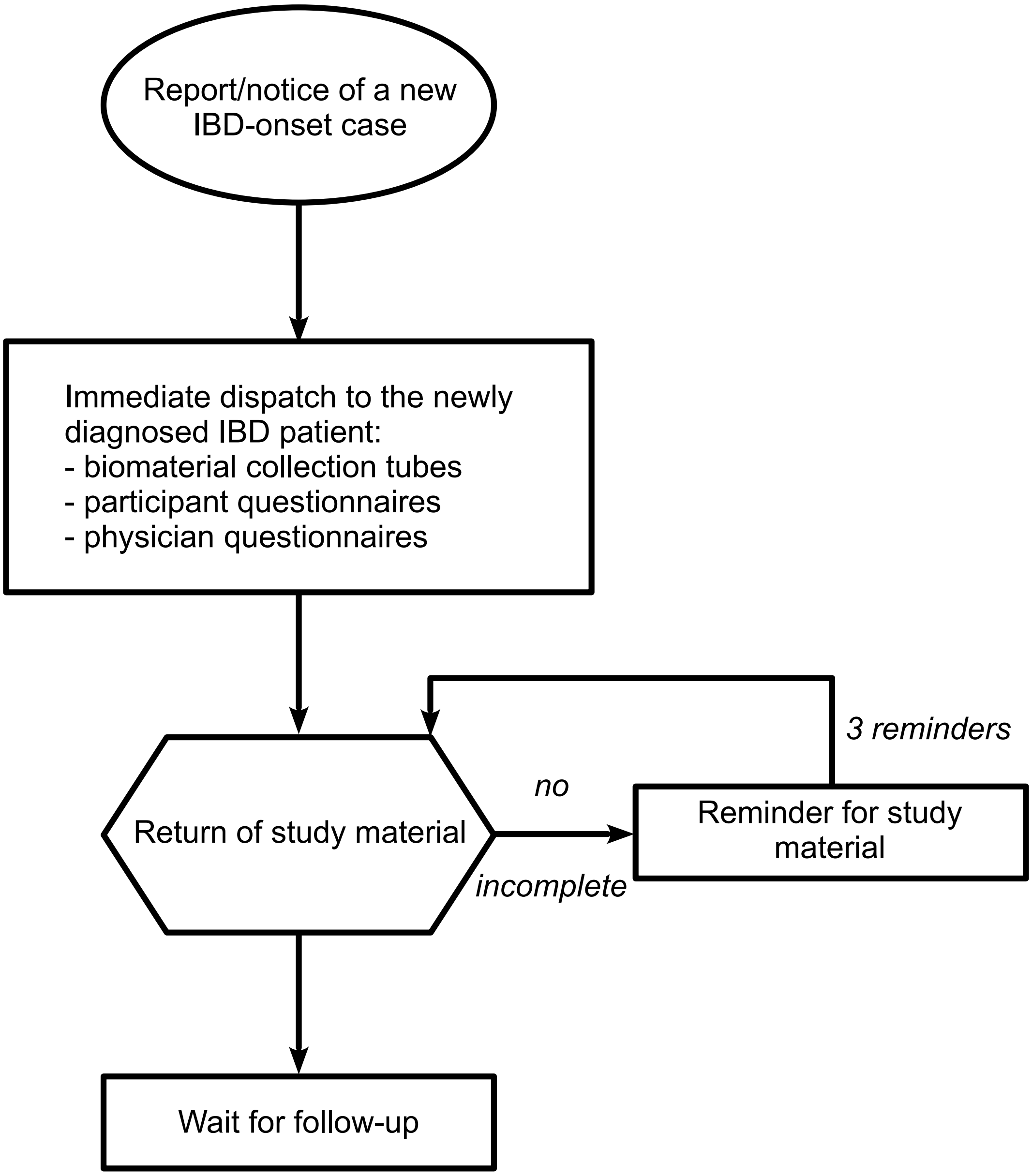


**Figure S3:** Procedure following the report of a new IBD onset case during study follow-up.

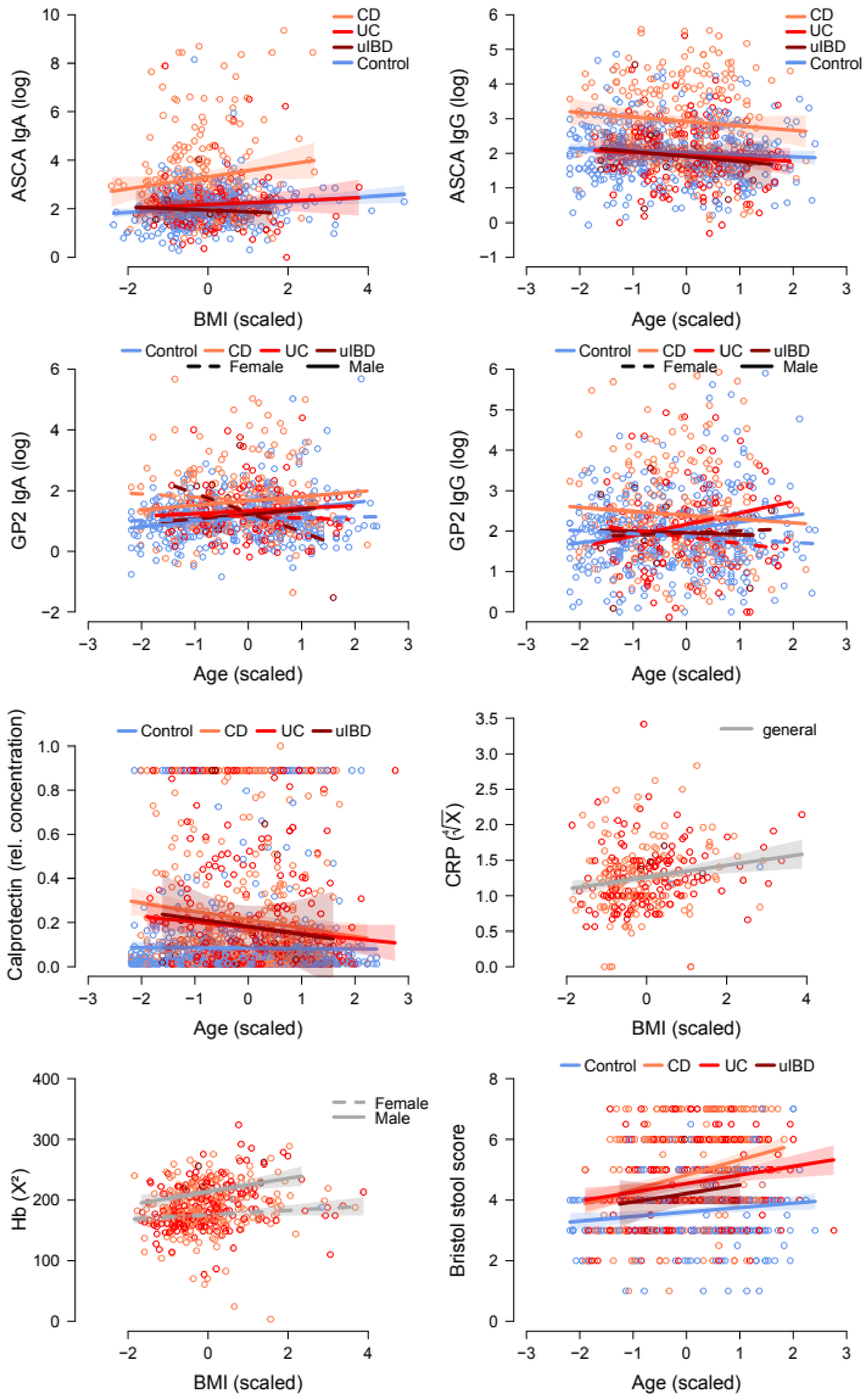

CD
UC
uIBD
Control
ASCA IgA (log)
BMI (scaled)
ASCA IgG (log)
Age (scaled)
Control CD UC uIBD
Female Male
GP2 IgA (log)
Age (scaled)
GP2 IgG (log)
Age (scaled)
Calprotectin (rel. concentration)
Age (scaled)
general
CRP ($\sqrt[4]{X}$)
BMI (scaled)
Female
Male
Hb ($X^2$)
BMI (scaled)
Bristol stool score
Age (scaled)

**Figure S4:** Scatterplots show the association of immune / physiological parameters (anti ASCA-IgA/IgG, anti GP2-IgA/IgG, relative calprotectin levels, CRP, Hb, Bristol stool score) with the main anthropometric variables (IBD condition, gender, BMI, age). Plots display the modelling results after model selection minimizing AIC. Model statistics are in Table 2. The polygons highlight the 95% CI.

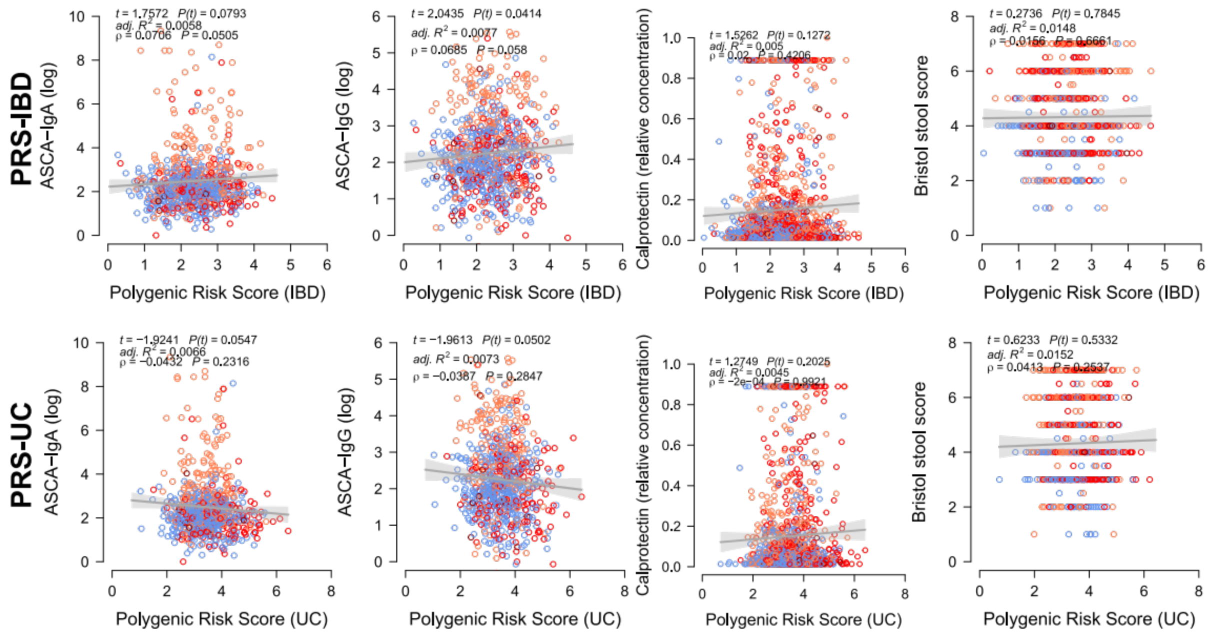


**Figure S5:** Scatterplots show the association of immune / physiological parameters (anti ASCA-IgA/IgG, relative calprotectin levels, Bristol stool score) with the *LDpred2* based polygenic risk scores for UC and IBD using LMs correcting for relevant covariates (age, BMI, gender), as well as Spearman rank correllation results. Grey polygons highlight the 95% CI.

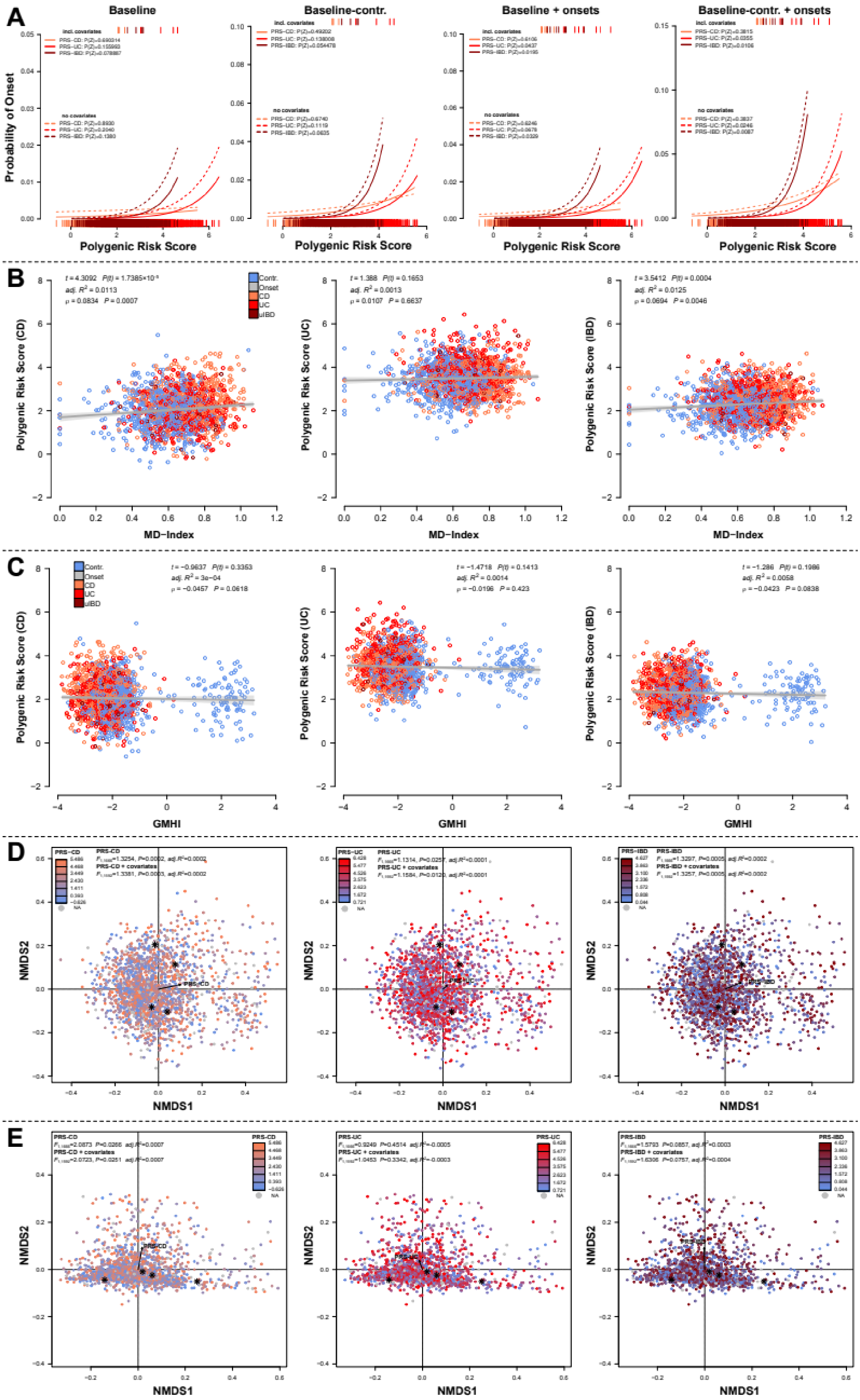

A
Baseline
Baseline-contr.
Baseline + onsets
Baseline-contr. + onsets
Probability of Onset
Polygenic Risk Score
B
Contr.
Onset
CD
UC
uIBD
Polygenic Risk Score (CD)
Polygenic Risk Score (UC)
Polygenic Risk Score (IBD)
MD−Index
C
GMHI
D
NMDS2
NMDS1
E

**Figure S6: (A)** Analysis of disease onset prediction via logistic regression of PRS, based on different slices of data, all baseline samples ($N_{onset}$=4, $N_{CD}$=551, $N_{UC}$=438, $N_{uIBD}$=32, $N_{contr.}$=787), baseline controls with 4 onset samples ($N_{onset}$=4, $N_{contr.}$=787), all baseline samples with onset cases of later time points ($N_{onset}$=7, $N_{CD}$=551, $N_{UC}$=438, $N_{uIBD}$=32, $N_{contr.}$=787) as well as baseline controls with the supplemental onset cases ($N_{onset}$=7, $N_{contr.}$=787). We used used models accounting for or ignoring potential covariates (age, BMI, sex). Depicted are predicted probabilities of the individual models for each PRS and colored disease and line type differ by model type (w/o covariates). Weak but significant predictions are most consistently possible for IBD genetic predisposition, and to a lesser amount for UC PRS. **(B)** Correlation of PRS with the microbial dysbiosis index (MD-index, Gevers et al. 2014) show signifcant positive relationships between microbial dysbiosis and genetic predisposition for CD and IBD, **(C)** while GMHI shows no significant association to genetic predisposition to IBD. PRS are based on *LDpred2* prediction [7]. **(D)** Genetic predisposition for either CD UC or IBD in general shows significant correlation to taxonomic community distance, **(E)** while functional community distance is only weakly correlated to CD-PRS.

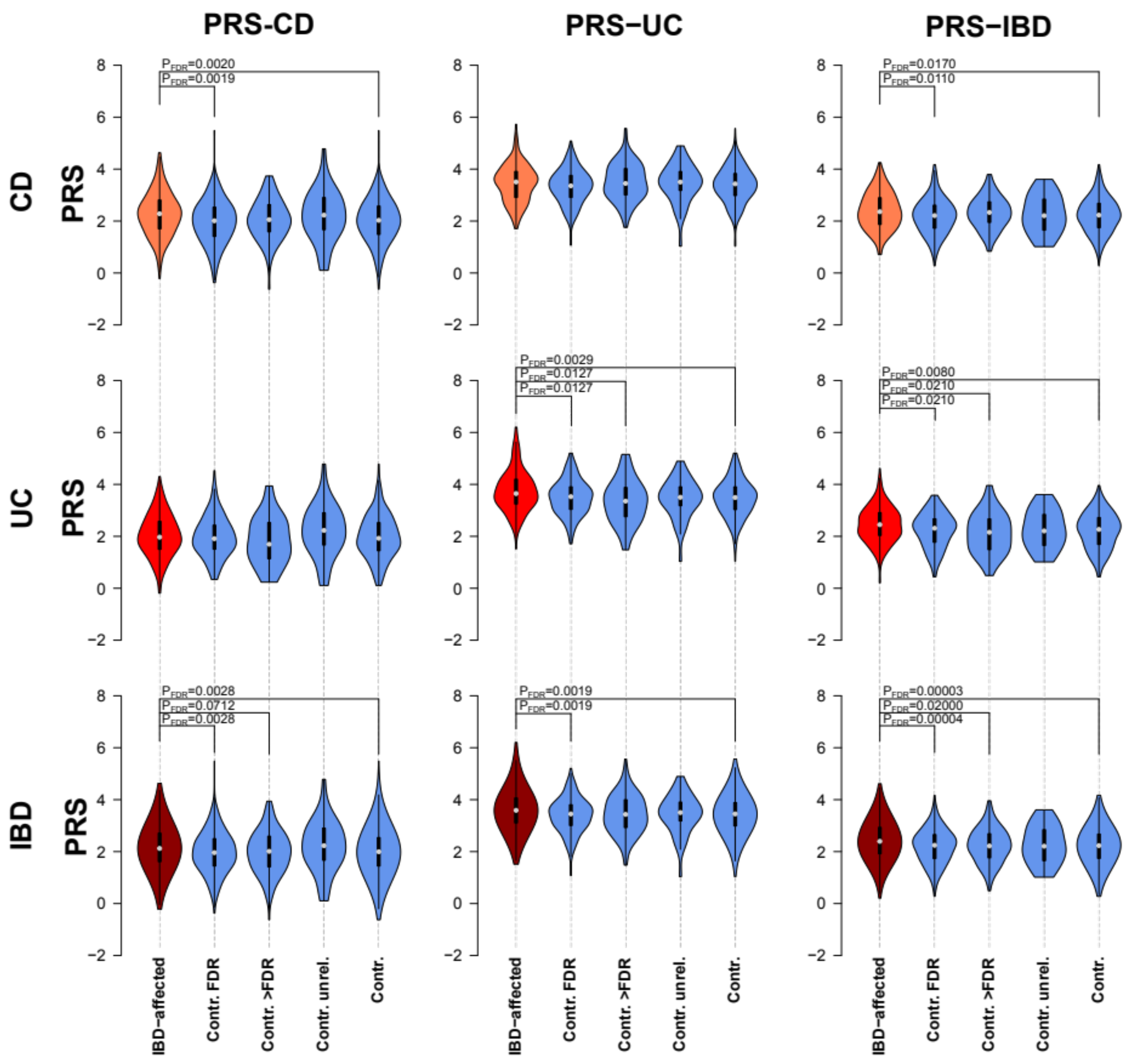


**Figure S7: (A)** Differences of *LDpred2* [7] based polygenic risk scores for CD, UC or general IBD affected individuals (CD/UC/IBD) and their healthy first degree relatives, distant relatives, unrelated controls, or all available healthy controls. All pairwise comparisons were made via Wilcoxon rank test and corrected for multiple testing via FDR. Affected individuals have on average a higher PRS than even their healthy family members, hinting towards a larger accumulation of risk variants in these individuals even in comparison to closely related family members. Further more, CD-PRS and UC-PRS are specific to the respective patient population, while the combined IBD-PRS is more general.

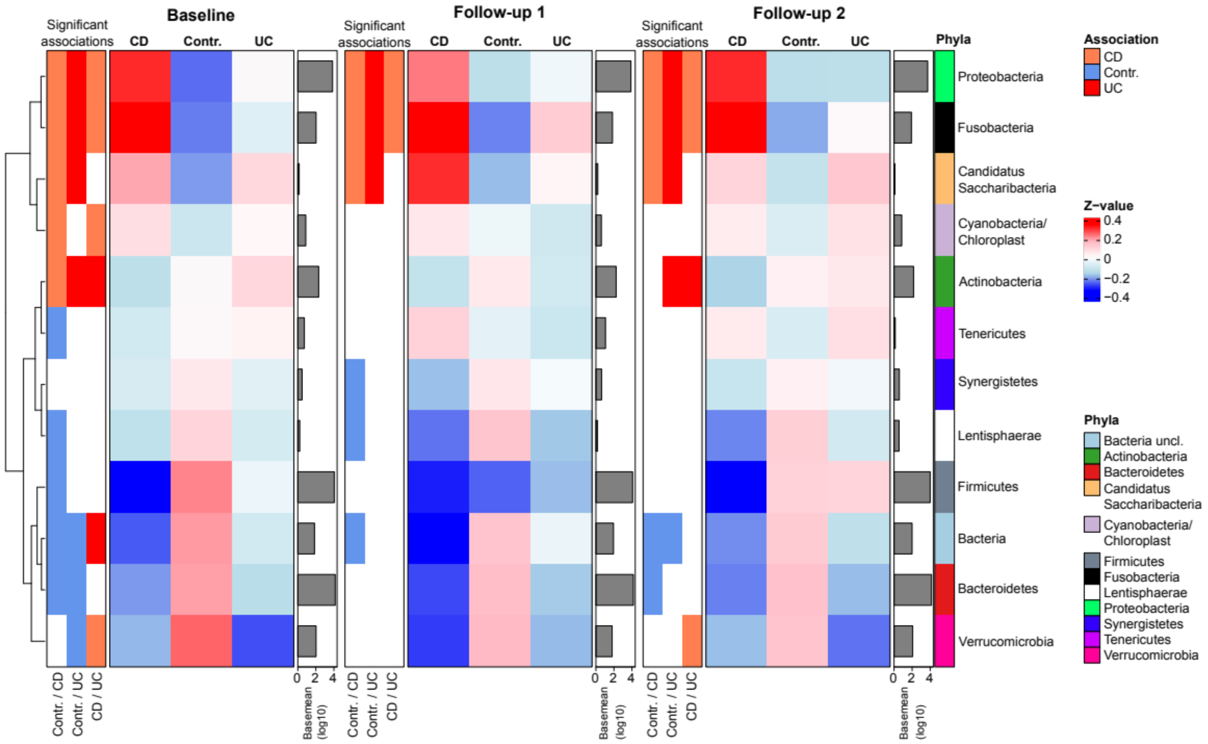


**Figure S8:** Differential abundance analyses of Phylum abundances between the main health conditions (healthy controls, CD, UC) across time points (BL; F1, F2, see Table S7). Color bars on the left highlight the direction of significant differential abundance change in the individual comparisons among health condition (Contr./CD, Contr,/UC, CD/UC, Contr./IBD ;$P_{FDR} \leq 0.05$) and right hand color bar signifies phylum membership. Z-values are based on group means deviation from the overall average, and base means derived from DESeq2 analysis are shown in the side barplots.

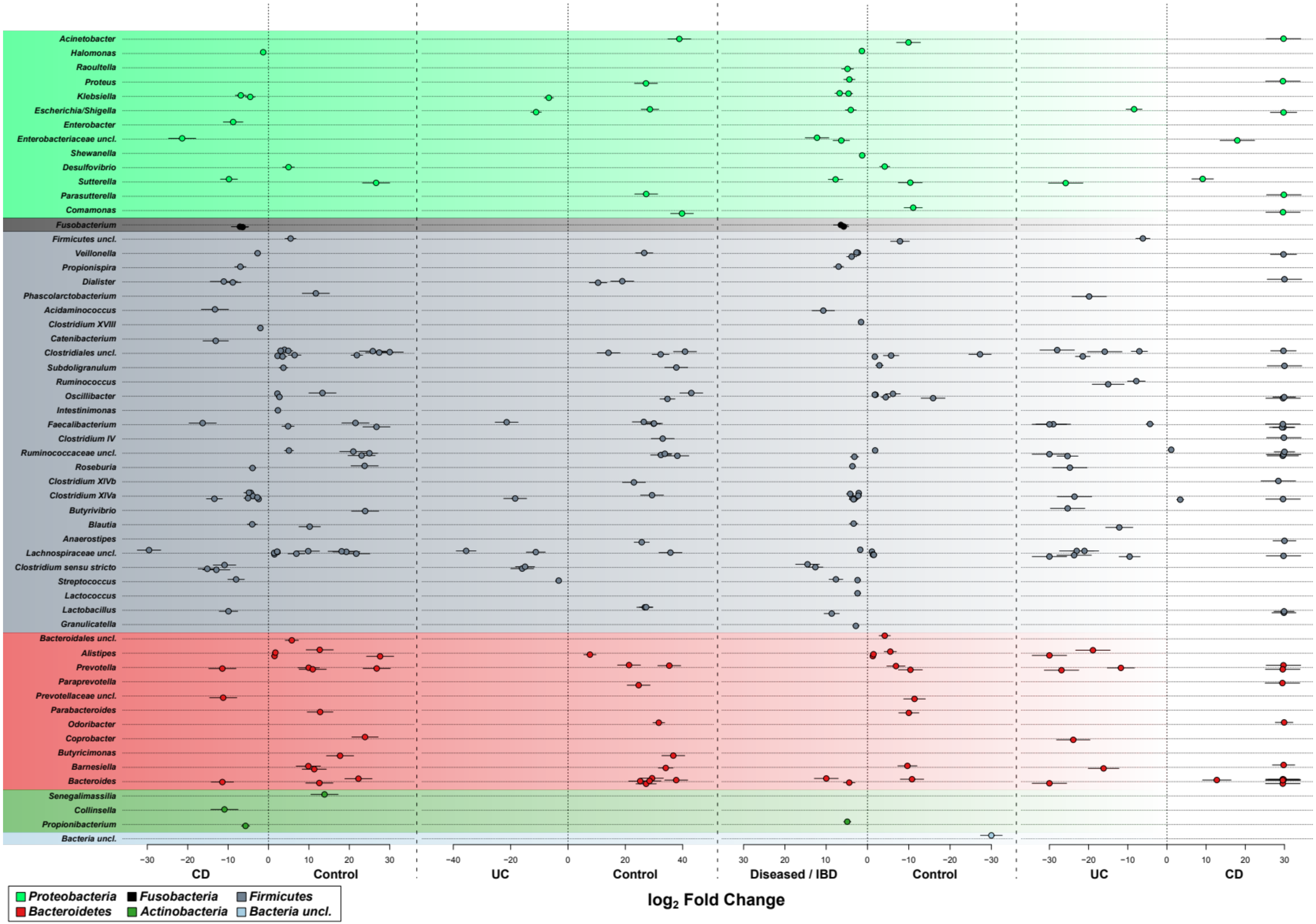

Acinetobacter
Halomonas
Raoultella
Proteus
Klebsiella
Escherichia/Shigella
Enterobacter
Enterobacteriaceae uncl.
Shewanella
Desulfovibrio
Sutterella
Parasutterella
Comamonas
Fusobacterium
Firmicutes uncl.
Veillonella
Propionispira
Dialister
Phascolarctobacterium
Acidaminococcus
Clostridium XVIII
Catenibacterium
Clostridiales uncl.
Subdoligranulum
Ruminococcus
Oscillibacter
Intestinimonas
Faecalibacterium
Clostridium IV
Ruminococcaceae uncl.
Roseburia
Clostridium XIVb
Clostridium XIVa
Butyrivibrio
Blautia
Anaerostipes
Lachnospiraceae uncl.
Clostridium sensu stricto
Streptococcus
Lactococcus
Lactobacillus
Granulicatella
Bacteroidales uncl.
Alistipes
Prevotella
Paraprevotella
Prevotellaceae uncl.
Parabacteroides
Odoribacter
Coprobacter
Butyricimonas
Barnesiella
Bacteroides
Senegalimassilia
Collinsella
Propionibacterium
Bacteria uncl.
CD
Control
UC
Control
Diseased / IBD
Control
UC
CD
log2 Fold Change
Proteobacteria
Fusobacteria
Firmicutes
Bacteroidetes
Actinobacteria
Bacteria uncl.

**Figure S9:** Differential abundance analyses of ASVs based on the first follow-up time point (F1, Table S9). Displayed are the log fold changes for each ASV clustered by genus classification, including standard errors of the fold changes as indicated by individual doted lines. Color bars indicate the phylum membership. DA only displays signifcant differential abundance for the respective comparison/contrast ($P_{FDR} \leq 0.05$).

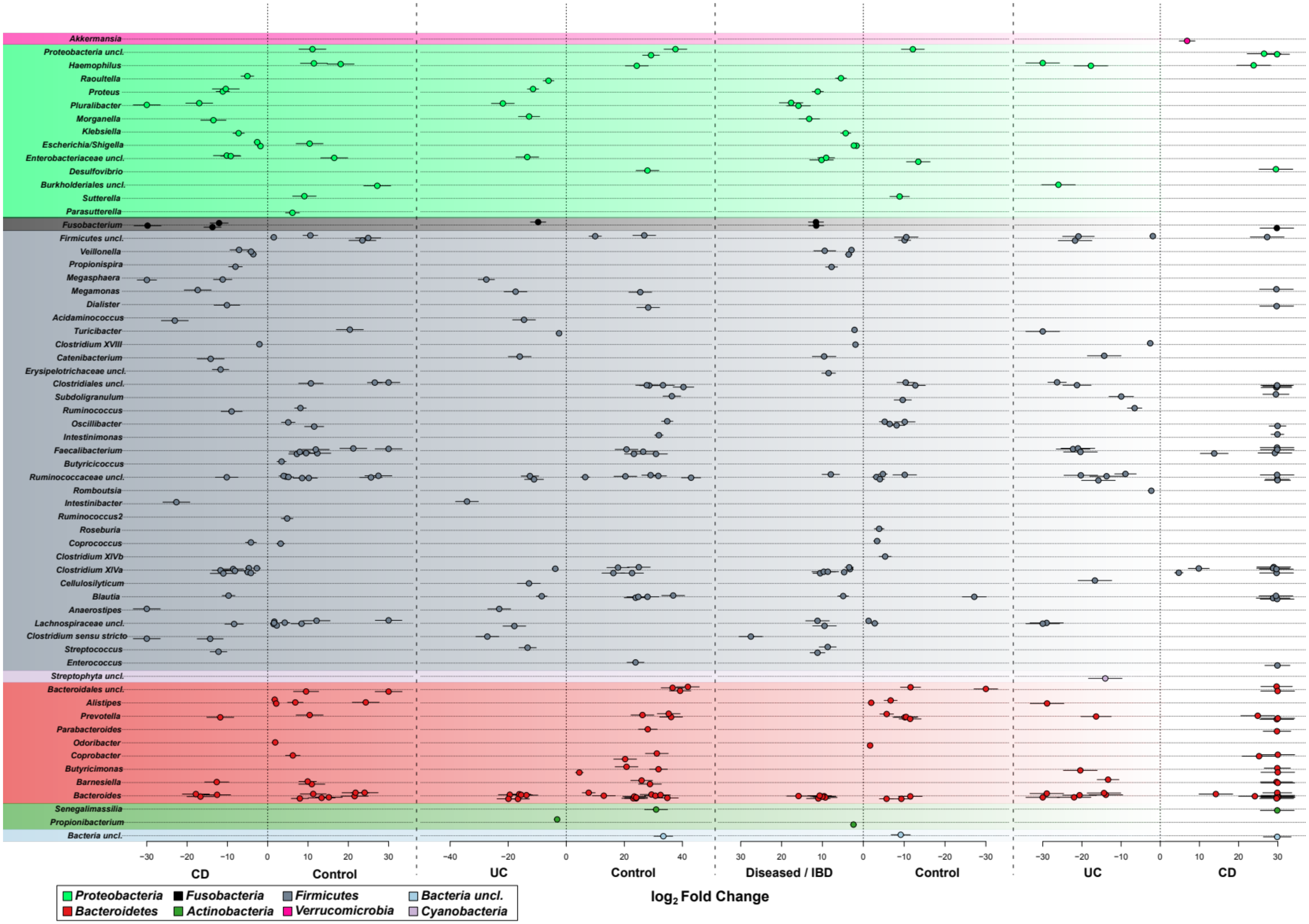

Akkermansia
Proteobacteria uncl.
Haemophilus
Raoultella
Proteus
Pluralibacter
Morganella
Klebsiella
Escherichia/Shigella
Enterobacteriaceae uncl.
Desulfovibrio
Burkholderiales uncl.
Sutterella
Parasutterella
Fusobacterium
Firmicutes uncl.
Veillonella
Propionispira
Megasphaera
Megamonas
Dialister
Acidaminococcus
Turicibacter
Clostridium XVIII
Catenibacterium
Erysipelotrichaceae uncl.
Clostridiales uncl.
Subdoligranulum
Ruminococcus
Oscillibacter
Intestinimonas
Faecalibacterium
Butyricicoccus
Ruminococcaceae uncl.
Romboutsia
Intestinibacter
Ruminococcus2
Roseburia
Coprococcus
Clostridium XIVb
Clostridium XIVa
Cellulosilyticum
Blautia
Anaerostipes
Lachnospiraceae uncl.
Clostridium sensu stricto
Streptococcus
Enterococcus
Streptophyta uncl.
Bacteroidales uncl.
Alistipes
Prevotella
Parabacteroides
Odoribacter
Coprobacter
Butyricimonas
Barnesiella
Bacteroides
Senegalimassilia
Propionibacterium
Bacteria uncl.
CD
Control
UC
Control
Diseased / IBD
Control
UC
CD
log2 Fold Change
Proteobacteria
Fusobacteria
Firmicutes
Bacteria uncl.
Bacteroidetes
Actinobacteria
Verrucomicrobia
Cyanobacteria

**Figure S10:** Differential abundance analyses of ASVs based on the second follow-up time point (F2, Table S10). Displayed are the log fold changes for each ASV clustered by genus classification, including standard errors of the fold changes as indicated by individual doted lines. Color bars indicate the phylum membership. DA only displays signifcant differential abundance for the respective comparison/contrast ($P_{FDR} \leq 0.05$).

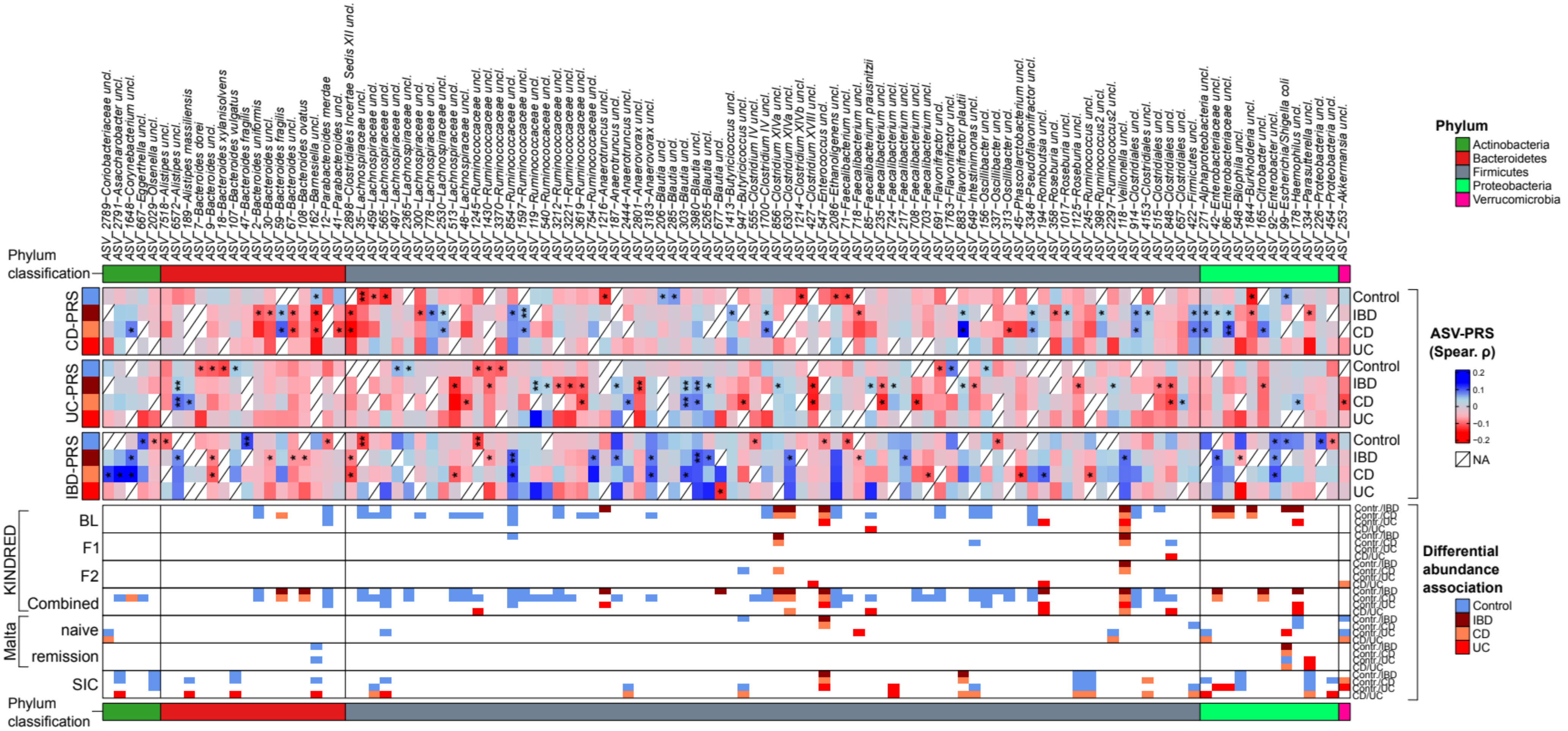


**Figure S11:** Partial correlation of CLR transformed taxon abundances with LDpred2 derived polygenic risk scores for CD, UC, and general IBDvia *ppcor* (Kim 2015), combining the *P* values of Spearman-, Kendall-, and Pearson correlations via Brown's method and corrected via FDR (Brown 1975). Correlations were adjusted for age, gender, and BMI. Spearman $\rho$ is used to visualize correlation strength between taxa and clinical measures

(# $P_{FDR}$≤0.1000, * $P_{FDR}$≤0.0500, ** $P_{FDR}$≤0.0100, *** $P_{FDR}$≤0.0010). Overlapping, significant patterns of differential abundance for the respective taxa in the KINDRED cohort, Maltese- and Swedish SIC cohort are indicated in the bottom color bars (see Table S12).

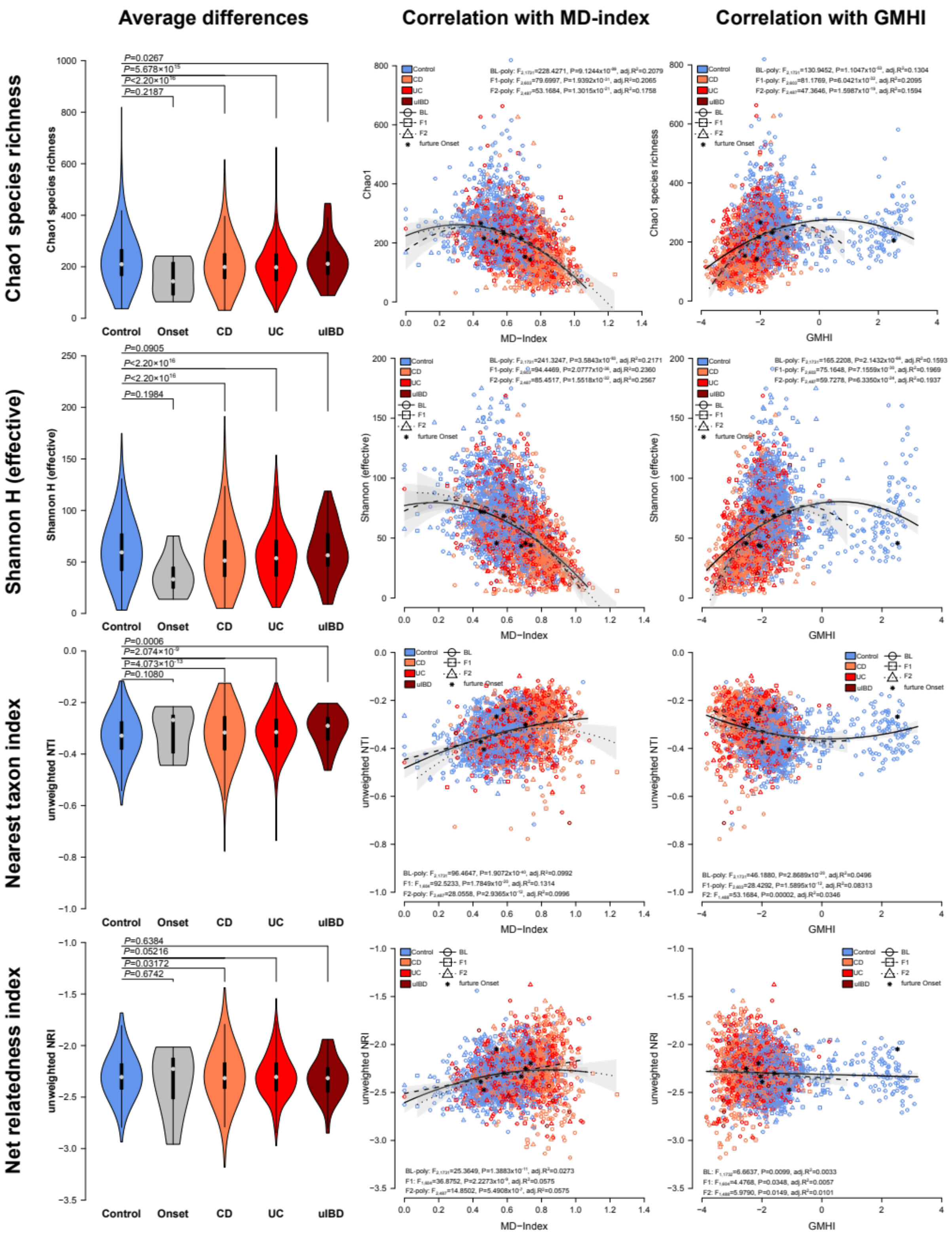


**Figure S12:** Alpha diversity analyses focussing on differences of onset cases, CD patients, UC patients, and uIBD patients to healthy control individuals, based on species richness (Chao1), general complexity (Shannon H), and relative phylogenetic relatedness at high phylogenetic levels (NTI) or across the complete phylogenetic tree (NRI). Differences were assessed via pairwise Wilcoxon tests. Correlation of dysbiosis indices MD and GMHI [8,9] with the different alpha diversity measures in the three available time points (Chao1: BL (poly): $F_{2,1809}$=226.97, P<2.2×$10^{-16}$, adj.$R^2$=0.1997; F1 (poly): $F_{2,645}$=83.778, P<2.2×$10^{-16}$, adj.$R^2$=0.2037; F2 (poly): $F_{2,536}$=55.745, P<2.2×$10^{-16}$, adj.$R^2$=0.1691; Shannon:BL (poly): $F_{2,1809}$=243.43, P<2.2×$10^{-16}$, adj.$R^2$=0.2112; F1 (poly): $F_{2,645}$=101.80, P<2.2×$10^{-16}$, adj.$R^2$=0.2376; F2 (poly): $F_{2,536}$=89.468, P<2.2×$10^{-16}$, adj.$R^2$=0.2475; NRI: BL (poly): $F_{2,1809}$=28.311, P=7.823×$10^{-13}$, adj.$R^2$=0.02928; F1 (lin.): $F_{2,645}$=38.058, P=1.212×$10^{-9}$, adj.$R^2$=0.05417; F2 (poly): $F_{2,536}$=16.126, P=1.581×$10^{-7}$, adj.$R^2$=0.05324; NRI: BL (poly): $F_{2,1809}$=96.927, P<2.2×$10^{-16}$, adj.$R^2$=0.09579; F1 (lin.): $F_{2,645}$=93.097, P<2.2×$10^{-16}$, adj.$R^2$=0.1246; F2 (poly): $F_{2,536}$=31.103, P=1.66×$10^{-13}$, adj.$R^2$=0.1006; linear models) [10–12]. Grey polygons highlight the 95% CI and "*" highlight the still healthy, future onset patients. Poly indicates a second order polynomial fit, instead of a linear model fit. See Table S13.

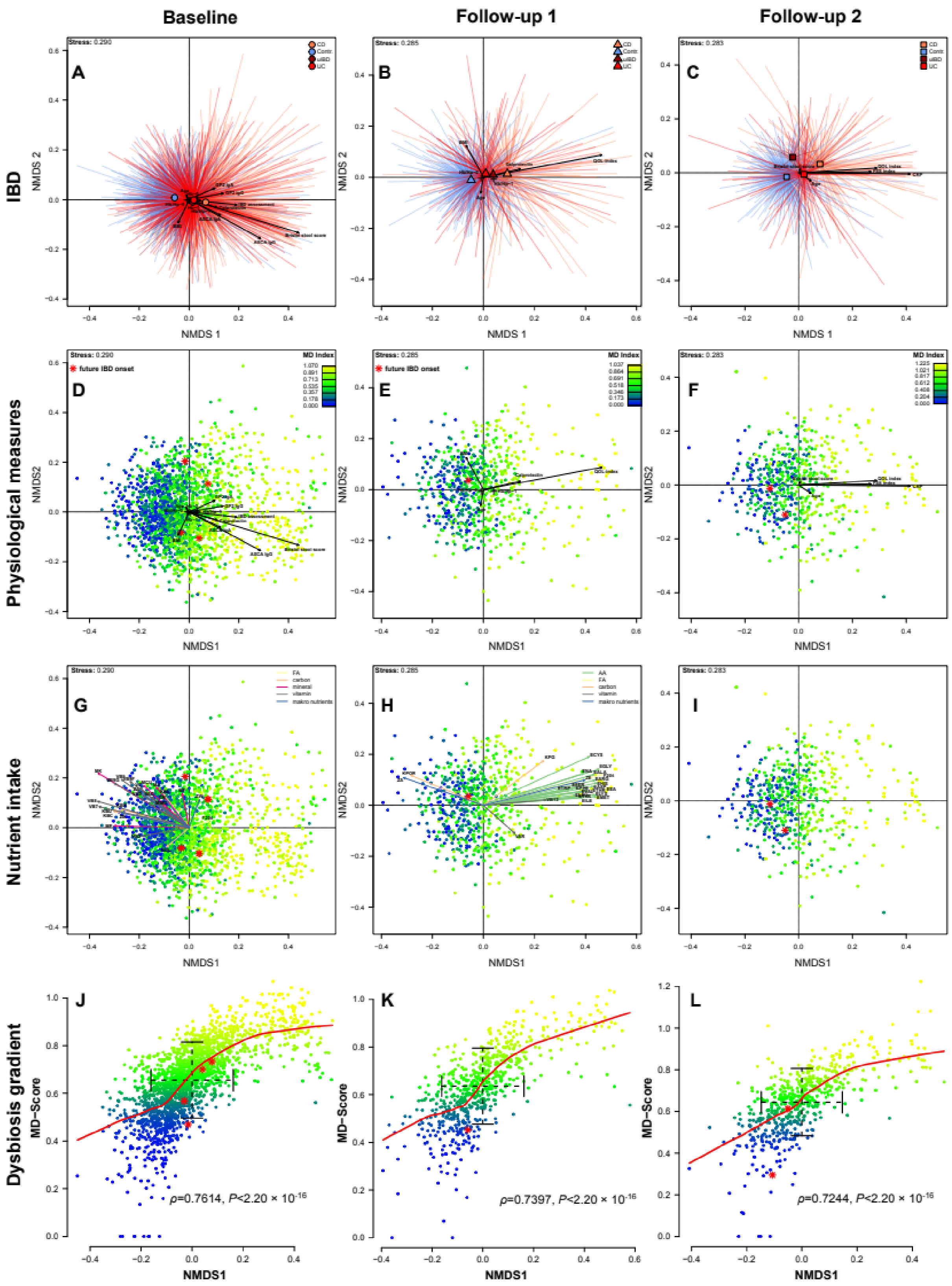


**Figure S13: (A)** Non-metric Multidimensional Scaling (NMDS) of Bray-Curtis distances among baseline samples, **(B)** follow-up 1, and **(C)** follow-up 2, displaying the significant clustering by health conditions and significant correlations of clinical inflammation measures with community distance (see Table S16, Table S17, Table S23). **(D)** NMDS displaying the gradient of community dysbiosis as expressed by MD-index [8], across the three time points, in parallel with significantly correlated clinical measures of inflammation and healthy onset cases highlighted in red (*, develop IBD until the next follow-up). **(G, H, I)** Nutrition (approximated normalized nutrient uptake) was also significantly correlated with community distances. Arrow colors represent different nutrient groups (AA-aminoacids, FA-fatty acids, carbon-carbohydrates, minerals-trace elements, vitamins, makro nutrients-larger nutrient clusters (i.e. proteins, fats, water)). **(J, K, L)** Correlation of MD-index and the first NMDS axis showing a clear gradient of dysbiosis in the community (Spearman rank correlation). Onset cases are distributed in the range of standard deviation around the mean of the community distribution (NMDS1) and the severity of dysbiosis (MD-index).

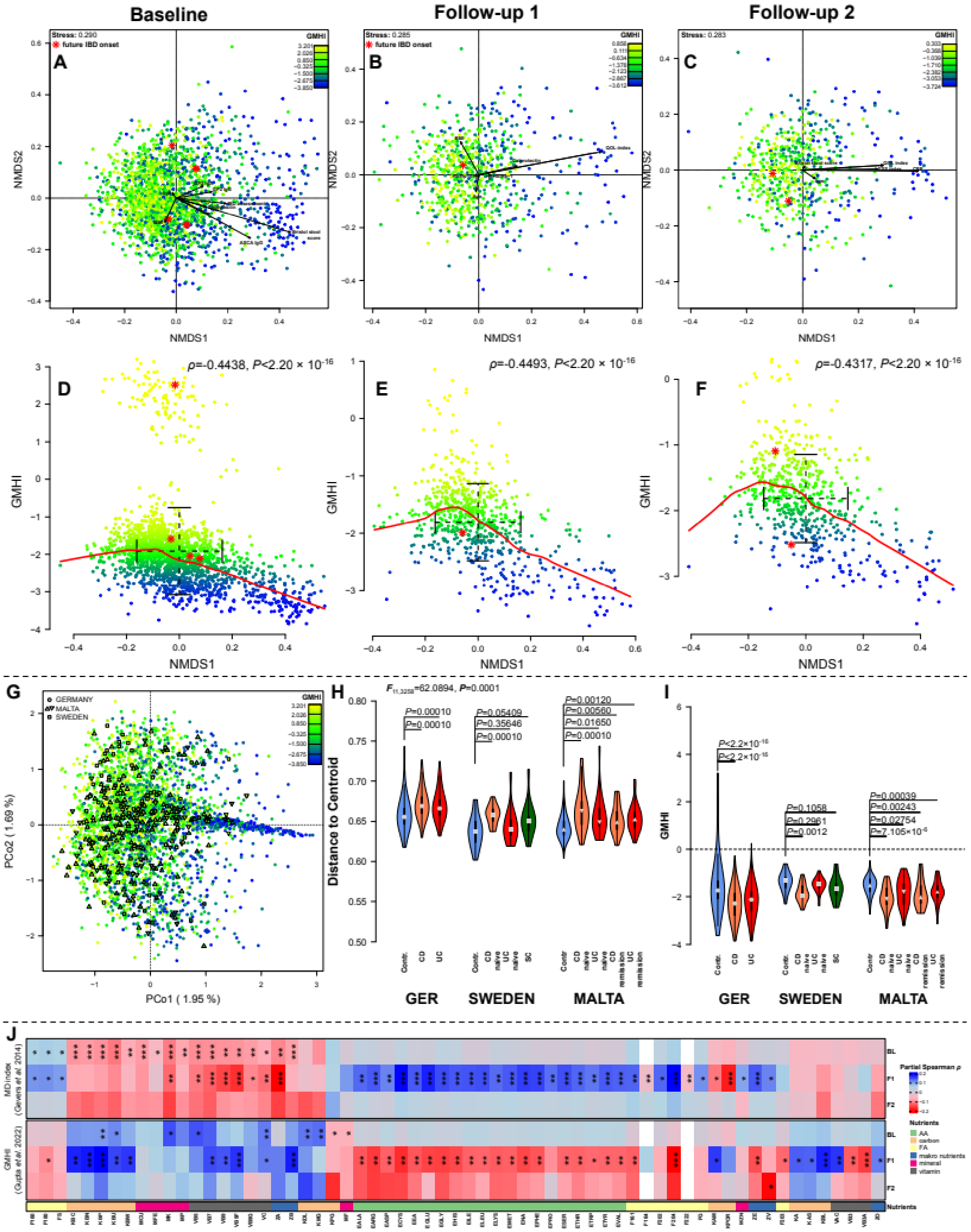


**Figure S14: (A)** Non-metric Multidimensional Scaling (NMDS) of Bray-Curtis distances among baseline samples, **(B)** follow-up 1, and **(C)** follow-up 2, displaying the significant gradient of community dysbiosis as expressed by GMHI [9], across the three time points, in parallel with significantly correlated clinical measures of inflammation and healthy onset cases highlighted in red (*, will develop IBD until the next follow-up, see Table S16, Table S17, Table S23). **(D, E, F)** Correlation of MD-index and the first NMDS axis showing a clear gradient of dysbiosis in the community (Spearman rank correlation). Onset cases are distributed in the range of standard deviation around the mean of the community distribution (NMDS1) and the severity of dysbiosis (GMHI). **(G)** Principle coordinate analysis of german swedish and maltese samples, highlighting the transferrability of GMHI across cohorts (derived from german samples). **(H)** Community variability between health/IBD conditions within and between the german, swedish and maltese cohorts showing a common theme of increased variability in IBD cases compared to healthy controls, as based on Jaccard distance. **(I)** Mean differences of GMHI between healthy individuals and diseased groups within different IBD cohorts. The strongest and most consistent differences occur between healthy and CD individuals observable accross cohorts (Wilcoxon test). **(J)** Partial correlation of MD-index and GMHI with approximated and scaled nutrient intake via *ppcor* (Kim 2015), combining the *P* values of Spearman, Kendall and Pearson correlations via Brown's method and corrected via FDR (Brown 1975). Correlations were adjusted for age, gender, and BMI (# $P_{FDR} \leq 0.1000$, * $P_{FDR} \leq 0.0500$, ** $P_{FDR} \leq 0.0100$, *** $P_{FDR} \leq 0.0010$).

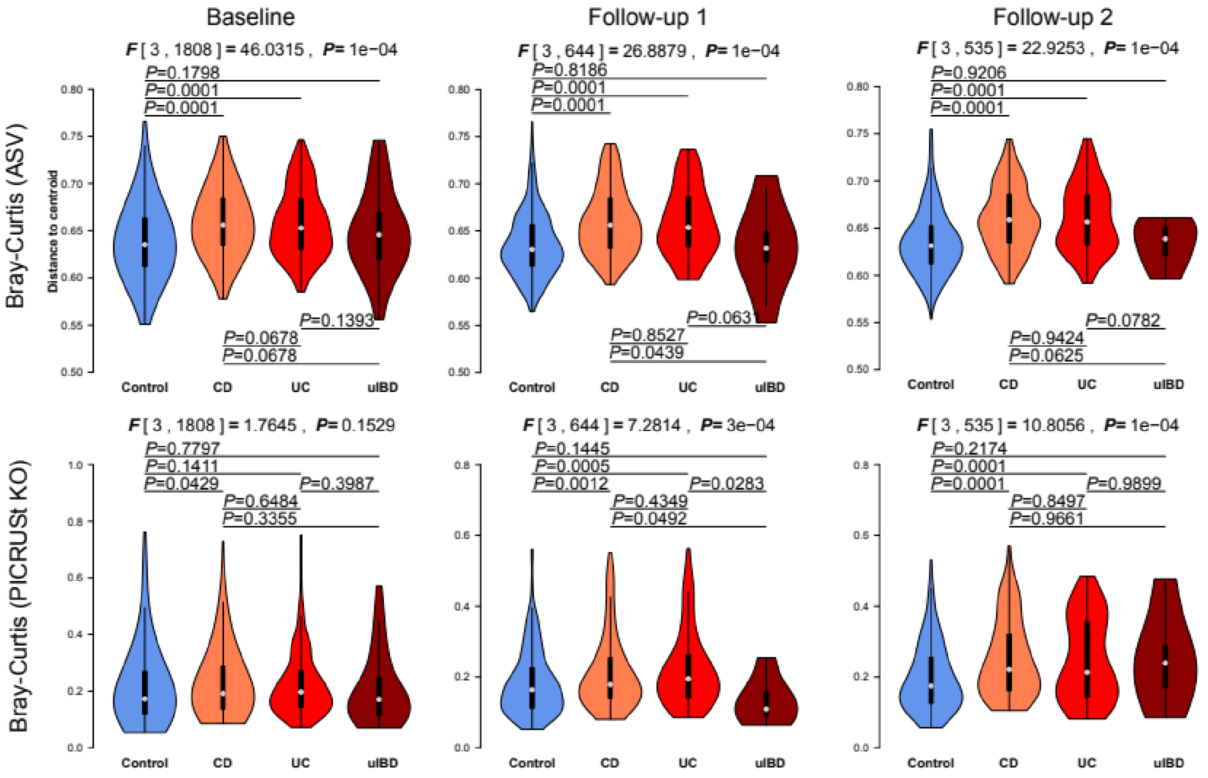


**Figure S15:** Violinplots visualize differences in community variability between the main health conditions (Table S20), as expressed by the within group distance to the centroid in NMDS of taxon and function based Bray-Curtis distances. (via *betadisper* function). Global P-values were derived via permutation test of multivariate homogeneity of group dispersions (10000 permutations). Overall, we can see a significant increas of community variability in CD and UC patients compared to healthy controls.

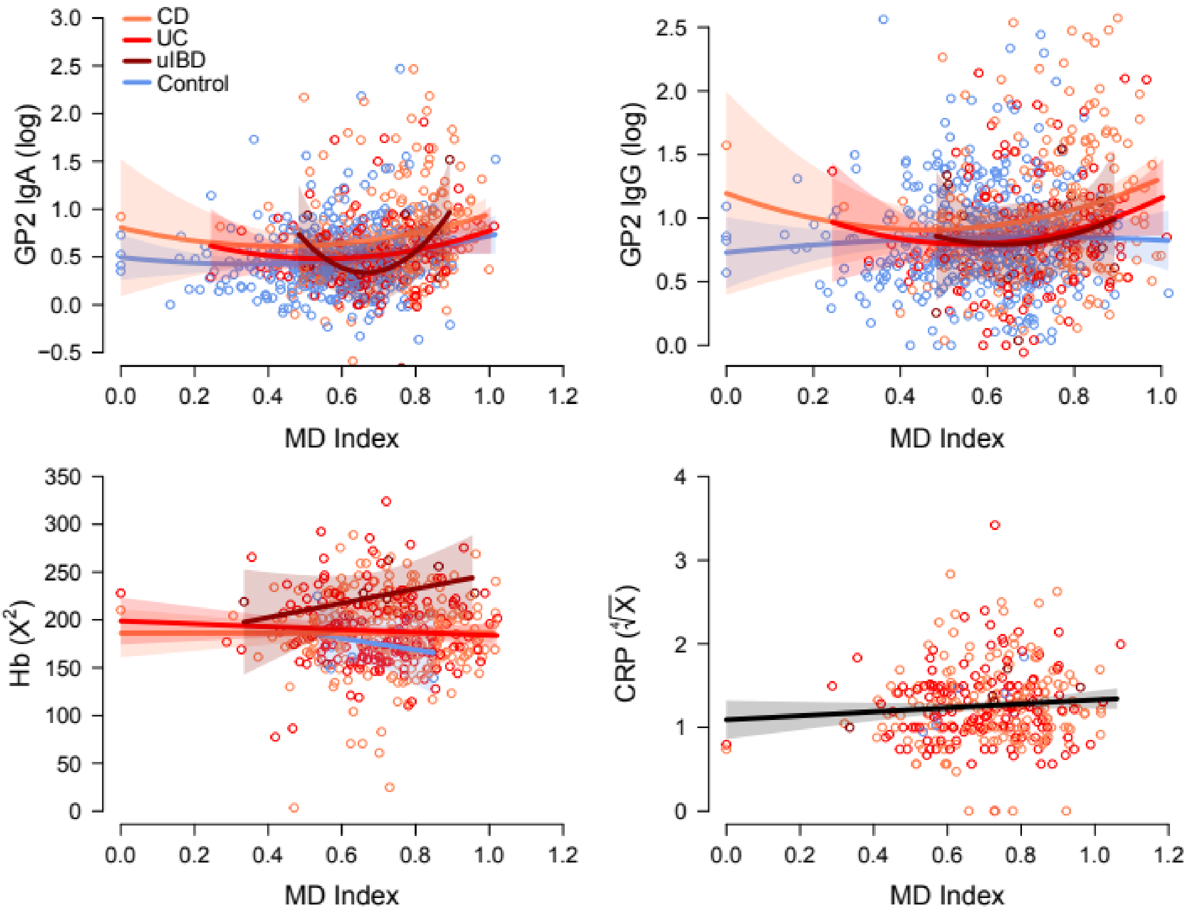


**Figure S16:** Association of immune / physiological parameters with the microbial dysbiosis index (Gevers et al. 2014) using LMs including relevant covariates (age, BMI, gender) after model selection (see Table S18). Polygons highlight the 95% CI.

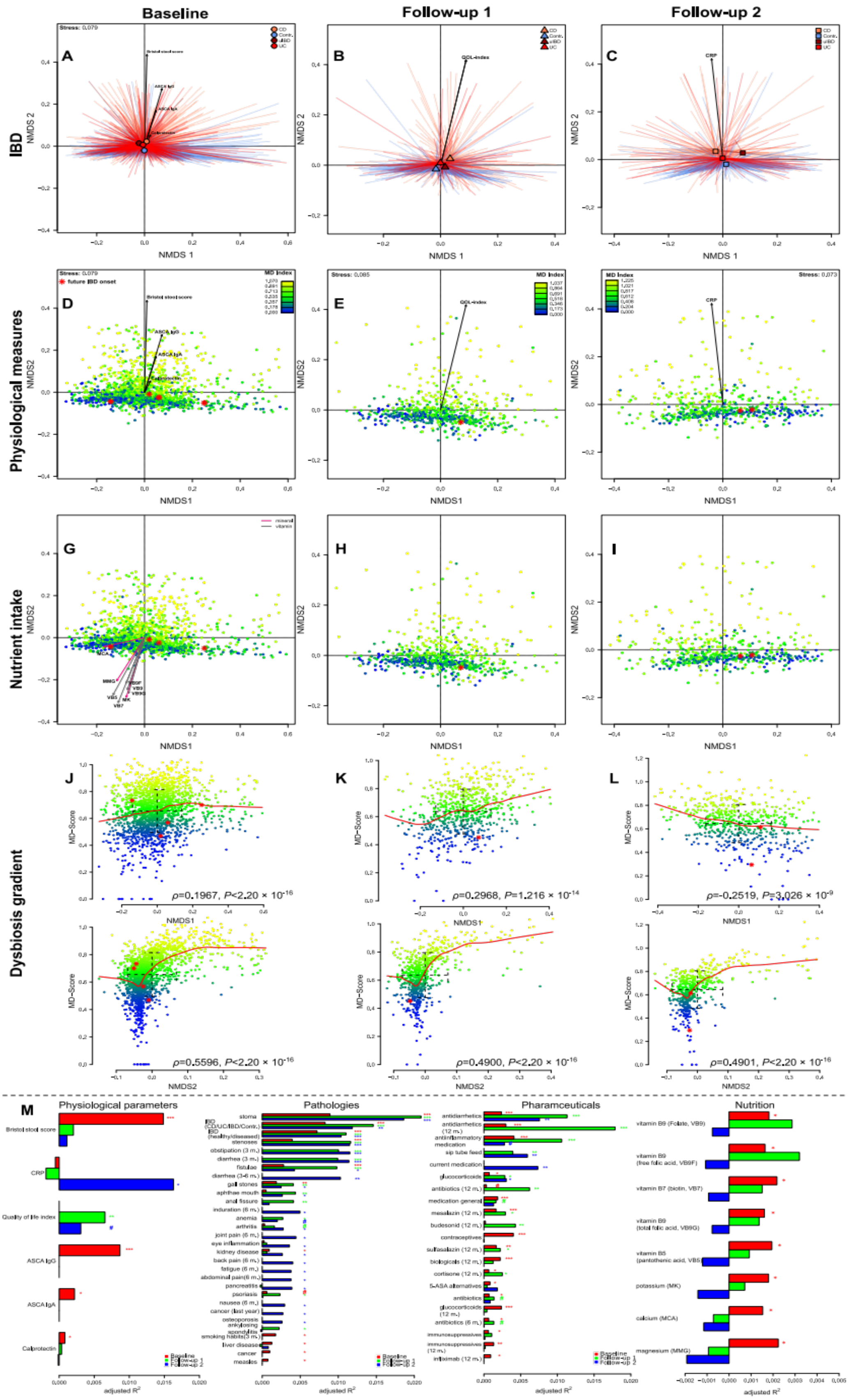


**Figure S17: (A)** Non-metric Multidimensional Scaling (NMDS) of Bray-Curtis distances of PICRUSt2 based KO abundances among baseline samples [13], **(B)** follow-up 1, and **(C)**

follow-up 2, displaying the significant clustering by health conditions and significant correlations of clinical inflammation measures with community distance (see Table S16, Table S17, Table S23)). **(D, E, F)** NMDS displaying the gradient of community dysbiosis as expressed by the taxonomy based MD-index [8], across the three time points, in parallel with significant clinical measures of inflammation correlated to the functional differences among communities. Healthy IBD onset cases are highlighted in red (*, develop IBD until the next follow-up). **(G, H, I)** Nutrition (approximated normalized nutrient uptake) was also significantly correlated with functional community distances. Arrow colors represent different nutrient groups (AA-aminoacids, FA-fatty acids, carbon-carbohydrates, minerals-trace elements, vitamins, makro nutrients-larger nutrient clusters (i.e. proteins, fats, water)). **(J, K, L)** Correlation of MD-index and the first and second NMDS axes show a clear gradient of dysbiosis in the community (Spearman rank correlation). Onset cases are distributed in the range of standard deviation around the mean of the community distribution (NMDS1, NMDS2) and the severity of dysbiosis (MD-index).**(M)** Visualization of the explained variation of significant anthropometric variables as based on serial PERMANOVA of Bray-Curtis distances in all three time points available and focused on physiological measures, different reported pathologies of individuals, use of pharmaceuticals, and nutrient intake as derived from FFQ data. Variables are displayed if they show significant clustering in at least one time point (# $P_{FDR} \leq 0.1000$, * $P_{FDR} \leq 0.0500$, ** $P_{FDR} \leq 0.0100$, *** $P_{FDR} \leq 0.0010$).

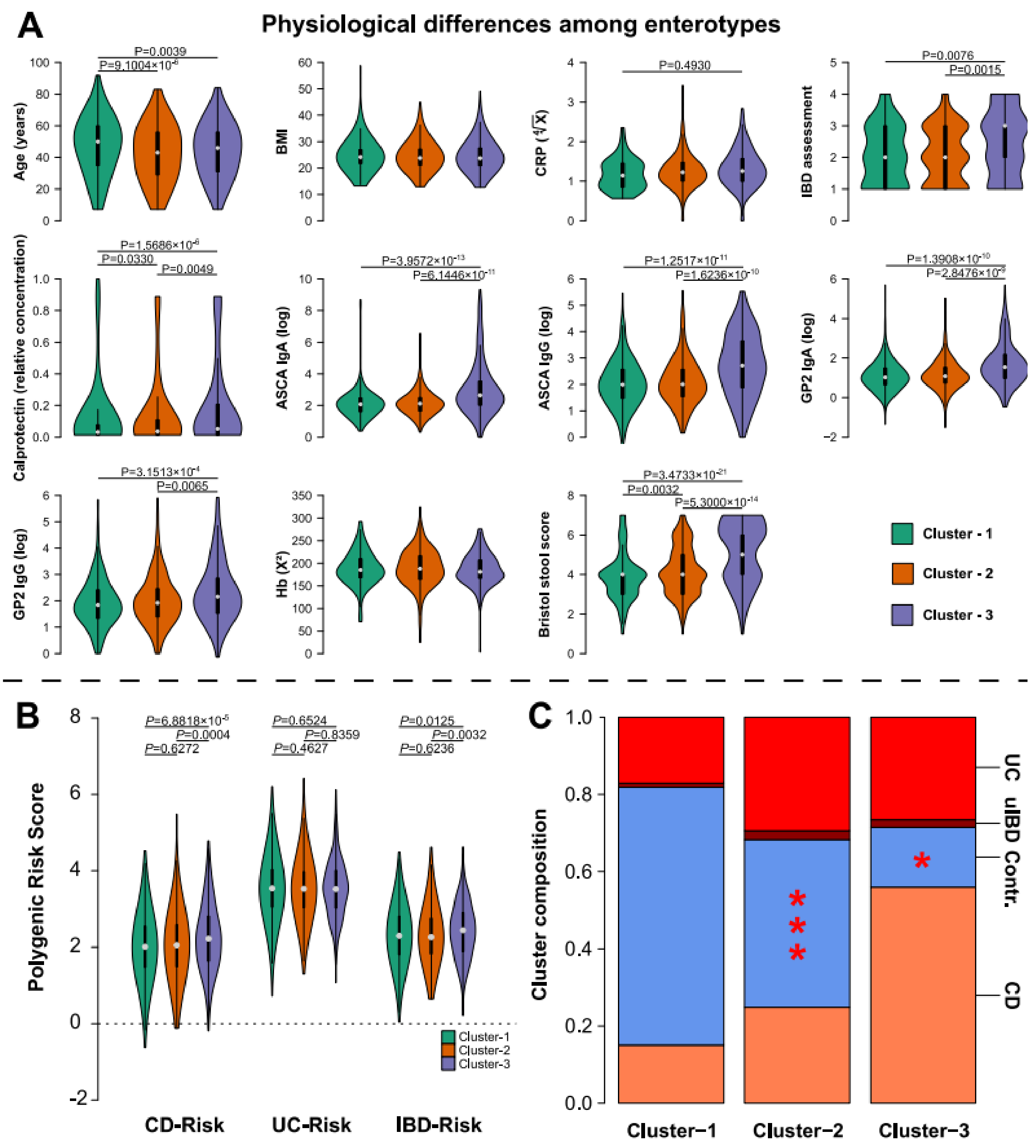


**Figure S18: (A)** Violinplots visualize the differences of inflammation related continuous and physiological variables between community clusters, whith elevated levels of inflammatory biomarkers in cluster-3 (pairwise Wilcoxon test). **(B)** Differences of polygenic risk scores for CD, UC or general IBD between the three microbiome community clusters at baseline. We can see a clearly increased PRS in the Proteobacteria dominated community cluster-3 (pairwise Wilcoxon test). **(C)** Barplots visualize the distribution of health conditions/pathologies between community clusters and the distribution of future IBD onset patients ("*").

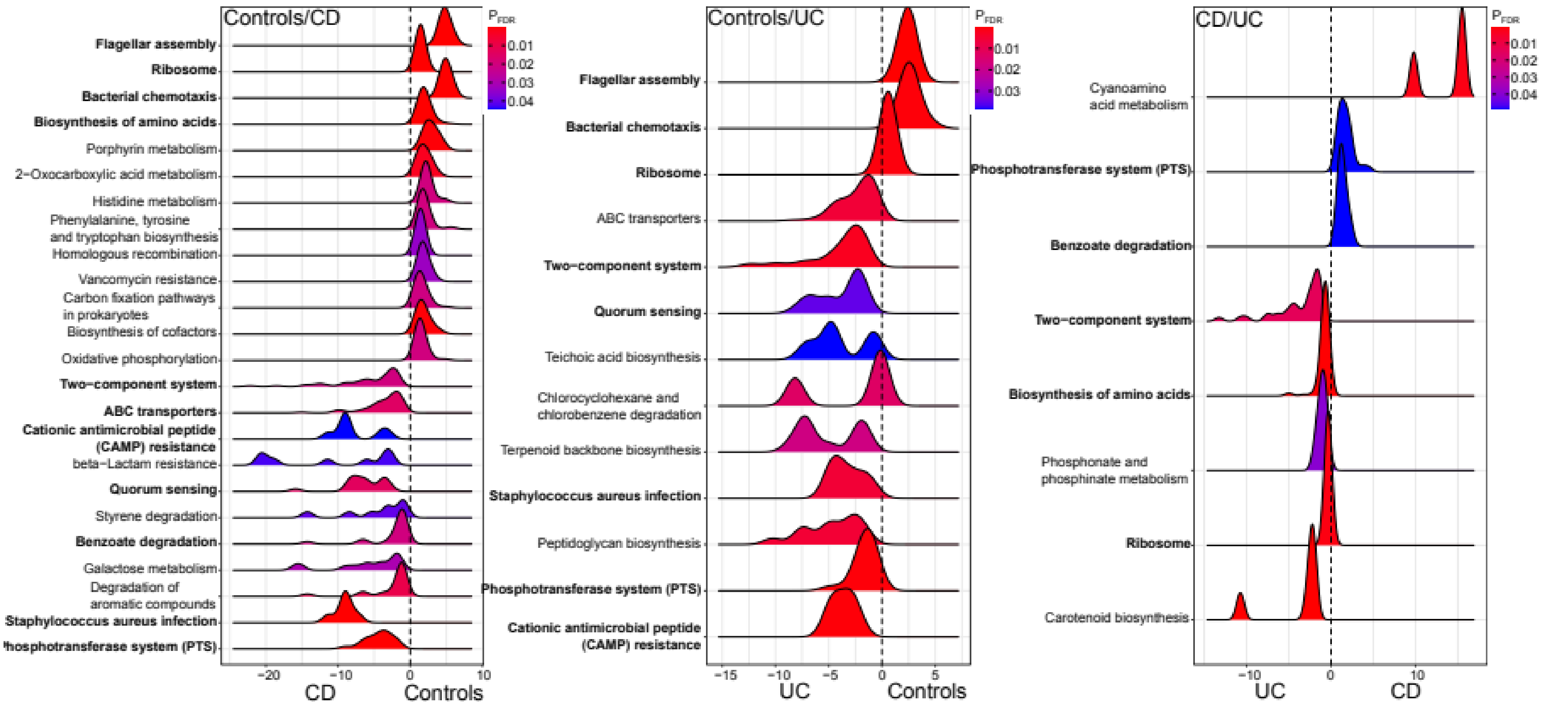


**Figure S19:** Significant functional enrichments in the Maltese, treatment naive IBD cohort, based on signficantly differential abundant Kos between single health conditions. Repeatedly detected metabolic pathways are highlighted in bold. Differential abundance was tested via *DESeq2* and the enrichment score was derived from -log10(*P*-values)×direction of fold change.

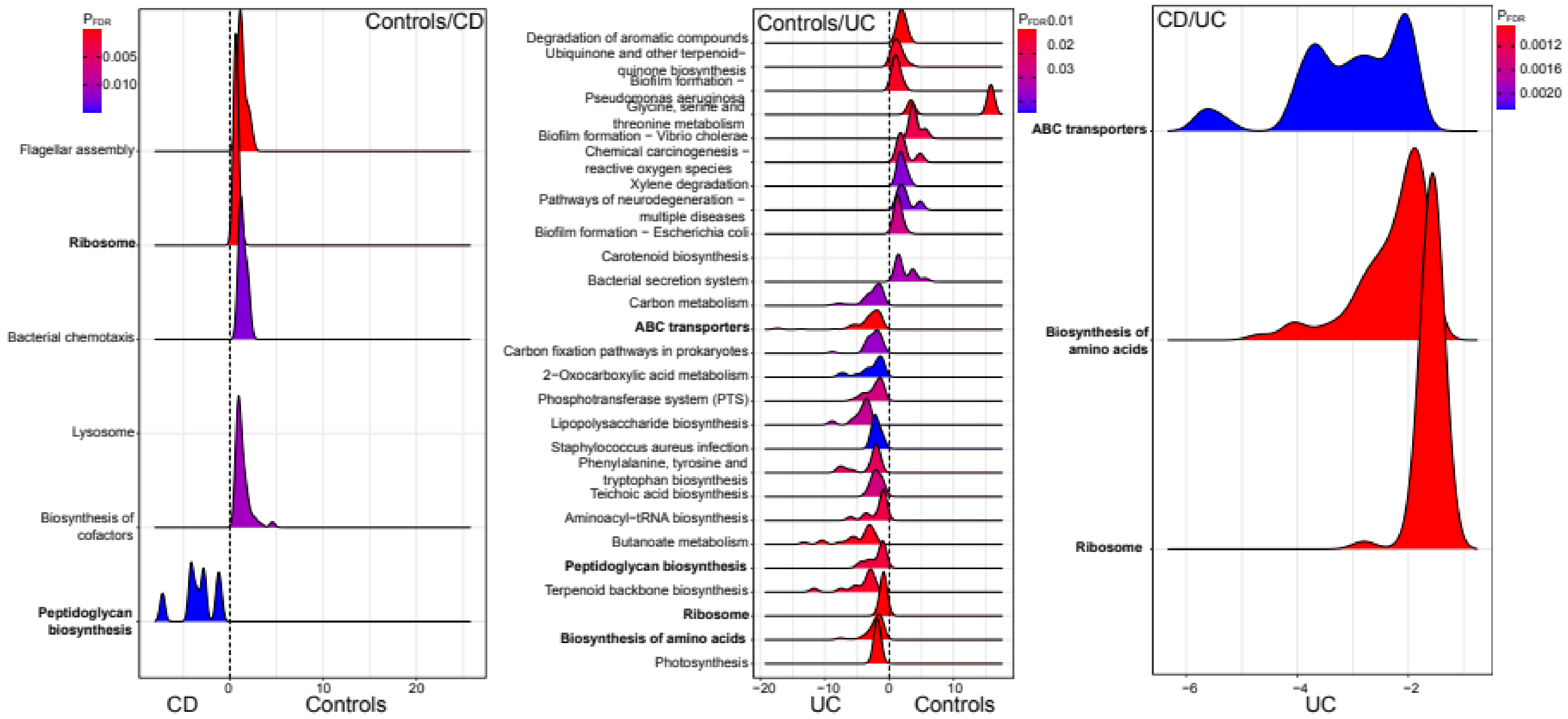


**Figure S20:** Significant functional enrichments in the Maltese IBD cohort in disease remission, based on signficantly differential abundant Kos between single health conditions. Repeatedly detected metabolic pathways are highlighted in bold. Differential abundance was tested via *DESeq2* and the enrichment score was derived from -log10(*P*-values)×direction of fold change.

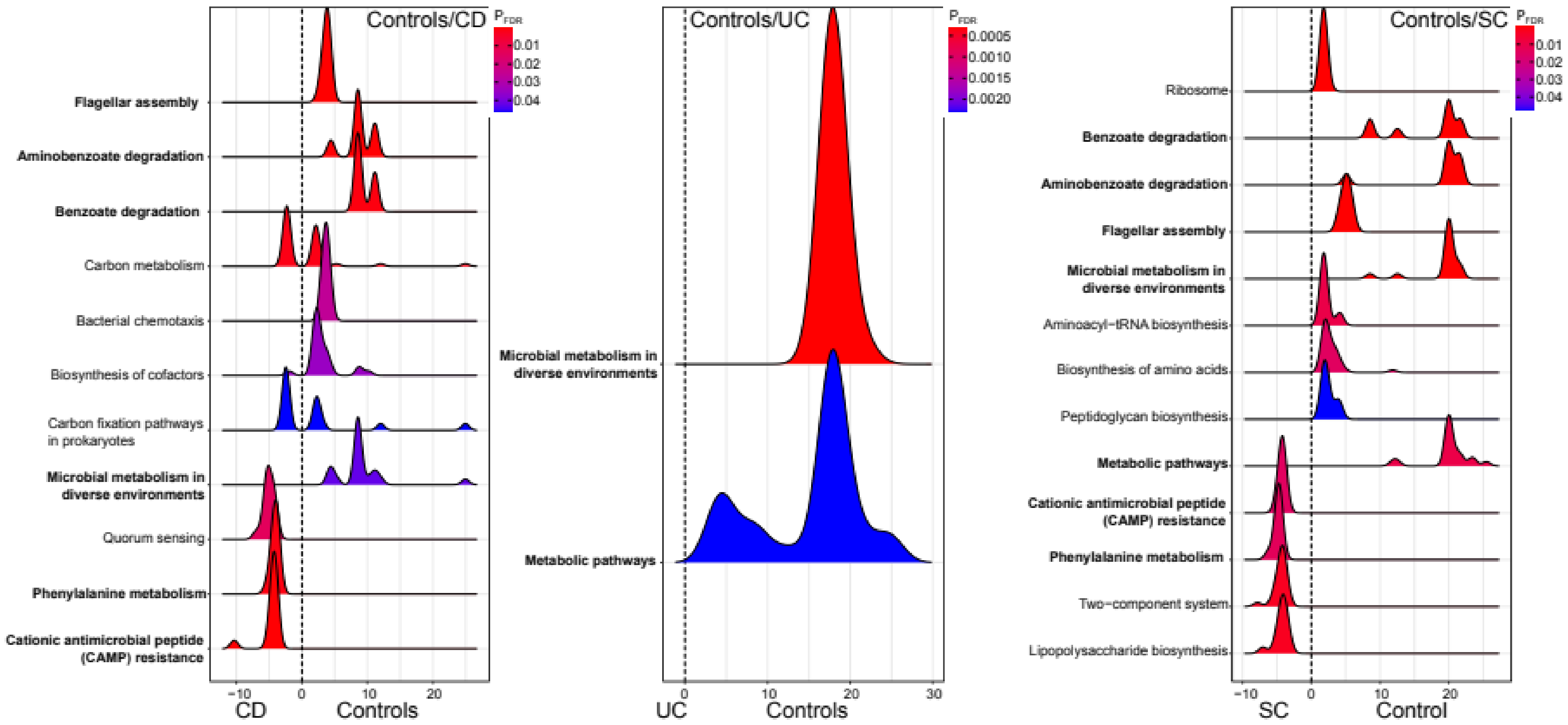


**Figure S21:** Significant functional enrichments in the treatment naive, Swedish IBD cohort, based on signficantly differential abundant Kos between single health conditions. Repeatedly detected metabolic pathways are highlighted in bold. Differential abundance was tested via *DESeq2* and the enrichment score was derived from -log10(*P*-values)×direction of fold change.

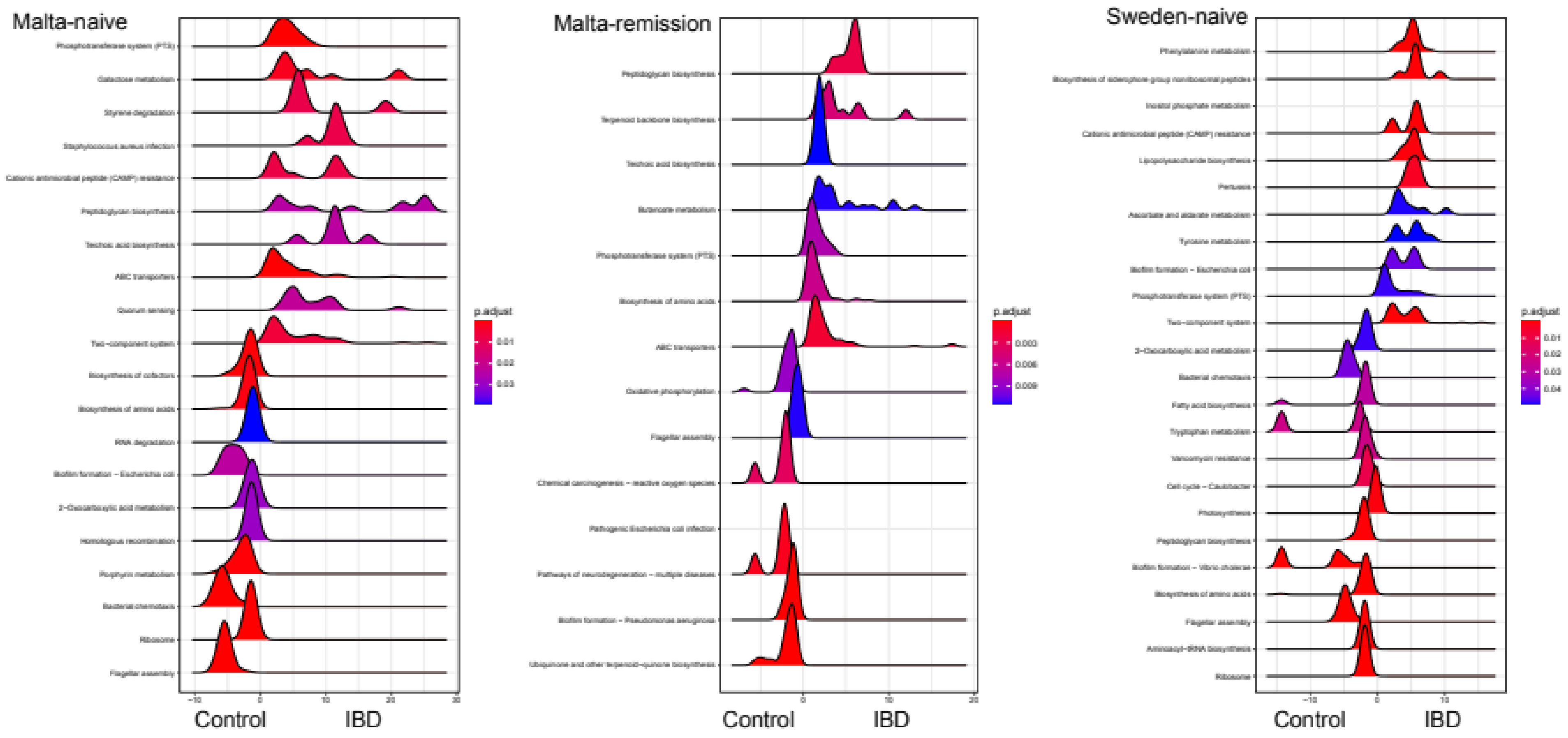


**Figure S22:** Significant functional enrichments in the treatment naive, Maltese IBD cohort, Maltese remission cohort, and treatment naive Swedish cohort, based on signficantly differential abundant Kos between healthy controls (+SC) and IBD conditions. Enrichments of repeated metabolic pathways are highlighted in bold. Differential abundance was tested via *DESeq2* and the enrichment score was derived from -log10(*P*-values)×direction of fold change.

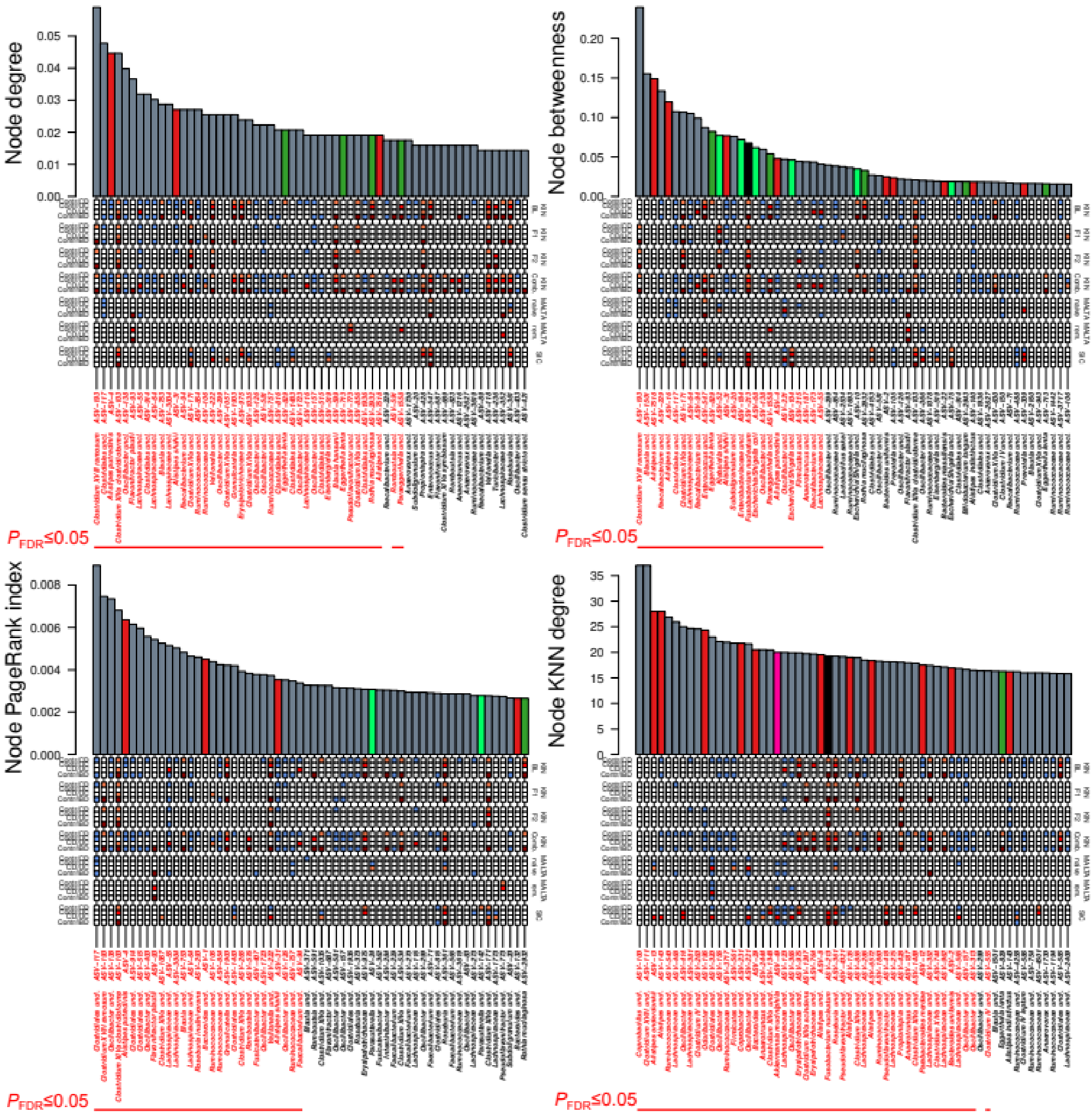


**Figure S23:** Network importance/centrality measures derived at the baseline time point and highlighting differential abundance associations at each node as derived from *DESeq2*. Centralities range from the number of connections (degree), the position on the shortest paths within the network (betweeness, [14]), generalized importance (PageRank index [15]) and the average neighborhood degree of any given vertex (k-nearest neighbor degree [16]). Colored boxes highlight significant associations in KIN and external cohorts with IBD pathologies or IBD onset. Significance of centralities is derived from Z-test against a large collection of randomized centralities of the network and ASV names in red highlight significantly higher than random network importance (FDR corrected, Table S25).

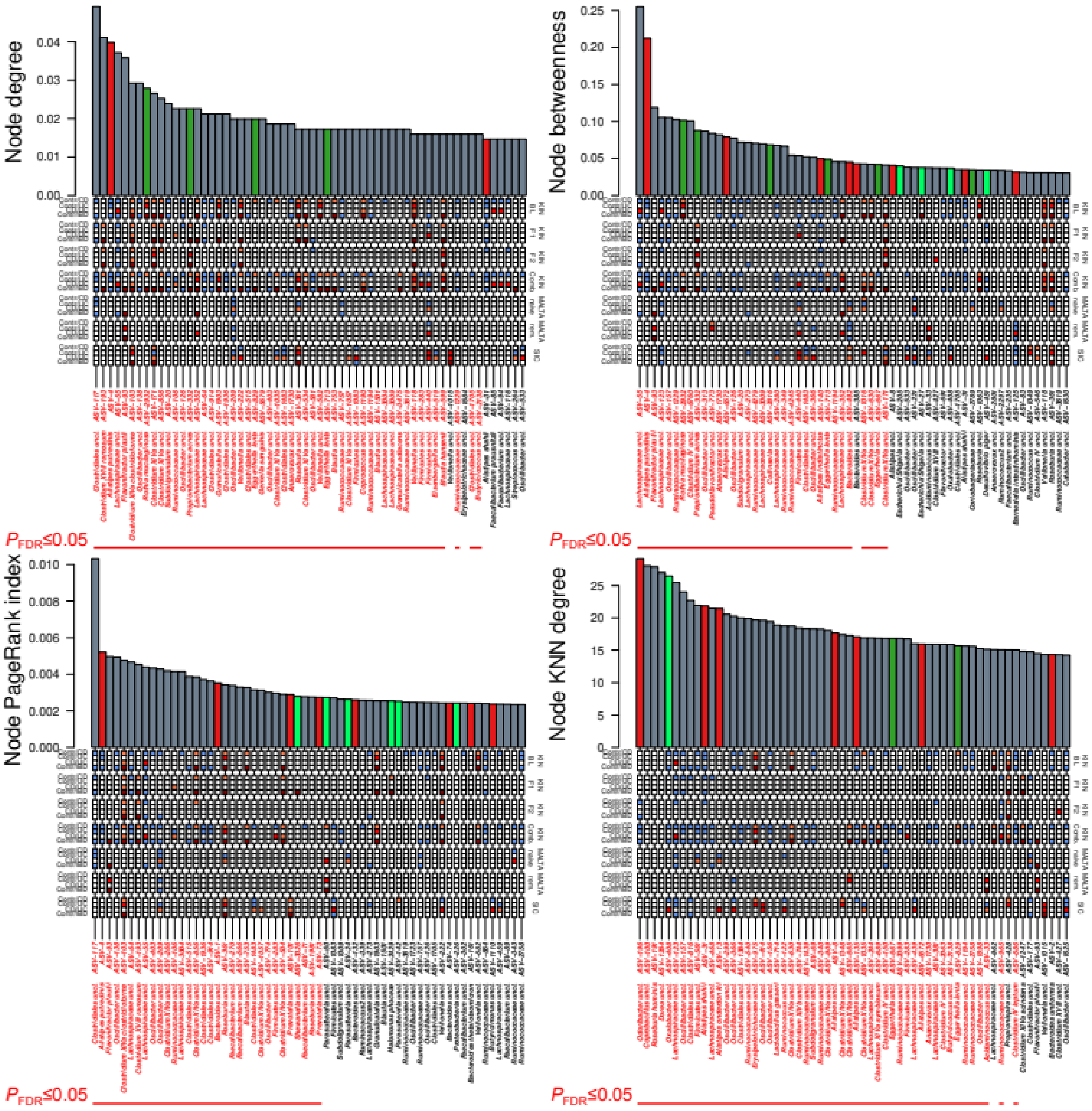


**Figure S24:** Network importance measures derived at the first follow-up time point and highlighting differential abundance associations at each node as derived from DESeq2. Centralities range from the number of connections (degree), the position on the shortest paths within the network (betweeness, [14]), generalized importance (PageRank index [15]) and the average neighborhood degree of any given vertex (k-nearest neighbor degree [16]). Colored boxes highlight significant associations in KIN and external cohorts with IBD pathologies or IBD onset. Significance of centralities is derived from Z-test against a large collection of randomized centralities of the network and ASV names in red highlight significantly higher than random network importance (FDR corrected Table S25).

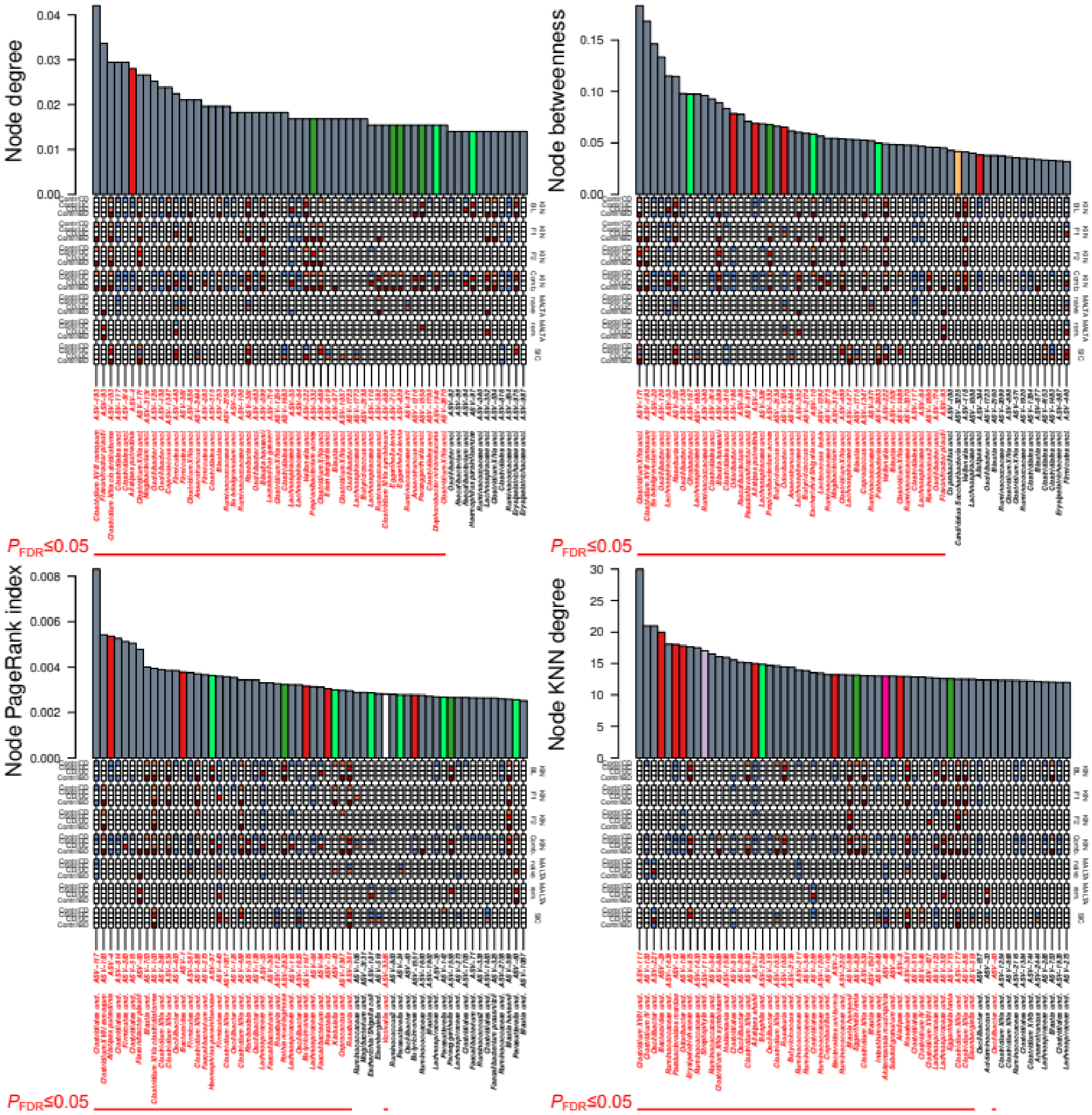


**Figure S25:** Network importance measures derived at the second follow-up time point and highlighting differential abundance associations at each node as derived from DESeq2. Centralities range from the number of connections (degree), the position on the shortest paths within the network (betweeness, [14]), generalized importance (PageRank index [15]) and the average neighborhood degree of any given vertex (k-nearest neighbor degree [16]). Colored boxes highlight significant associations in KIN and external cohorts with IBD pathologies or IBD onset. Significance of centralities is derived from Z-test against a large collection of randomized centralities of the network and ASV names in red highlight significantly higher than random network importance (FDR corrected Table S25).

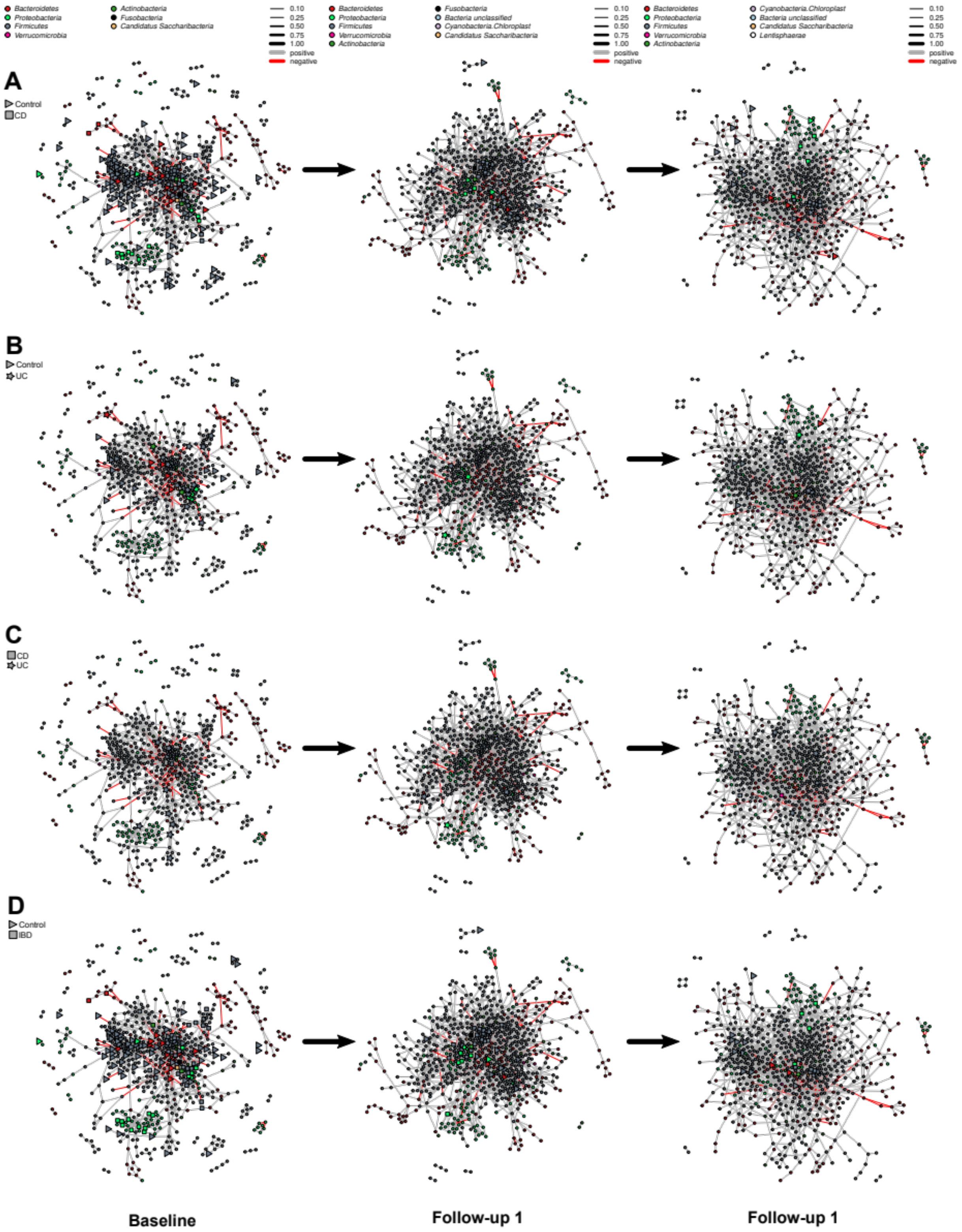


**Figure S26:** Spiec-Easi networks of baseline samples, follow-up 1, and follow-up 2. Bacterial nodes highlight significant differentially abundant ASVs in the network. **(A)** Bacteria not showing any differential abundance patterns between CD patients and healthy controls are signified via (●), bacteria overabundant in CD via (■) and bacteria more abudnant in controls are signified via (►). **(B)** Bacteria not showing any differential abundance patterns between UC patients and healthy controls are signified via (●), bacteria overabundant in UC via (★) and bacteria more abudnant in controls are signified via (►). **(C)** Bacterial nodes highlight significant differentially abundant ASVs in the network. Bacteria not showing any differential abundance patterns between UC patients and healthy controls are signified via (●), bacteria overabundant in UC via (★) and bacteria more abudnant in CD are signified via (■). **(D)** Bacterial nodes highlight significant differentially abundant ASVs in the network. Bacteria not showing any differential abundance patterns between UC patients and healthy controls are signified via (●), bacteria overabundant in healthy controls via (►) and bacteria more abudnant in IBD (CD+UC) are signified via (■).

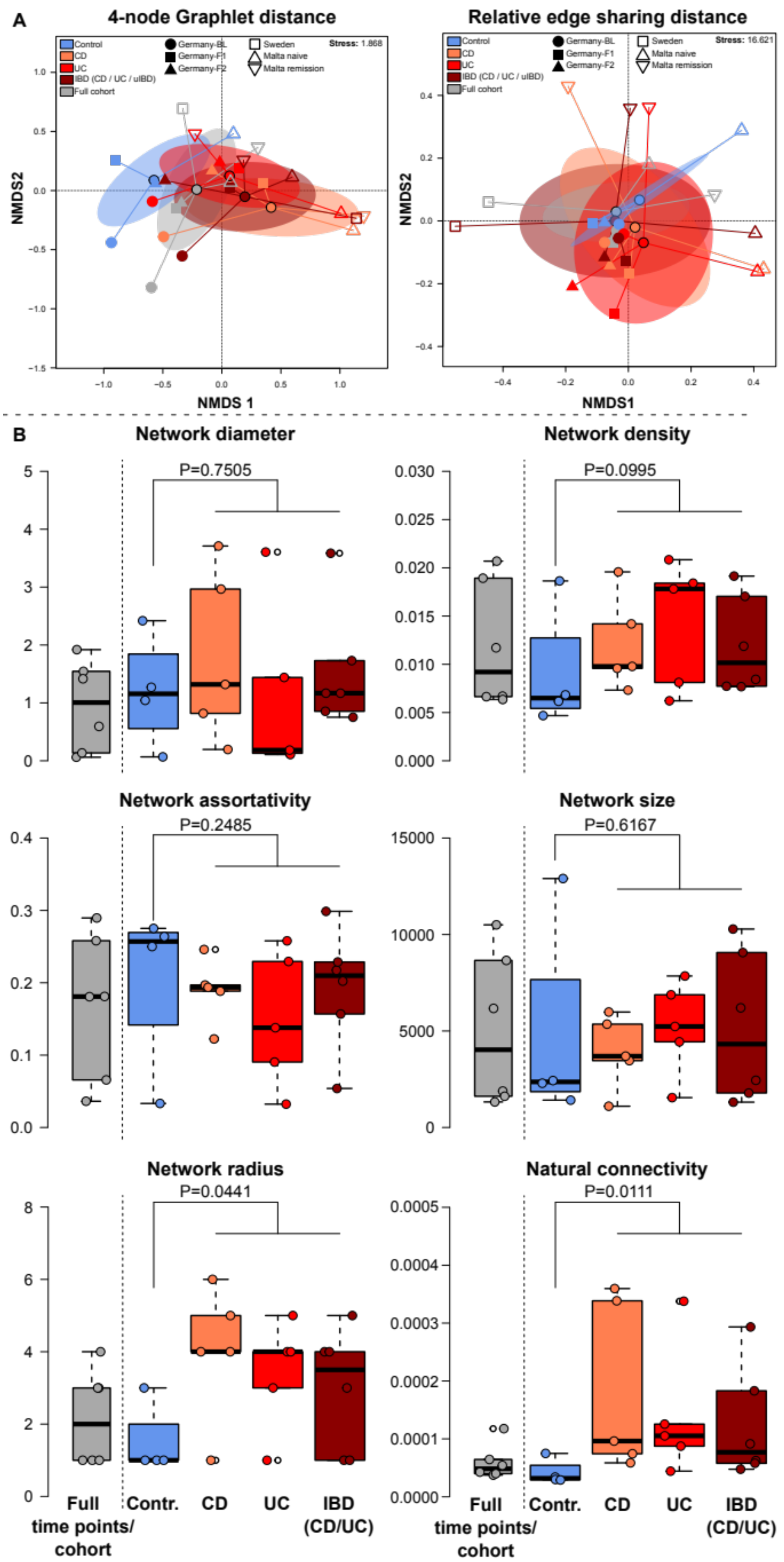


**Figure S27: (A)** Network similarity of disease and time point specific sub networks, as well as subnetworks of external cohorts (Malta, Sweden) based on graphlet distance (Yaveroğlu et al. 2014) and relative edge sharing distance, displayed via NMDS. Networks show a clear compositional difference between healthy and diseased networks as based on graphlet distance (Control *vs.* IBD (incl. IBD networks): $F_{1,18}$=2.4144, $P$=0.0412, $R^2$=0.1183, *adj.* $R^2$=0.0693; PERMANOVA) and relative edge sharing distance (KINDRED only- Control *vs*. IBD: $F_{1,7}$=1.0775, $P$=0.0621, $R^2$=0.1334, *adj.* $R^2$=0.0096; all cohorts (Contr., CD, UC)- Control *vs*. IBD: $F_{1,12}$=1.1096, $P$=0.0935, $R^2$=0.0846, *adj.* $R^2$=0.0084; all cohorts (Contr., CD, UC, IBD)- Control *vs*. IBD (incl. general IBD networks): $F_{1,18}$=1.1201, $P$=0.0853, $R^2$=0.0586, *adj.* $R^2$=0.0063; PERMANOVA). **(B)** Global network characteristics derived from KINDRED, Malta IBD-naive, Malta IBD-remission, and Sweden-SIC cohorts. Significance is based on Wilcoxon tests between healthy control based networks and networks derived from diseased individuals (CD, UC, IBD). Assortativity [17], diameter, radius, and size [18], density [19], natural connectivity [20]. Full time point specific (KINDRED BL, KINDRED F1, KINDRED F2) and cohort specific (Malta IBD-naive incl. controls, Malta IBD-remission incl. controls, Sweden-SIC cohorts)

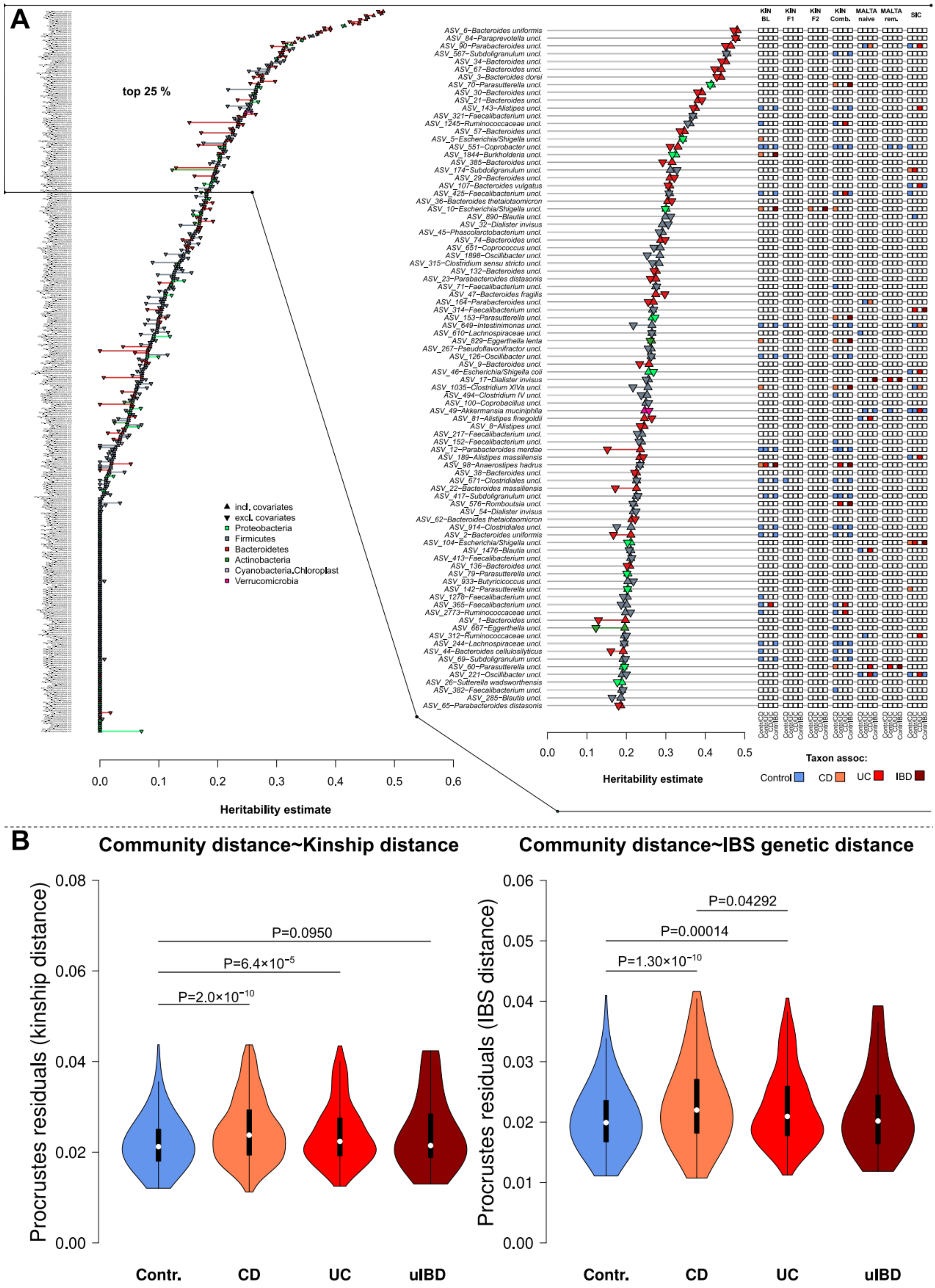


**Figure S28: (A)** Heritability estimates derived from the likelihood based method *lme4qtl* [21] using either only kinship information with or without additional environmental and anthropometric covariates. The upper 25% percentile of taxa are highlighted (based on $h^2$ estimate including environmental covariate). Additional information like differential abundance in IBD accross cohorts, as well as their association to IBD onset or remission are depicted (Table S27). **(B)** Comparison of community distance and topography (Bray-Curtis distance ASV) with kinship distances and IBS genetic distance based on Procrustes superimposition [22]ckson, 2001). Divergence between community distance and relatedness based on procrustes residuals, shows a higher divergence in IBD cases, and thus a stronger divergence from the inheritance and transmission patterns of the microbiome (pairwise Wilcoxon tests).

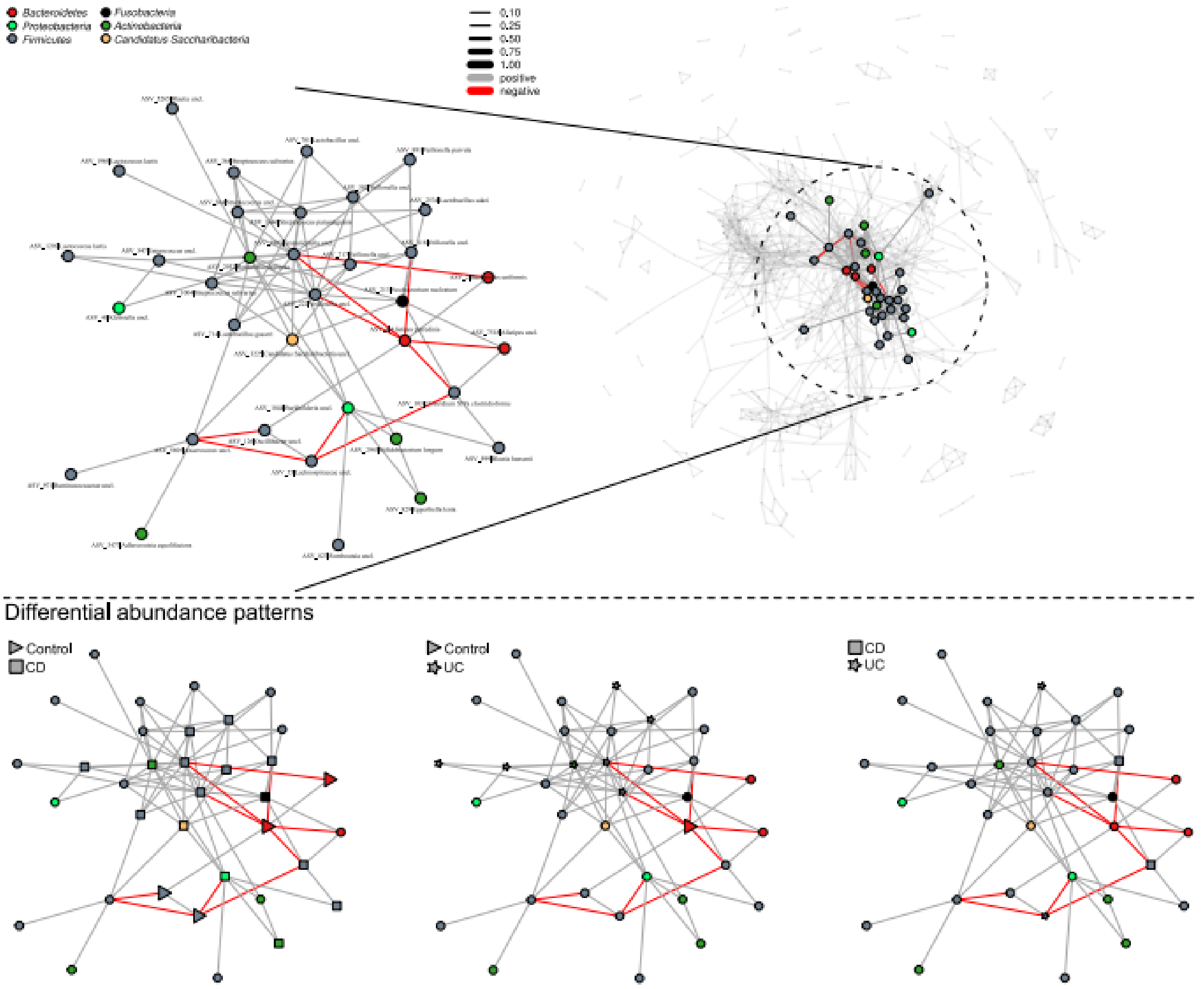

Figure S29: Subnetwork at baseline based on Cand. Saccharibacteria and its 1st and 2nd order naighbourhood, including other oral taxa such as *Veillonella*, *Granulicitella*, *Klebsiella*, *Rothia*, *Fusobacteria*, or *Streptococcus*, and their significant differential abundance patterns (Table S8).

**Supplemental tables:**

**Table S1:** Overview of items included in the different questionnaires of the Kiel IBD Family Cohort BL=baseline assessment; F1=since first follow-up assessment; F2=since second follow-up assessment. Abbreviations: CDAI, Crohn's Disease Activity Index; FSS, Fatigue Severity Scale; HBI, Harvey-Bradshaw-Index; IBD, inflammatory bowel disease.)

**Table S2:** Overview of biomaterial sample collection, processing, and storage time between processing and storage.

**Table S3:** Distribution of IBD types within the IBD patients (n=1321) of the Kiel IBD Family Cohort. Abbreviations: CD, Crohn's disease; IBD, inflammatory bowel disease; UC, Ulcerative colitis; uIBD, undefined inflammatory bowel disease.

**Table S4:** Baseline characteristics of unaffected (healthy) relatives of IBD patients (n=1072) in the Kiel IBD Family Cohort, stratified by age group. Values are median (IQR) or absolute and relative frequencies. Abbreviations: IBD, inflammatory bowel disease.

**Table S5:** Linear model analyses of clinical inflammation markers with respect to *LDpred2* Polygenic Risk Scores (CD, UC and general IBD), IBD pathology, and other covariates. Analyses were performed on baseline samples (BL).

**Table S6:** Analysis of disease onset prediction via logistic regression of PRS estimates, using models accounting or not accounting for covariates (age, BMI, sex).

**Table S7:** Analyses of differential phylum abundance (RPD16 based) via negative binomial models (DESeq2, Wald test), between control individuals, CD and UC patients (excluding uIBD) and between control individuals, IBD patients (CD, UC, uIBD patients). Models were adjusted by scaled age, scaled BMI and sex as covariates. The three sampling time points were analysed separately and all P values were adjusted via FDR.

**Table S8:** Differential abundance analyses of baseline samples (BL) at the ASV level (RPD16 based classification included) via negative binomial models (DESeq2, Wald test), between control individuals, CD and UC patients (excluding uIBD), and between control individuals and IBD patientis (including uIBD). Models were adjusted by age, BMI and sex as covariates. All P values were adjusted via FDR. Additional support from external independent cohorts is included based on DA following the same methodology (Malta treatment naive, Malta remission, Sweden-SIC).

**Table S9:** Differential abundance analyses of samples from the first follow-up (F1) at the ASV level (RPD16 based classification included) via negative binomial models (*DESeq2*, Wald test), between control individuals, CD and UC patients (excluding uIBD), and between control individuals and IBD patientis (including uIBD). Models were adjusted by age, BMI and sex as covariates. All P values were adjusted via FDR. Additional support from external independent cohorts is included based on DA following the same methodology (Malta treatment naive, Malta remission, Sweden-SIC).

**Table S10:** Differential abundance analyses of samples from the second follow-up (F2) at the ASV level (RPD16 based classification included) via negative binomial models (*DESeq2*, Wald test), between control individuals, CD and UC patients (excluding uIBD), and between control individuals and IBD patientis (including uIBD). Models were adjusted by age, BMI and sex as covariates. All P values were adjusted via FDR. Additional support from external independent cohorts is included based on DA following the same methodology (Malta treatment naive, Malta remission, Sweden-SIC).

**Table S11:** Differential abundance analyses of combined samples (BL, F1, F2) at the ASV level (RPD16 based classification included) via negative binomial models (*DESeq2*, Wald test), between control individuals, CD and UC patients (excluding uIBD), and between control individuals and IBD patientis (including uIBD). Models were adjusted by age, BMI, sex, and time point as covariates. All P values were adjusted via FDR. Additional support from external independent cohorts is included based on DA following the same methodology (Malta treatment naive, Malta remission, Sweden-SIC).

**Table S12:** Partial correlation of CLR transformed taxon abundances with core physiological/clinical measures and *LDpred2* derived polygenic risk scores (PRS) for CD, UC, and IBD via *ppcor* (Kim 2015). P-values were derived from combining the *P* values of Spearman-, Kendall-, and Pearson correlations via Brown's method and corrected via FDR (Brown 1975). Correlations were adjusted for age, gender and BMI. The table includes additional information of overlapping and significant differential abundance patterns in the KINDRED cohort, Maltese-, and Swedish SIC cohort.

**Table S13:** Correlation of alpha diversity with MD and GMHI across baseline, follow-up 1, and follow-up 2 using either a linear or polynomial (quadratic) fit, as based on minimal AIC. Models were either adjusted for covariates or without.

**Table S14:** Linear model analyses of alpha diversity and MD-index in relation to IBD condition, including differences in average diversity changes as well as of the disease specific correlation between alpha diversity and MD.

**Table S15:** Analyses of the relationship between clinical markers of inflammation (ASCA IgA/IgG, GP2 IgA/IgG, calprotectin, Bristol stool score, Hb, CRP, IBD severity (clinician)), alpha diversity measures (Chao1 species richness, Shannon Diversity (effective number), Net relatedness index, Nearest taxon index). Analyses were performed on the residuals of the respective markers, after fitting a linear model including age, BMI, and sex.

**Table S16:** Betadiversity analyses of Bray-Curtis dissimilarity based on ASV and PICRUSt2-KO abundances. Analyses were done using PERMANOVA either with or without conditioning for potential covariates (age, BMI, sex). Analyses were performed globally, pairwise, as well dysbiosis scores in the different sampling time points (Baseline, Follow-up 1, Follow-up 2). Pairwise comparisons were corrected for multiple testing via FDR.

**Table S17:** Betadiversity analyses of Bray-Curtis dissimilarity based on ASV and PICRUSt2-KO abundances focusing on physiological and anthropometric measures at different sampling time points (Baseline, Follow-up 1, Follow-up 2).

**Table S18:** Linear model analyses of dysbiosis scores with respect to IBD pathology, and other covariates. Analyses were performed on the residuals of the respective markers, after fitting a linear model including age, BMI, and sex and then selected by minimizing AIC.

**Table S19:** Pairwise PERMANOVA analyses of ASV based Bray-Curtis dissimilarities of combined KINDRED, Maltese, and Swedish cohorts, based on the main pathologies.

**Table S20:** Community variability analysis of Bray-Curtis dissimilarity via PERMANOVA, as based on ASV and PICRUSt2-KO abundances focusing on IBD pathologies across sampling time points (Baseline, Follow-up 1, Follow-up 2).

**Table S21:** Betadiversity analysis of Bray-Curtis dissimilarity based on ASV and PICRUSt2-KO abundances focusing on pathologies and morbidities reported by the subjects across sampling time points (Baseline, Follow-up 1, Follow-up 2).

**Table S22:** Betadiversity analyses of Bray-Curtis dissimilarity based on ASV and PICRUSt2-KO abundances focusing on medical/pharmaceutical treatments reported by the subjects across sampling time points (Baseline, Follow-up 1, Follow-up 2). Naive and conditioned PERMANOVA results.

**Table S23:** Betadiversity analyses of Bray-Curtis dissimilarity based on ASV and PICRUSt2-KO abundances focusing on normalized nutrient intake derived from 2 week food frequency questionnaires, across sampling time points (Baseline, Follow-up 1, Follow-up 2). Naive and conditioned PERMANOVA results.

**Table S24:** Gene set enrichment analyses (GSEA) based on differential abundance of PICRUSt2 KOs with respect to IBD pathology among the different time points. GSEA derived from external cohorts are included for additional support ($P_{FDR} \leq 0.05$) [23].

**Table S25:** Network importance/centrality measures derived at the different time points (BL, F1, F2) of the KINDRED cohort, as well as subsets by helath condition (CD, UC, Controls) within each time point. Results of differential abundance associations, as well as potential role in disease onset or remission, for each significant node/taxon are included. Centralities range from the number of connections (degree), the position on the shortest paths within the network (betweeness, [14], generalized importance (PageRank index [15] and the average neighborhood degree of any given vertex (k-nearest neighbor degree [16]. Significance of centralities is derived from Z-test against a collection of randomized network centralities (FDR corrected). Taxa which are significantly more central than expected by chance, and consistently over-abundant in healthy individuals are highlighted in lightblue, taxa consistently over-abundant in IBD cases (CD/UC) are highlighted in orange.

**Table S26:** Taxon heritability as derived from linear mixed models including kinship matrices. Heritability ($h^2$) was estimated with and without environmental variables via *lme4qtl*. The table includes differential abundance patterns in KINDRED and external cohorts (incl. their summary). Values indicating a better fit solely by pedigree information (yellow) or better fit when covariates (sex, age, BMI, IBD pathology) are included in addition to pedigree information.
